# Interpretable machine learning classifiers implicate *GPC6* in Parkinson’s disease from single-nuclei midbrain transcriptomes

**DOI:** 10.1101/2024.11.19.24317547

**Authors:** Michael R. Fiorini, Jialun Li, Edward A. Fon, Sali M.K. Farhan, Rhalena A. Thomas

## Abstract

Parkinson’s disease (PD) is a progressive and devastating neurodegenerative disease. An incomplete understanding of its genetic architecture remains a major barrier to the clinical translation of targeted therapeutics, necessitating novel approaches to uncover elusive genetic determinants. Single-cell and single-nuclear RNA sequencing (scnRNAseq) can help bridge this gap by profiling individual cells for disease-associated differential gene expression and nominating genes for targeted genomic analyses. Here, we introduce a machine learning framework to identify molecular features that characterize post-mortem brain cells from PD patients. We train classifiers to distinguish between PD and healthy cells, then decode the models to unravel the ‘reasons’ behind the classifications, revealing key genes expression signatures that characterize cells from the parkinsonian brain. Application of this framework to three publicly available snRNAseq datasets characterizing the post-mortem midbrain identified cell-type-specific gene sets that accurately classify PD cells across all datasets, demonstrating our approach’s capacity to identify robust molecular markers of disease. Targeted genomic analyses of the key genes characterizing PD cells revealed a previously undescribed association between PD and rare variants in *GPC6*, a member of the heparan sulfate proteoglycan family, which have been implicated in the intracellular accumulation of α-synuclein preformed fibrils. We replicate this association in three separate case-control cohorts. Our method promises to enhance understanding of the genetic architecture in complex diseases like PD, representing a critical step toward targeted therapeutics. Our publicly available framework is readily applicable across diseases.

## Introduction

Parkinson’s disease (PD) is a neurodegenerative disease marked by motor symptoms such as slow movement, shuffling gait, tremor, and balance disturbances (1). Pathological hallmarks of PD include the presence of Lewy bodies, which are primarily composed of α-synuclein aggregates (2), and the loss of dopaminergic neurons (DaNeurons) in the substantia nigra pars compacta (SNpc) of the midbrain (3). PD affects approximately 1% of the population over the age of 60 (4) and displays heritability estimates around 30% (5). Only 5-10% of cases are attributed to monogenic variants in genes known to cause a familial history of PD, including *PRKN*, *PINK1*, *PARK7*, *SNCA*, and *LRRK2* (6, 7). The remaining cases likely result from the complex interplay between genetic variants that confer an increased risk of PD (8, 9) and environmental exposures, including pesticides (10, 11). The etiology of PD is further complicated by considerable clinico-pathological heterogeneity amongst patients including variable ages of onset, symptoms, and progression rates (5). Despite advancements in our understanding of the genetic architecture of PD, it is currently estimated that only 16-36% of its heritability is accounted for by known PD-associated genes (5), necessitating the use of novel methodologies to uncover elusive genetic determinants. Indeed, a comprehensive understanding of the genetic basis of PD will advance our understanding of disease biology and facilitate the clinical translation of findings to targeted therapeutics. At present, the disease remains relentlessly progressive as there are no disease-modifying treatments (12).

Transcriptomic analysis, which facilitates transcriptome-wide differential gene expression (DGE) profiling, is a promising approach to bridge the gap between genetics and disease biology. By comparing samples from individuals with disease to samples from healthy controls, we can investigate how the disease process influences gene expression or how differential expression leads to pathogenesis. Single-cell and single-nuclei RNA sequencing (scnRNAseq) provide particularly valuable insights into the role of gene expression in pathology by allowing us to examine gene expression changes at the level of individual cells within disease-relevant tissues. To date, this high-resolution technology has proven opportunistic for revealing cell types vulnerable to degeneration in PD and has informed the contribution of specific cell types to the disease process (13–15). Additionally, scnRNAseq has the potential to identify intra-cell type, differentially expressed genes (DEGs) between diseased and healthy cells, nominating genes for targeted genomic analyses to reveal novel associations that may have been overlooked in broader exome-or genome-wide analyses.

Despite this prospect, the robust identification of DEGs in scnRNAseq is challenging. First, the costs of scnRNAseq remain prohibitively high for analyzing entire patient cohorts, which is critical to account for the heterogeneity of PD and ensure robust DGE signals (16). Pan-dataset analyses could, in theory, mitigate this limitation, but the nuances of scnRNAseq technologies — variations in library preparation protocols, technical artifacts, and dropout — can introduce inconsistencies across datasets (17); thus, a pan-dataset, intersectional approach for identifying DEGs may be overly stringent. Second, benchmarking analyses have yielded conflicting conclusions regarding the best statistical methods for identifying DEGs from scnRNAseq data, and the choice of statistical method can provide vastly different results (17–19). One perspective advocates for treating each cell as a distinct replicate and utilizing methods tailored to scnRNAseq to compute DGE (20–22). However, this approach can lead to statistical inflation due to exceedingly large sample sizes. An alternative strategy, known as a pseudo-bulk approach, aggregates the counts from all cells belonging to each individual subject prior to computing DGE with statistical methods originally designed for bulk RNAseq analyses (23–25). While this method reduces the risk of inflated significance, it may overlook important biological signals due to the limited statistical power associated with treating each subject as a distinct replicate.

The limitations associated with standard DGE methods, coupled with advancements in machine learning (ML) raises the question of whether these state-of-the-art algorithms can identify robust changes in gene expression at a single-cell resolution. ML encompasses computational methods that allow computers to learn from data and make decisions without explicit programming. Unsupervised ML, which identifies underlying structures in unlabeled data, is standard in scnRNAseq analyses, encompassing processes such as dimensionality reduction and clustering (26). In contrast, supervised ML methods are trained on labeled data to learn the relationship between input features and the corresponding target value or class label, and are commonly used for cell type classification (27).

An exciting additional application of supervised machine learning is classifying transcriptomic samples based on disease status, a technique that has been previously applied to bulk RNA sequencing data (28). In the context of scnRNAseq, if ML classifiers can be trained to accurately distinguish between the transcriptomes of diseased and healthy cells, this would open avenues for deeper investigation into the underlying “reasons” for a particular cell being classified as diseased. However, many ML classifiers operate as “black boxes”, making it difficult to interpret their decision-making processes (29). To address this, interpretable explainers like Local Interpretable Model-agnostic Explanations (LIME) can be applied to approximate the behavior of the complex model and reveal the most influential features driving a classification decision (30). For scnRNAseq data, these influential features would constitute the expression profiles of specific genes. We reasoned that these LIME-identified features could provide valuable insights into the molecular characteristics of PD cells and nominate genes for targeted genomic analyses aimed at uncovering novel genetic associations.

In this work, we introduce an interpretable, ML classifier-based framework for identifying gene expression changes between diseased and healthy cells. We begin by evaluating the prospect of training ML classifiers to distinguish between PD and healthy cells and outline the best methods for this task. We then apply our novel approach to three distinct single-nuclei RNA sequencing (snRNAseq) datasets characterizing the post-mortem midbrain of individuals with PD and healthy controls to identify the most important genes for classifying eight distinct, broad cell types from the PD midbrain (13–15). Finally, we employ a reductionist approach to pinpoint the most biologically informative molecular features characterizing diseased cells and corroborate the role of these genes in PD using multiple sources of genetic data.

## Results

### Deep neural networks best classify single nuclei transcriptomes by disease status

To determine which ML models are best suited to classify single-nuclei transcriptomes according to disease status, we used a snRNAseq dataset characterizing seven broad cell types from the post-mortem substantia nigra (SN) of individuals with PD (*n =* 6) and healthy controls (*n* = 8) prepared by Kamath et al. (13) (**Figure 1A**). We tested the combination of four feature selection methods — highly variable genes (HVG), principal component analysis (PCA), non-negative matrix factorization (NMF), and embedded topic modelling (ETM) (31) — with four types of ML models — neural network (NN), logistic regression, random forest, and support vector machine (**Figure 1A**). For each combination of feature selection method and ML model (*n* = 16), we trained cell type-specific classifiers on 80% of the data and evaluated the model’s performance on the remaining 20% of the unseen data (see Methods). A cell was considered correctly classified if the disease status predicted by the ML classifiers matched the disease status of the individual from which the cell derived.

**Figure 1.**
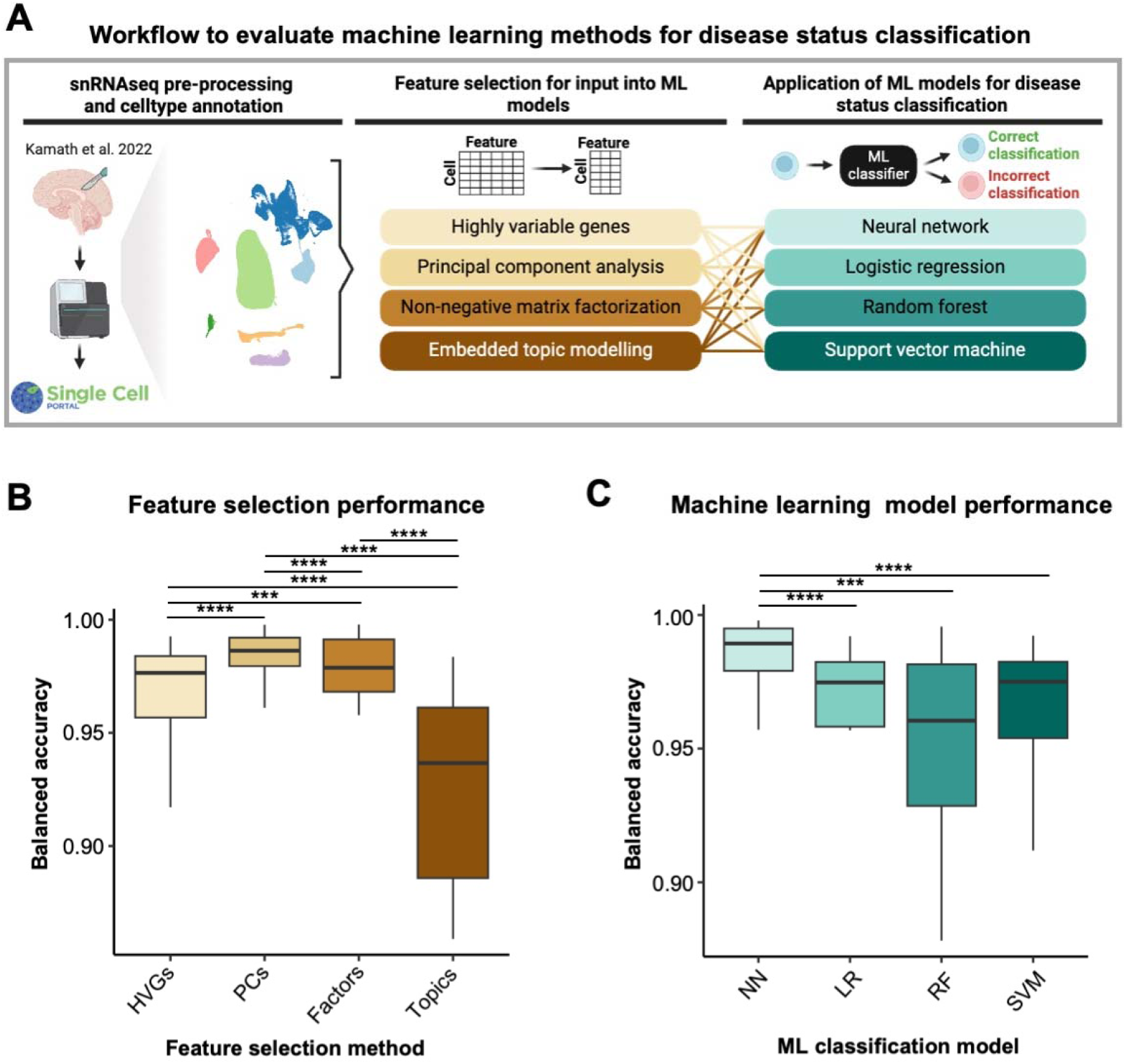
Evaluating different methods for classifying single-nuclei transcriptomes by disease status. **A)** Schematic of the analysis workflow. For this analysis, we used publicly available single-nuclei RNA sequencing (snRNAseq) data from the post-mortem substantia nigra of individuals with Parkinson’s disease and healthy controls prepared by Kamath et al. Four feature selection methods were independently applied to the snRNAseq data before being input into four distinct machine learning (ML) models for disease status classification of individual cells. A total of 16 unique combinations of feature selection methods and ML models were tested, with each combination applied to classify seven distinct cell types independently and all cell types together. For each feature selection-ML model combination, we trained five independent models per cell type using five-fold cross validation (*n* = 128 models). **B)** Boxplot showing the disease status classification accuracy obtained using different feature selection methods. **C)** Boxplot showing the disease status classification accuracy obtained using different ML models. A Wilcoxon rank-sum test was used to compare mean balanced accuracies; non-significant comparisons are not shown. Abbreviations: NN, neural network; HVG, highly variable gene; LR, logistic regression; PC, principal component; RF, random forest; SVM, support vector machine. *** P < 0.001; **** P < 0.0001.

The balanced accuracies of the cell type-specific ML classifiers when leveraging different feature selection methods for input are shown in **Figure S1**. On average, the principal components (PCs) obtained from PCA showed the highest disease status classification accuracy across all ML models and cell types (mean balanced accuracy = 0.984), followed by the components obtained from NMF (mean balanced accuracy = 0.975), HVGs (mean balanced accuracy = 0.965), and topics obtained from ETM (mean balanced accuracy = 0.921) (**Figure 1B)**. In turn, the NN classifiers showed the highest disease status classification balanced accuracy, on average, across all feature selection methods and cell types (mean balanced accuracy = 0.984), followed by random forest (mean balanced accuracy = 0.955), logistic regression (mean balanced accuracy = 0.954), and support vector machine (mean balanced accuracy = 0.952) (**Figure 1C**). Based on these results, we elected to leverage HVGs — genes that show variation across all cells, independent of disease status — for feature selection and NNs for disease status classification in subsequent analyses. The HVG feature selection method was selected because, while its accuracy was very slightly below that of the PC method, it facilitated high ML classification accuracy and will provide directly interpretable results upon decoding the classifiers to elucidate the most important features characterizing PD cells. NN was selected due to its superior disease status classification performance compared to the three remaining ML classifiers.

### Neural networks accurately classify midbrain transcriptomes from Parkinson’s disease and healthy controls

Hereafter, we trained NN classifiers using HVGs as input to distinguish between PD and healthy cells with the ultimate goal of decoding the models to reveal the most important genes for classifying diseased cells. To this end, we leveraged three separate snRNAseq datasets characterizing the post-mortem midbrain of individuals with PD and healthy controls prepared by i) Kamath et al. (13) (**Figures 2Ai-iii**); ii) Wang et al. (14) (**Figures 2Bi-iii**); and iii) Smajic et al. (15) (**Figures S2A-C)**. A summary of these datasets is presented in **Table 1**. We selected the Kamath et al. and Wang et al. datasets for exploratory analysis, where we trained cell type- and dataset-specific NN classifiers and identified important genes for classifying PD cells. The Smajic et al. dataset was reserved as a validation set to evaluate the generalizability of the genes identified in the exploratory phase.

**Figure 2.**
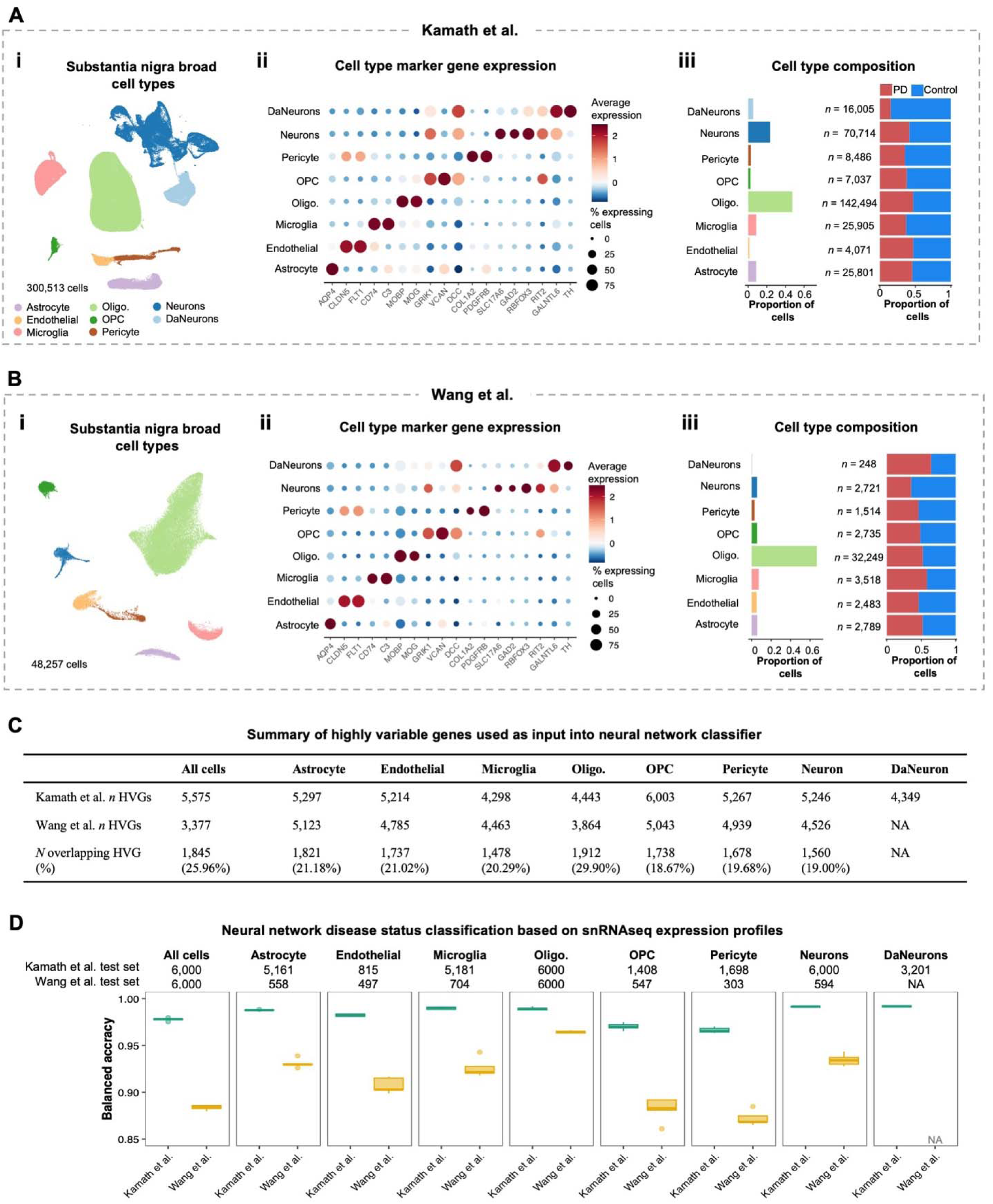
Application of neural networks to classify single-nuclei transcriptomes from the substantia nigra by disease status. **A,B)** Characterization of the single-nuclei RNA sequencing (snRNAseq) datasets prepared by Kamath et al. and Wang et al., respectively. **i)** Uniform manifold approximation and projection (UMAP) showing the annotated cell types comprising the substantia nigra. **ii)** Dot plot showing the expression of literature-curated cell type marker genes. **iii)** Bar plots showing the proportion of cells annotated to each cell type (left) and the distribution of cells from individuals with Parkinson’s disease (PD) and healthy controls (right). The total number of cells per cell type is shown. **C)** Summary table of highly variable gene (HVG) counts identified for each cell type from the Kamath et al. and Wang et al. datasets. These HVGs were used as input to the neural networks (NN) for disease status classification. HVGs were not computed for dopaminergic neurons (DaNeurons) from Wang et al. due to insufficient cells for the machine learning (ML) test-train split workflow. **D)** Boxplots showing the disease status classification balanced accuracy of the NN models. Independent NN classifiers were trained and evaluated for unique cell types from each dataset. Five-fold cross-validation on the training set resulted in five distinct models that were each independently applied to the test set for model evaluation. The number of cells in the test set reserved for NN evaluation is indicated at the top of the panels. DaNeurons from Wang et al. were excluded due to insufficient cells for the ML test-train split workflow. Abbreviations: oligo, oligodendrocytes; OPC, oligodendrocyte precursor cells.

**Table 1.** Summary of single-nuclei RNA sequencing datasets characterizing Parkinson’s disease and healthy control midbrains.

| Author | *N* subjects |  | *N* cells |  | Genes<br>per cell<br>(median) | UMI per<br>cell<br>(median) | %<br>mitochondrial<br>genes (mean) | %<br>ribosomal<br>proteins<br>(mean) |
| --- | --- | --- | --- | --- | --- | --- | --- | --- |
|  | PD | Control | PD | Control |  |  |  |  |
| Kamath<br>et al.<br>2022 | 6 | 8 | 127,647 | 172,866 | 3,259 | 7,442 | 2.33 | 0.89 |
| Wang et<br>al. 2024 | 5 | 6 | 21,890 | 26,367 | 1,211 | 2,065 | 5.40 | 0.61 |
| Smajic et<br>al. 2022 | 6 | 5 | 17,995 | 22,289 | 2,453 | 5,279 | 1.49 | 0.91 |

Abbreviations: PD, Parkinson’s disease; UMI, unique molecular identifier.

We annotated each dataset to eight distinct, broad cell types based on published cell type annotations and the expression of literature-curated marker genes: astrocytes (*AQP4*), endothelial cells (*CLDN5*, *FLT1*), microglia (*CD74*, *C3*), oligodendrocytes (*MOBP*, *MOG*), oligodendrocyte precursor cells (OPCs; *GRIK1*, *VCAN*, *DCC*), pericytes (*COL1A2*, *PDGFRB*), neurons (*SLC17A6*, *GAD2*, *RBFOX3*, *RIT2*), and DaNeurons (*GALNTL6*, *TH*) (**Figures 2Aii, 2Bii, and S2B**). Evaluating the transcriptional similarity of cell types across datasets using *MetaNeighbor* (32) revealed high inter cell-type replicability (mean area under the receiver operating characteristic curve (AUROC) = 0.987), supporting our pan-dataset analytical approach (**Figure S3**). **Figures 2Aiii, 2Biii, and S2C** show the number of cells annotated to each cell type and their distribution in PD and healthy controls from each dataset. Importantly, we annotated too few DaNeurons for the ML classifier workflow in the Wang et al. and Smajic et al. datasets (*n* = 248 and 51, respectively), which can be attributed to the relative sparsity of this cell type and the vulnerability of neuronal cells to the tissue dissociation process, resulting in their underrepresentation in cell suspensions prior to sequencing (33, 34). Conversely, Kamath et al. enriched their dataset for neurons using fluorescence activated cell sorting (FACS) prior to sequencing, affording 16,005 cells annotated as DaNeurons; thus, subsequent analyses of DaNeurons were restricted to the Kamath et al. dataset.

To reduce the number of features input into the NN classifiers, we identified cell type- and dataset-specific HVGs from each exploratory dataset (see Methods). The average number of cell-type specific HVGs was 5,015 and 4,678 for Kamath et al. and Wang et al., respectively (**Figure 2C**). Additionally, we identified HVGs across all cell types combined (Kamath et al. *n* HVGs = 5,575; Wang et al. *n* HVGs = 3,377) to train pan-cell type NN classifiers. Taking the HVGs as input, we trained separate NN classifiers for each cell type in each exploratory dataset, as well as two dataset-specific models for classifying all cell types together, resulting in a total of 17 distinct models. The average disease status classification balanced accuracy across all models from Kamath et al. and Wang et al. was 0.983 and 0.913, respectively (**Figure 2D**). Additional analyses suggested that the discrepancy in the classification accuracy between the exploratory datasets can be attributed to both a higher number of available cells and increased sequencing depth in the Kamath et al. dataset compared to the Wang et al. dataset (**Table 1; Figures S4-S6**).

Given the remarkable capacity of machine learning models to discern complex patterns for accurate classification (35), we investigated whether our NN classifiers were truly capturing PD-associated gene expression changes or merely responding to random variations in the data. To address this, we randomized the disease status labels of a proportion of cells and retrained the NN classifiers to assess how randomization affected classification accuracy. Across the cell type- and dataset-specific NN classifiers, the mean balanced accuracy was 0.948 with 0% randomization; 0.789 with 25% randomization; 0.645 with 50% randomization; 0.537 with 75% randomization; and 0.496 with 100% randomization, the latter being equivalent to chance performance (**Figure S7**). These results suggest that the NN classifiers were effectively identifying PD-associated changes in gene expression, supporting our approach to leverage these models to identify important genes characterizing PD cells in subsequent analyses.

### LIME identifies transcriptional markers for Parkinson’s disease cells consistent across datasets

Despite the capacity of the NN models to stratify diseased and heathy cells, the underlying reasons for their classification decision remained unresolved. To address this, we applied LIME to decode the “black box” of our NN classifiers and reveal the most important genes influencing the classification decision for an individual cell (30). In brief, LIME approximates the behavior of the complex model with a simpler, interpretable model around each prediction. It perturbs the input data, observes the changes in the predictions, and uses this information to build a local linear model resulting in a feature importance score for each gene, reflecting the gene’s impact on the disease status classification decision (30). To approximate a global explanation — that is, gene-specific feature importance values across all cells — we first aggregated the LIME feature importance values across all cells belonging to a single subject and then across all subjects (see Methods).

To assess the pan-dataset generalizability of LIME feature importance values, we first calculated Pearson’s correlation between the LIME feature importance Z-scores across the two exploratory datasets. We observed moderately strong correlations within each cell type, indicating pan-dataset consistency in key genes for classifying PD cells (**Figure 3A**). To further interrogate the pan-dataset generalizability of LIME, we compared this method to MAST (36) and DESeq2 (37), two well-established DGE methods. We computed DEGs (adjusted p-value < 0.05; absolute log2 fold-change (Log2FC) > 1.00) using both methods separately for each exploratory dataset and determined the proportion of cell type-specific DEGs identified in both datasets (pan-dataset genes). Similarly for LIME, we retained genes with a cell type-specific LIME feature importance Z-score > 1 from each dataset and computed the proportion of pan-dataset genes. The average proportion of cell type-specific pan-dataset genes was 3.88% for MAST, 0.63% for DESeq2, and 10.25% for LIME. A Wilcoxon rank-sum test showed that LIME identified a significantly higher proportion of pan-dataset genes compared to both MAST (p-value = 1.2e-3) and DESeq2 (p-value = 2.1e-3) (**Figure S8**). Operating under the assumption that pan-dataset genes are more robust and more likely to result from true biological signal, these results suggest that LIME is highly suitable to identify gene expression changes between diseased and healthy cells.

**Figure 3.**
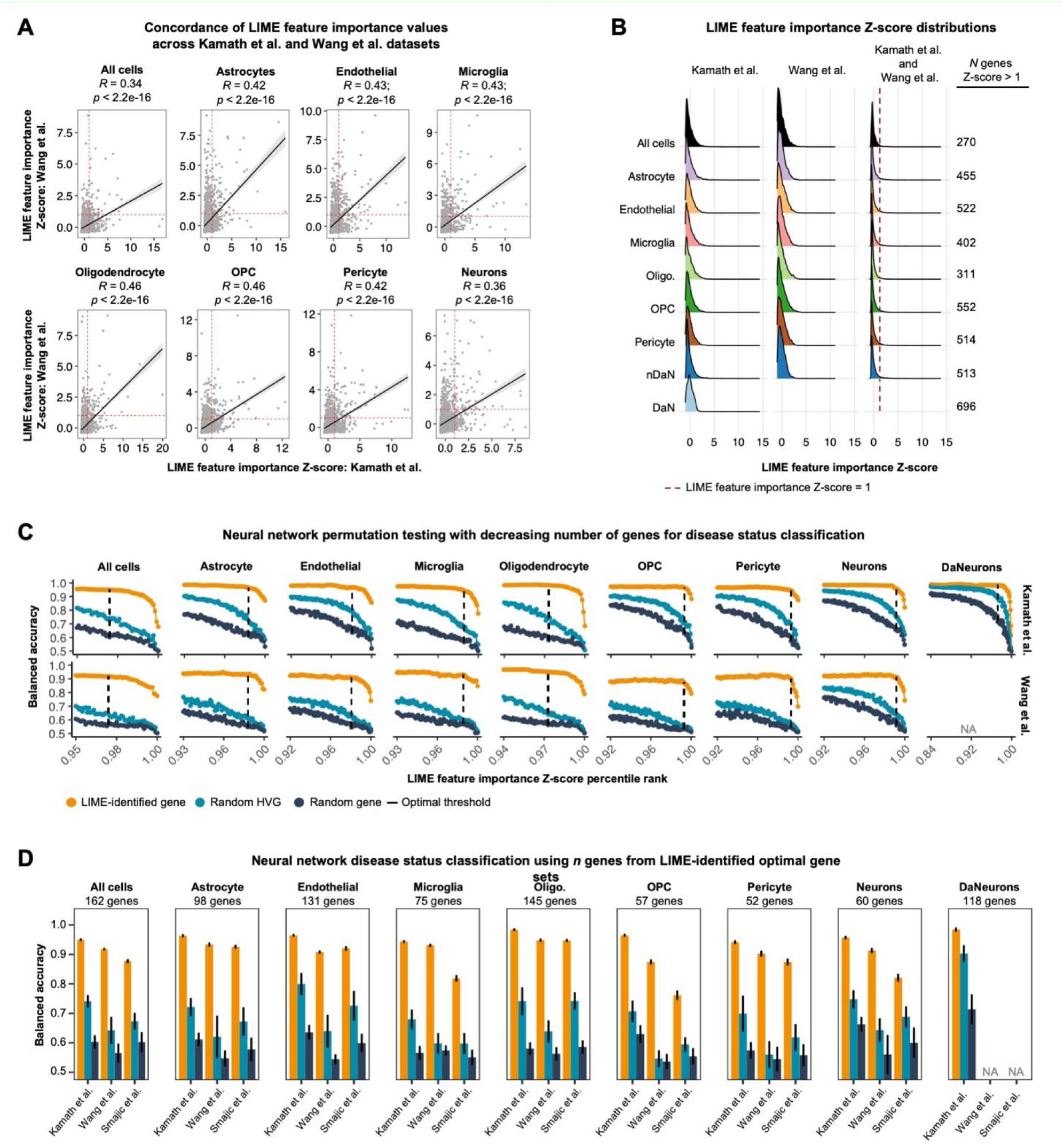
Application of LIME to identify genes influencing the classification decision by neural networks. **A)** Scatter plots showing the cell type-specific correlations between the LIME feature importance Z-scores in Kamath et al. and Wang et al. Pearson’s correlation coefficient (R) and the corresponding p-value are shown at the top of the panels. **B)** Ridge plots showing the LIME feature importance Z-score distributions across cell types in Kamath et al. (left), Wang et al. (middle), and the mean Z-score across both datasets (right). **C)** Dot plots showing the neural network (NN) disease status classification balanced accuracy obtained from permutation tests with decreasing feature counts used as input. HVGs with a mean LIME feature importance Z-score > 1 were incrementally eliminated based on their Z-score percentile rank until only the most important features remained. The same permutation tests were performed with an equal number of randomly selected HVGs (turquoise) and randomly selected genes (dark blue) as a benchmark to compare the classification accuracy of NNs using LIME-identified genes (orange). The dashed LIME indicates the optimal threshold: the number of input genes that maximized the discrepancy in classification accuracy between using LIME-identified genes and random genes, across both Kamath et al. (top) and Wang et al. (bottom). **D)** Bar plots showing the NN disease status classification balanced accuracy when using the cell type-specific LIME optimal gene sets as input for three separate datasets (orange). The number of genes in the LIME optimal gene sets are shown at the top of the panels. For comparison, NNs were trained and evaluated using an equal number of randomly selected HVGs (turquoise) and randomly selected genes (dark blue). Error bars represent the standard deviation of the balanced accuracy across ten independent models; a distinct set of random HVGs and random genes were used for each model. Abbreviations: DaNeurons, dopaminergic neurons; oligo, oligodendrocytes; OPC, oligodendrocyte precursor cells.

Having substantiated LIME’s capacity to identify generalizable genes, we next employed a multi-step approach to pinpoint the most biologically-relevant genes for characterizing PD cells. We first aimed to identify genes that were consistently important for disease status classification by computing the average, cell type-specific LIME feature importance Z-score across both exploratory datasets for each HVG input to the NN classifiers (**Figure 3B**). Filtering the HVGs to only retain those with a mean Z-score > 1 resulted in cell type-specific lists of the most influential transcriptional markers in PD cells, which averaged 471 genes per cell type (**Figure 3B**).

Next, we aimed to identify the number of cell type-specific genes with a mean LIME feature importance Z-score > 1 that were essential for accurate disease status classification, allowing us to exclude features that did not enhance NN performance. To this end, we conducted permutation tests using progressively fewer LIME-identified genes as input to the cell type- and dataset-specific NN classifiers and evaluated the model’s performance. Throughout the permutations, genes were incrementally eliminated on the basis of their mean LIME feature importance Z-score until only the most important features remained (see Methods). Additionally, we performed the same permutation tests with both randomly selected HVGs and randomly selected genes as a benchmark to compare the classification accuracy of NNs using LIME-identified genes. Through this exercise, we found that NN classifiers using LIME-identified genes outperformed those using randomly selected features, with the discrepancy in performance increasing as the number of input genes decreased (**Figure 3C**). Notably, this trend was consistent across both exploratory datasets. Given that the same cell type-specific genes were input to the dataset-specific classifiers for each permutation, these results suggest that LIME effectively identified features that were consistently important for disease status classification across datasets. To identify the cell type-specific, optimal gene sets for disease status classification, we determined the number of genes that maximized the performance gap between the NN classifiers using LIME-identified genes and those using random genes across both exploratory datasets (indicated by a dashed line in **Figure 3C**) (see Methods). The genes comprising the cell type-specific LIME optimal gene sets are shown in **Table S1** (all cells = 162 genes; astrocytes = 98 genes; endothelial cells = 131 genes; microglia = 75 genes; oligodendrocytes = 145 genes; OPCs = 57 genes; neurons = 60 genes; DaNeurons = 118 genes).

Next, we sought to establish whether the cell type-specific, LIME optimal gene sets obtained from the exploratory datasets could accurately classify cells from the Smajic et al. validation dataset. We re-trained cell type- and dataset-specific NN classifiers using the genes from the LIME optimal gene sets, alongside an equivalent number of randomly selected HVGs and random genes. In Kamath et al., the average balanced accuracy of the NN classifiers across all distinct cell types was 0.962 when using the LIME optimal gene set as input, 0.749 when using randomly selected HVGs as input, and 0.620 when using randomly selected genes as input; in Wang et al., the average balanced accuracy of the classifiers was 0.916 when using the LIME optimal gene set, 0.611 when using randomly selected HVGs, and 0.554 when using randomly selected genes; in the Smajic et al. validation dataset, the average balanced accuracy of the classifiers was 0.868 when using the LIME optimal gene set, 0.664 when using randomly selected HVGs as input, and 0.578 when using randomly selected genes (**Figure 3D**). Indeed, the cell type-specific NN classifiers using the LIME optimal gene sets showed higher performance than the classifiers using randomly selected features of equal sizes in all three datasets. Interestingly, the classification accuracy using randomly selected genes was highest for DaNeurons compared to the other cell types, which may be due to high transcriptional dissimilarity between diseased and healthy DaNeurons (**Figure 3D**). Taken together, this analysis demonstrated that the cell type-specific LIME optimal gene sets can effectively classify PD cells across three separate datasets, highlighting the robustness of the findings and the ability of our LIME-based approach to identify generalizable molecular markers.

### LIME-identified genes are biologically relevant to Parkinson’s disease pathology

While the LIME-identified genes demonstrated a remarkable ability to distinguish PD cells from their healthy counterparts across three independent datasets, it remained unclear whether these genes were capturing true disease-associated biological perturbations. To address this, we subjected the cell type-specific LIME optimal gene sets to gene set overrepresentation analyses to evaluate their functional relevance and determine whether they were enriched for known biological processes in PD. Across all non-neuronal cell types, we observed an enrichment of processes related to the heat shock and unfolded protein response, consistent with its well-established role in PD (38) (**Figures S9-17**). Additionally, we observed that the LIME optimal gene sets for microglia, oligodendrocytes, and OPCs were enriched for terms related to antigen presentation, corroborating the evidence for the immune response in PD pathophysiology (39) (**Figures S11-13**). Astrocytes were uniquely enriched for processes related to copper ion dysregulation, which has been linked to aberrant dopamine metabolism, oxidative stress, and α-synuclein aggregation — the major protein component of Lewy bodies (40) (**Figures S9**). Interestingly, the LIME optimal gene set for neurons was enriched for genes involved in aggrephagy — the selective degradation of misfolded or aggregated proteins (41) (**Figures S15**). In contrast, the LIME optimal gene set for DaNeurons was enriched for terms suggesting synaptic dysfunction, which has been linked to the loss of this vulnerable cell type in PD (42) (**Figures S16**). These results illustrate that the LIME-identified genes are not only valuable for *in silico* disease status classification by the NN models but also reflect relevant biological processes in PD.

### A multi-step filtering process pinpoints the most biologically informative genes for disease status classification

Nominating genes for targeted genomic analysis requires a careful balance: retaining enough genes to capture biologically relevant signals while eliminating noise that can undermine analytical power due to the burden of multiple hypothesis testing. Accordingly, we aimed to further refine the cell-type specific LIME optimal gene sets and pinpoint prospective genetic determinants in PD by employing a three-step filtering criteria. Here, we only focused on genes that we deemed to be biologically informative, excluding mitochondrially encoded genes, ribosomal proteins, sex-linked genes, and non-coding RNAs. In **Figure 4**, we leverage astrocytes to illustrate this filtering process.

**Figure 4.**
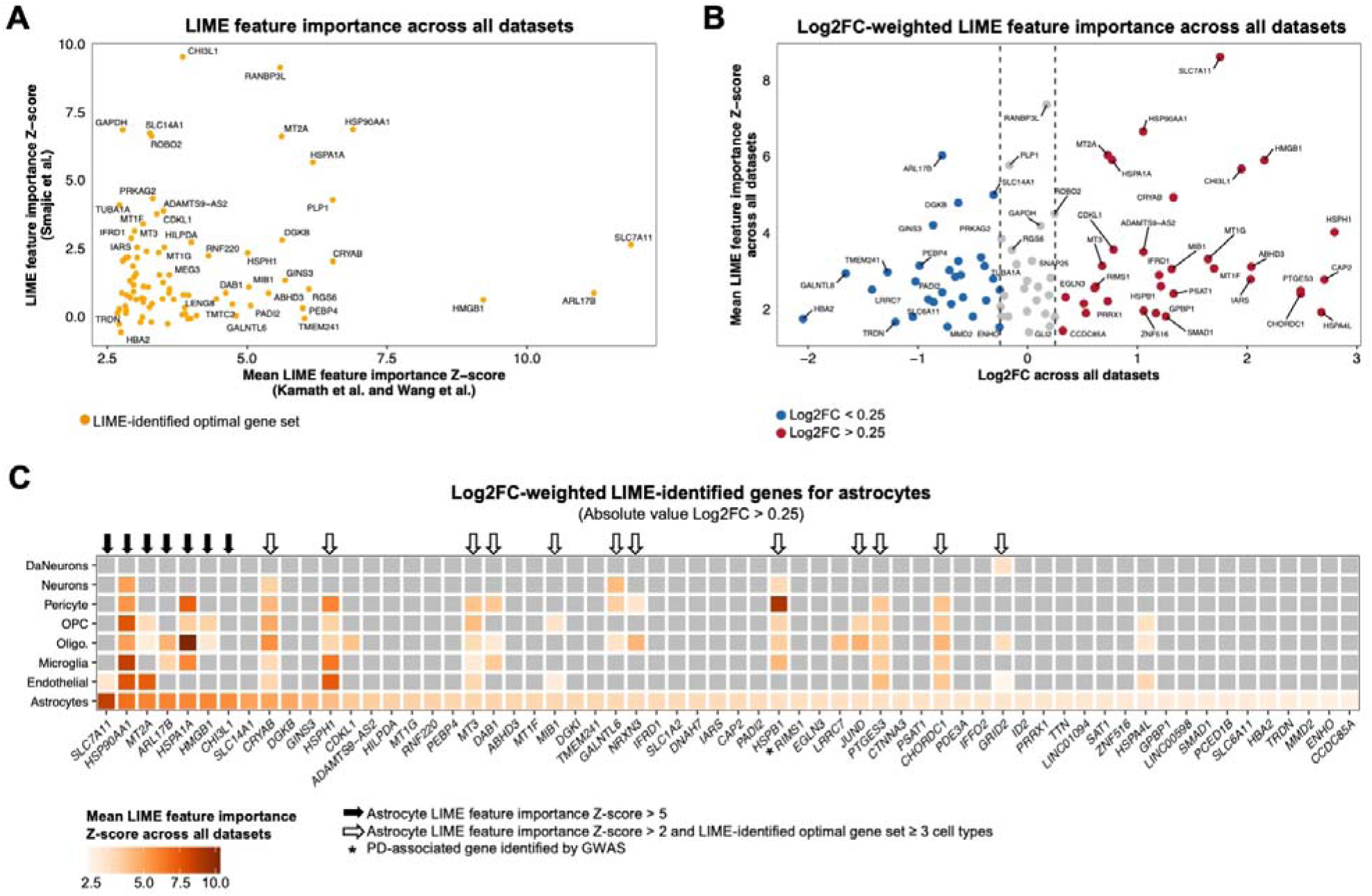
Prioritization of LIME-identified genes for characterizing Parkinson’s disease cells. This figure uses astrocytes as a case study to demonstrate our approach for identifying the most biologically relevant genes identified by LIME. **A)** Scatter plot showing the LIME feature importance Z-score for genes in the LIME optimal gene set for astrocytes. **B)** Volcano plot showing the mean LIME feature importance Z-score across the Kamath et al., Wang et al., and Smajic et al. datasets and the gene expression log2 fold-change (Log2FC) between Parkinson’s disease (PD) and control astrocytes across all datasets. Only genes from the LIME-identified optimal gene set for astrocytes are included. **C)** Heatmap showing the mean LIME feature importance Z-scores for genes in the astrocyte-specific optimal gene set with an absolute Log2FC > 0.25. Grey tiles indicate genes that were not included in the cell type-specific optimal gene set. The genes retained for subsequent analysis are labelled. Abbreviations: oligo, oligodendrocytes; OPC, oligodendrocyte precursor cells.

Prior to filtering, we directly investigated the feature importance values of the LIME-identified genes obtained from the exploratory datasets (Kamath et al. and Wang et al.) in the Smajic et al. validation dataset. For astrocytes, this exercise revealed the consistent importance of select genes for classifying PD cells across all three datasets, including *HSP90AA1*, *HSPA1A*, *MT2A*, *PLP1*, and *CRYAB*, all of which have been previously implicated in PD (43–47), corroborating the ability of our LIME-based framework to identify replicable, disease-relevant signals (**Figure 4A**). Next, we integrated all three snRNAseq datasets (Kamath et al., Wang et al., and Smajic et al.) and computed the cell type-specific Log2FC in gene expression between PD and control cells. We then only retained biologically informative genes from the LIME optimal gene sets with an absolute value Log2FC > 0.25 (criteria 1). For astrocytes, this initial criterion reduced the number of genes from 98 to 62 (**Figure 4B**). Amongst the LIME-identified genes with an absolute value Log2FC > 0.25, we retained genes with a LIME feature importance Z-score > 5 in a particular cell type (criteria 2) or genes with a Z-score > 2 in a particular cell type but identified by LIME in ≥ 3 cell types (criteria 3). For astrocytes, 7 genes — *SLC7A11*, *HSP90AA1*, *MT2A*, *ARL17B*, *HSPA1A*, *HMGB1*, and *CHI3L1* — had a LIME Z-score > 5, indicating high importance for characterizing PD astrocytes. In turn, 12 genes — *CRYAB*, *HSPH1*, *MT3*, *DAB1*, *MIB1*, *GALNTL6*, *NRXN3*, *HSPB1*, *JUND*, *PTGES3*, *CHORDC1*, and *GRID2* — had a Z-score > 2 in astrocytes and were identified in ≥ 3 cell types by LIME, reflecting their broader importance for distinguishing between PD and healthy cells across multiple cell types (**Figure 4C**). We also retained LIME-identified genes previously associated with PD through genome-wide association studies (GWAS).

This same three-step filtering process was applied to the LIME optimal gene sets for each cell type (**Figure S18-24**). After filtering each cell type-specific gene set, we retained 66 unique genes passing our filtering criteria: 41 genes showed high LIME feature importance values in a single cell type (criteria 2) and 25 genes were identified by LIME in ≥ 3 cell types (criteria 3) (**Tables S2 and S3**). These 66 unique genes constituted the final set of LIME-identified genes and were the focus of our subsequent analyses.

### LIME-identified genes show varying levels of support from different modalities for their implication in Parkinson’s disease

The 66 unique, LIME-identified genes are shown in **Figure 5**. Of the 25 genes that LIME designated as important for characterizing PD in ≥ 3 cell types, 19 were identified uniquely in non-neuronal cell types. Unsurprisingly, neurons and DaNeurons exhibited the lowest overlap of LIME-identified genes with other cell types. Many of the genes identified by LIME in ≥ 3 cell types were heat shock proteins, including *HSP90AA1*, *HSPB1*, *HSPH1*, *HSPA1A*, *HSP90AB1*, *HSPA1B*, *HSPA4L*, and *DNAJB1*, which function as molecular chaperones aiding in protein stabilization against stress-induced aggregation (48). Additional genes identified in ≥ 3 cell types that are known to interact with heat shock proteins to confer proteostasis included *CRYAB*, *CHORDC1*, *PTGES3*, *STIP1*, and *CACYBP* (**Figure S25**). Importantly, all LIME-identified genes related to proteostasis showed increased expression in non-neuronal cells from individuals with PD compared to healthy controls across the three snRNAseq datasets (**Figure 5**), possibly reflecting the accumulation of α-synuclein aggregates in the Parkinsonian brain. *MT3, MT2A, MIB1,* and *FTL* were also identified by LIME in ≥ 3 non-neuronal cell types and showed increased expression in PD cells compared to healthy cells. On the other hand, *DAB1*, *GALNTL6*, *ARL17B*, *JUND*, *NRXN3*, and *B2M* were identified by LIME in ≥ 3 non-neuronal cell types and showed decreased expression in PD. The intersection of LIME-identified features across cell types indicates that genetic factors, environmental influences, or a combination of both may elicit a partially similar pathophysiological response in distinct cell types in PD.

**Figure 5.**
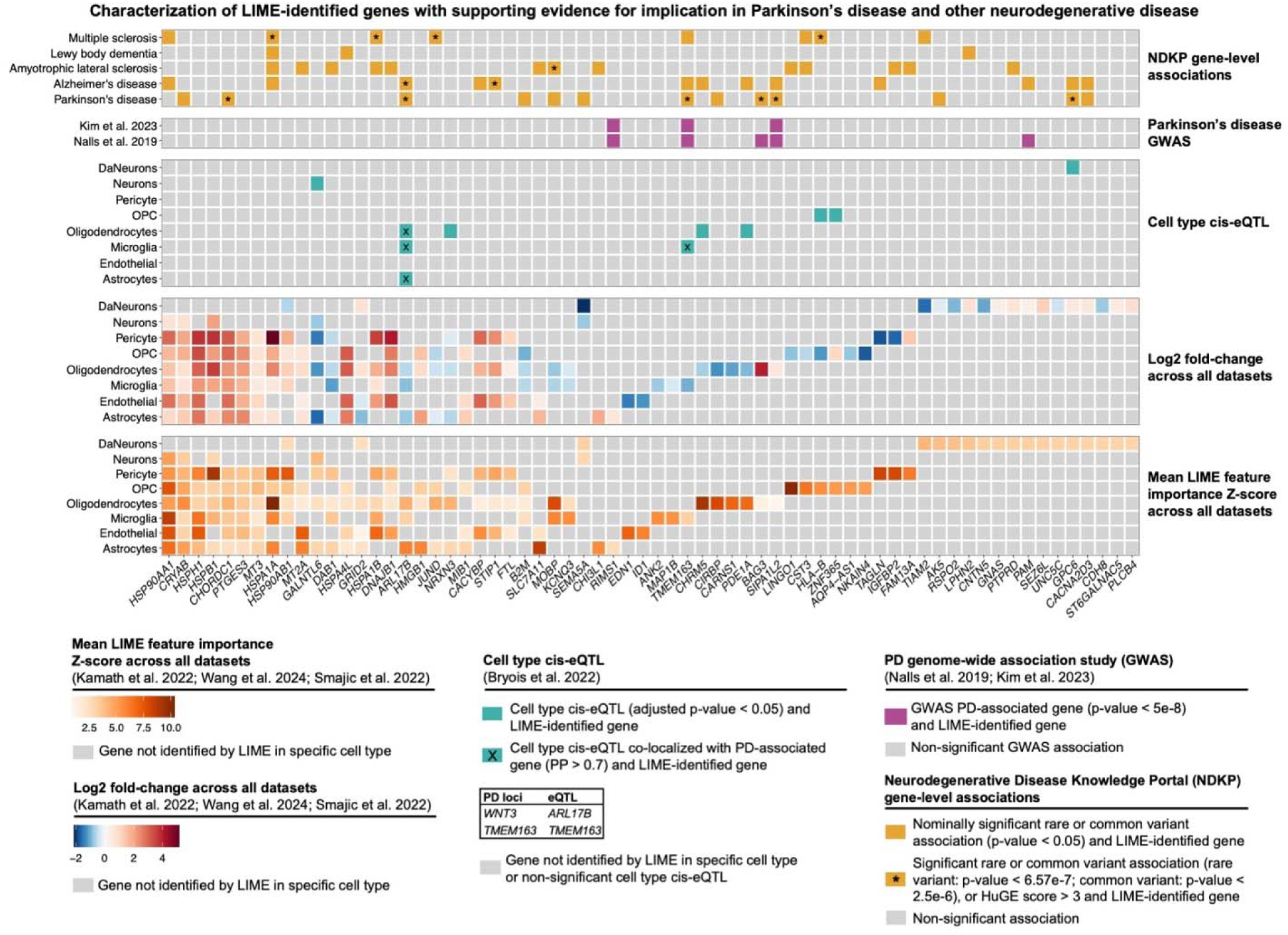
Characterization of filtered LIME-identified genes. Heatmap characterizing the 66 unique genes included in the final LIME-identified gene set. The panels are arranged from bottom to top as follows: 1) Mean LIME feature importance Z-Scores: This panel shows the mean LIME feature importance Z-scores across the Kamath et al., Wang et al., and Smajic et al. datasets. Grey tiles indicate genes that were not identified by LIME in the corresponding cell type. 2) Log2 fold-change: this panel shows the log2 fold-change between Parkinson’s disease (PD) and control cells across all snRNAseq datasets. Grey tiles indicate genes that were not identified by LIME in the corresponding cell type. 3) Cell type cis-expression quantitative trait loci (eQTL): this panel highlights LIME-identified genes that were identified as cell type cis-eQTLs by Bryois et al. Tiles marked with an “X” represent cell type cis-eQTLs that co-localized with PD-associated genes (posterior probability (PP) > 0.7). Grey tiles indicate genes that were not identified by LIME or were not identified as cis-eQTLs in the corresponding cell type. 4) PD genome-wide association analysis (GWAS): this panel highlights LIME-identified genes that showed an association with PD-risk by GWAS performed by Nalls et al. or Kim et al. Only genes that met genome wide significance (p-value < 5e-8) are shown. Grey tiles indicate genes that were not found to be significantly associated with PD in Nalls et al. (bottom) or Kim et al. (top). 5) Neurodegenerative Disease Knowledge Portal (NDKP) gene-level associations: this panel highlights LIME-identified genes that showed gene-level associations with various diseases in the NDKP cohorts. All nominally significant common and rare variant associations are shown. Tiles marked with an asterisk denote significant associations with the corresponding disease or a Human Genetic Evidence (HuGE) Score ≥ 3. Grey tiles indicate non-significant associations. Abbreviations: DaNeurons, dopaminergic neurons; OPC, oligodendrocyte precursor cells.

To further characterize the LIME-identified genes, we performed a comprehensive review of publicly available data for evidence supporting their implication in PD and other neurodegenerative diseases. First, we explored cell type cis-expression quantitative trait loci (eQTL) identified by Bryois et al. (49), which investigated how genetic variants influence the expression of proximal genes across eight distinct cell types from post-mortem brain tissue. Nine cell type cis-eQTLs identified by Bryois et al. were identified by LIME in the corresponding cell type: *GALNTL6* (neurons), *ARL17B* (astrocytes, microglia, and oligodendrocytes), *NRXN3* (oligodendrocytes), *TMEM163* (microglia), *CHRM5* (oligodendrocytes), *PDE1A* (oligodendrocytes), *HLA-B* (OPC), *ZNF365* (OPC), and *GPC6* (DaNeurons) (**Figure 5; Table S4**). Amongst these cis-eQTLs, *ARL17B* and *TMEM163* co-localized with genetic variants associated with PD risk at a posterior probability (PP) > 0.7 (49) (**Table S5**). *ARL17B* co-localized with *WNT3* in astrocytes, endothelial cells, microglia, oligodendrocytes, OPCs, excitatory neurons, and inhibitory neurons, and was identified by LIME in astrocytes, microglia, and oligodendrocytes. In turn, *TMEM163* co-localized exclusively in microglia and was identified by LIME only in this cell type. The overlap between the LIME-identified genes and the cell type-specific cis-eQTLs identified by Bryois et al. highlights genes that merit further investigation to elucidate how genetic variants may influence cell type-specific gene expression and how this altered expression could contribute to PD pathology.

Next, we utilized PD GWAS summary statistics from Nalls et al. (8) and Kim et al. (9) to highlight the LIME-identified genes that have been previously associated with PD risk (**Table S6**). Interestingly, all five PD-associated genes captured by LIME were uniquely identified in a single cell type (**Figure 5**). *RIMS1* (Nalls et al.: p-value = 2.00e-10, beta = 0.0657; Kim et al.: p-value = 7.00e-12, beta = 0.0208) was identified by LIME in astrocytes and showed decreased expression in PD. *TMEM163* (Nalls et al.: p-value = 5.00e-14, beta = 0.0807; Kim et al.: p-value = 5.00e-12, beta = 0.0400) was identified by LIME in microglia and showed decreased expression in PD. *BAG3* (Nalls et al.: p-value = 2.00e-11, beta = 0.0763) and *SIPA1L2* (Nalls et al.: p-value = 7.00e-17, beta = 0.1114; Kim et al.: p-value = 1.00e-17, beta = 0.0269) were both identified by LIME in oligodendrocytes and both showed increased expression in PD. Finally, *PAM* (Nalls et al.: p-value = 2.00e-9, beta = 0.0621) was identified by LIME in DaNeurons and showed increased expression in PD. Indeed, the identification of known PD-associated genes by LIME provides validity to our approach for nominating prospective disease-associated genes.

Finally, we utilized the Neurodegenerative Disease Knowledge Portal (NDKP) to explore gene-level associations of LIME-identified genes with various neurodegenerative diseases, including PD, Alzheimer’s disease (AD), amyotrophic lateral sclerosis (ALS), Lewy body dementia (LBD), and multiple sclerosis (MS) (50) (**Tables S7 and S8**). Our analysis focused on common variant associations using Multi-marker Analysis of GenoMic Annotation (MAGMA) (51), rare variant gene burden analyses, and Human Genetic Evidence (HuGE) Scores (52) (**Figure 5**). *ARL17B*, which was identified by LIME in astrocytes, microglia, and oligodendrocytes, showed a statically significant, common variant association with both PD (MAGMA p-value = 1.03e-8, Z-statistic = 5.61) and AD (MAGMA p-value = 1.53e-6, Z-statistic = 4.67). Gene burden analyses revealed that individuals with PD were significantly enriched for rare variants in *CHORDC1* (p-value = 6.78e-8, beta = 8.73) and *GPC6* (p-value = 7.67e-8, beta = 16.46). Beyond PD, common variation in *HSPA1B*, *JUND*, and *HLA-B* were significantly associated with MS, while individuals with AD were found to be significantly enriched for rare variants in *STIP1*. These results demonstrate the power of our LIME-based approach to nominate relevant genes for targeted genomic analyses aimed at uncovering novel genetic associations.

Considering the comprehensive evidence outlined above, we compiled a list of 15 LIME-identified genes that exhibit the strongest support for involvement in PD and therefore warrant further investigation by different modalities (**Table 2**). Within this list, we noted *GPC6*, which was uniquely identified by LIME in DaNeurons and was of particular interest due to the well-established role of DaNeurons in PD pathophysiology, the enrichment of rare variants in *GPC6* among individuals with PD from the NDKP cohort (50), and its identification as a cis-eQTL in the adult human brain (49). Hereafter, we further explore the role of *GPC6* in PD.

**Table 2.** LIME-identified genes with supporting evidence for implication in Parkinson’s disease.

| Gene | Cell type identified by LIME | Supporting evidence |
| --- | --- | --- |
| *AK5* | DaNeurons | Nominally significant gene-level rare variant association with PD (p-value = 4.25e-2, beta = 6.87) |
| *ARL17B* | Astrocytes, microglia, oligodendrocytes | Significant gene-level common variant association with PD (MAGMA p-value = 1.03e-8, z-statistic = 5.61)<br>Cell type cis-eQTL co-localization with *WNT3* (astrocytes: PP = 0.98, beta = 1.22; microglia: PP = 0.98, beta = 1.29; oligodendrocytes: PP = 0.98, beta = 1.41) |
| *BAG3* | Oligodendrocytes | GWAS PD-associated gene (Nalls et al. 2019: p-value = 2.00e-11, beta = 0.0763) |
| *B2M* | Microglia, oligodendrocytes, OPC | Nominally significant gene-level common variant association with PD (MAGMA p-value = 2.55e-3, z-statistic = 2.8) |
| *CACNA2D3* | DaNeurons | Nominally significant gene-level rare variant association with PD (p-value = 1.70e-3, beta = 3.77) |
| *CHORDC1* | Astrocytes, endothelial, microglia, oligo., OPC, pericytes | Significant gene-level rare variant association with PD (p-value = 6.78e-8, beta = 8.73) |
| *CIRBP* | Oligodendrocytes | Nominally significant gene-level common variant association with PD (MAGMA p-value = 3.03e-2, z-statistic = 1.88) |
| *CRYAB* | Astrocytes, endothelial, microglia, oligo., OPC, pericytes, neurons | Nominally significant gene-level common variant association with PD (MAGMA p-value = 2.96e-2, z-statistic = 1.89) |
| *GPC6* | DaNeurons | Significant gene-level rare variant association with PD (p-value = 7.67e-8, beta = 16.46) |
| *MOBP* | Microglia, oligodendrocytes | Nominally significant gene-level common variant association with PD (MAGMA p-value = 9.27e-3, z-statistic = 2.35) |
| *PAM* | DaNeurons | GWAS PD-associated gene (Nalls et al. 2019: p-value = 2.00e-09, beta = 0.0621) |
| *SEMA5A* | Neurons, DaNeurons | Nominally significant gene-level rare variant association with PD (p-value = 2.23e-3, beta = 4.62) |
| *SIPA1L2* | Oligodendrocytes | GWAS PD-associated gene (Nalls et al. 2019: p-value = 7.00e-17, beta = 0.1114; Kim et al. 2023: p-value = 1.00e-17, beta = 0.0269) |
| *RIMS1* | Astrocytes | GWAS PD-associated gene (Nalls et al. 2019: p-value = 2.00e-10, beta = 0.0657; Kim et al. 2023: p-value = 7.00e-12, beta = 0.0208) |
| *TMEM163* | Microglia | GWAS PD-associated gene (Nalls et al. 2019: p-value = 5.00e-14, beta = 0.0807; Kim et al. 2023: p-value = 5.00e-12, beta = 0.0400)<br>Nominally significant gene-level common variant association with PD (MAGMA p-value = 1.81e-3, z-statistic = 2.91)<br>Cell type cis-eQTL co-localization with *TMEM163* (microglia: PP = 0.99, beta = -0.47) |

Abbreviations: DaNeurons, dopaminergic neurons; eQTL, expression quantitative trait loci; GWAS, genome-wide association study; MAGMA, Multi Marker Analysis of Genomic Annotation; PP, posterior probability; OPC, oligodendrocyte precursor cell; PD, Parkinson's disease.

### Expression and genetic evidence supports GPC6 as a novel PD associated gene

Among the 4,349 HVGs input to the NN classifier, *GPC6* was identified by LIME as being in the 99.5th percentile of important features for distinguishing between PD and healthy DaNeurons (**Figure 6A**). In DaNeurons from the post-mortem midbrain, *GPC6* showed increased expression in individuals with PD compared to healthy controls. However, a major limitation of post-mortem PD brains is the restriction of the analysis to surviving populations of DaNeurons, which may not capture the transcriptional profile of cells most susceptible to degeneration. Accordingly, we elected to corroborate the post-mortem midbrain tissue data with single-cell RNA sequencing (scRNAseq) of induced pluripotent stem cell-(iPSC) derived DaNeurons from individuals with PD (*n* = 29) and healthy controls (*n* = 11) (53) (**Figure 6B**). Reflecting the post-mortem midbrains, *GPC6* showed increased expression in iPSC-derived DaNeurons from individuals with PD compared to those derived from heathy controls (**Figure 6C**). Interestingly, three of the 18 genes identified by LIME were found to be significantly differentially expressed with the same directionality in both post-mortem and iPSC-derived DaNeurons: *PAM*, which has been associated with PD risk through GWAS (8); *GRID2*, whose loss-of-function (LoF) has been identified as the causative genetic defect in spinocerebellar ataxia 18 (54); and *GPC6*.

**Figure 6.**
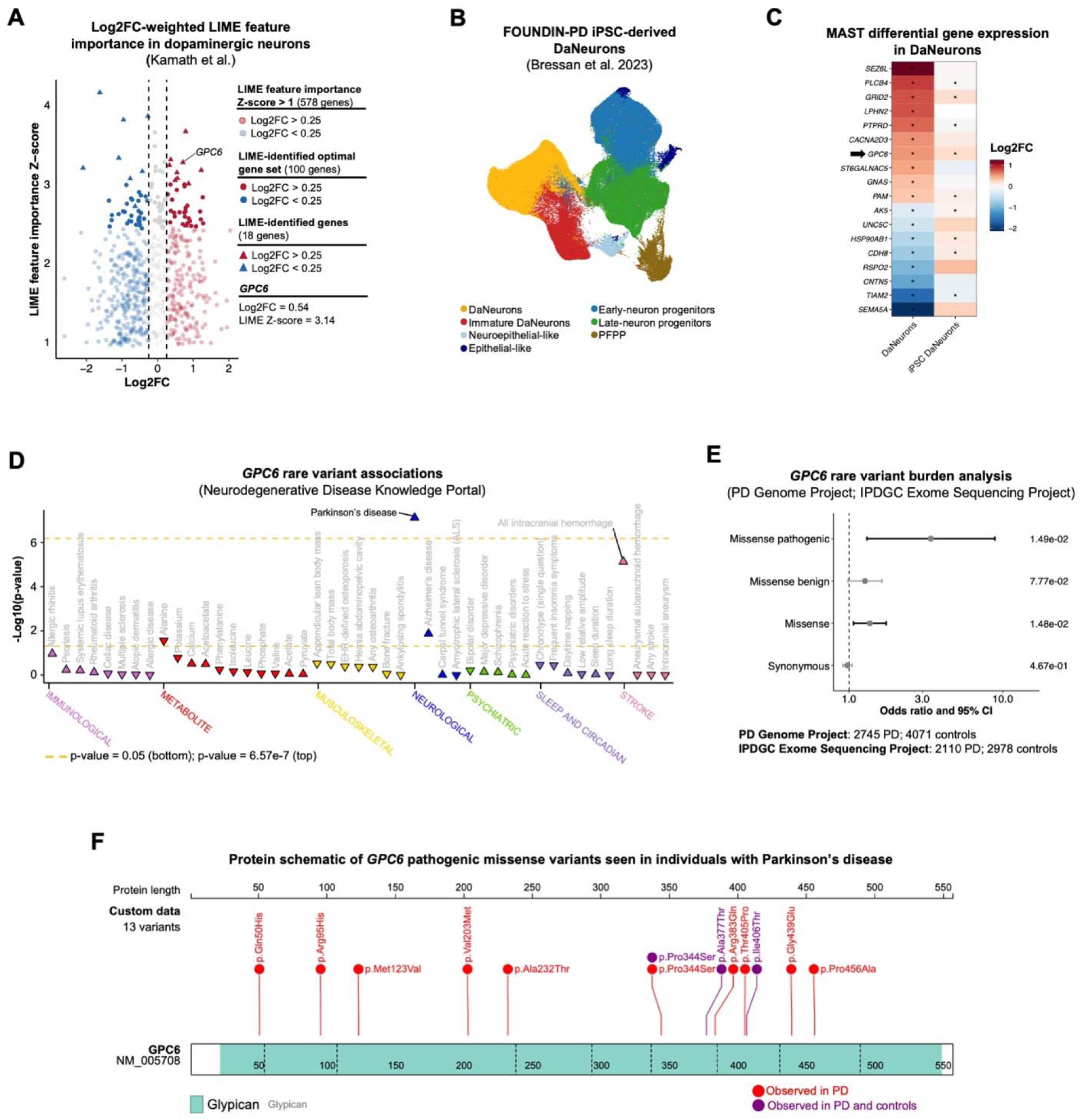
Aberrant expression and rare variant enrichment implicate *GPC6* in PD. **A)** Volcano plot showing the LIME feature importance Z-score and the gene expression log2 fold-change (Log2FC) between Parkinson’s disease (PD) and control dopaminergic neurons (DaNeurons) from the post-mortem substantia nigra (SN). Only highly variable genes (HVG) input to the neural network classifier with a LIME feature importance Z-score > 1 are shown. **B)** Uniform manifold approximation and projection (UMAP) showing the annotated cell types from a single-cell RNA sequencing (scRNAseq) dataset characterizing induced pluripotent stem cells (iPSCs) from individuals with PD and healthy controls differentiated towards a DaNeuron state by Bressan et al. **C)** Heatmap showing the Log2FC of the LIME-identified genes in DaNeurons from the post-mortem SN and iPSC-derived DaNeurons. Asterisks denote significantly differentially expressed genes (adjusted p-value < 0.05) between individuals with PD and healthy controls computed by MAST. **D)** Dot plot showing *GPC6* rare variant associations with various diseases across the Neurodegenerative Disease Knowledge Portal (NDKP) cohorts. **E)** Cochran-Mantel-Haenszel (CMH) tests investigating whether individuals with PD from the PD Genome Project and International Parkinson’s Disease Genomics Consortium (IPDGC) Exome Sequencing Project cohorts were enriched for rare variants in *GPC6*. *P*-values corresponding to each variant category are shown on the right of the plot. The sample sizes for each cohort are shown below the plot. **F)** Protein schematic showing the *GPC6* pathogenic missense variants observed uniquely in PD or in both PD and healthy controls across both cohorts.

We initially elected to further investigate *GPC6* in PD due to the enrichment of rare LoF variants in individuals with PD compared to healthy controls observed in the NDKP cohorts (50) (**Figure 6D**). To corroborate this evidence, we leveraged genomics data from the Parkinson’s Disease Genome Project (PD Genome Project; *n PD =* 2,745; n control = 4,071) and the International Parkinson’s Disease Genomics Consortium (IPDGC) Exome Sequencing Project (*n PD =* 2,110; n control = 2,978) for rare variant gene burden analyses (55) (see Methods). Independent Fisher’s exact tests for each dataset revealed an enrichment of damaging missense variants in individuals with PD compared to controls; however, these enrichments did not meet statistical significance (PD Genome Project: p-value = 7.8e-2, odds ratio = 2.97; IPDGC Exome Sequencing Project: p-value = 7.32e-2, odds ratio = 4.24) (**Figure S26**). To increase the power of our analysis, we performed a combined analysis across both cohorts using the Cochran-Mantel-Haenszel (CMH) test, which revealed a significant enrichment of pathogenic missense variants in individuals with PD (p-value = 1.49e-2, odds ratio = 3.40) (**Figure 6E**). **Figure 6F** shows a schematic of the pathogenic missense variants observed uniquely in PD or in both PD and controls across both cohorts. The aberrant expression of *GPC6* in post-mortem tissue and *in vitro*, coupled with the enrichment of damaging variants in individuals with PD from three separate case-control cohorts implicate *GPC6* in PD.

## Discussion

Identifying genetic contributions to disease is a critical step towards nominating therapeutic targets and developing treatments. Advancements in high-throughput sequencing have facilitated remarkable insight to the genetic determinants of monogenic disease, as well as risk factors for complex, polygenic disease (56). Yet, many genetic determinants of complex, polygenic diseases remain elusive; in PD, risk variants identified through GWAS only explain 16-36% of the disease’s heritability (5). Uncovering this missing heritably, which is further complicated by the interaction of environmental exposures on epigenetics, will afford a stronger understanding of disease biology, which remains a major barrier for clinical translation of targeted therapeutics (56). Due to its unprecedented resolution, scnRNAseq is uniquely suited to help bridge the gap between genetics and biology by revealing intra-cell type gene expression changes between diseased and healthy cells, and nominating genes for targeted genomic analyses to reveal rare-variant associations that may have been overlooked in broader exome-or genome-wide analyses. However, this prospect depends on our ability to identify replicable, disease-associated changes in gene expression from scnRNAseq data, which is complicated by technical inconsistencies and elevated costs that impede the analysis of entire patient cohorts (16).

In this work, we introduced a novel framework that leverages ML models trained to classify single-nuclei transcriptomes based on disease status and incorporated an interpretable model explainer — LIME — to uncover the “reasons” why certain cells are labeled as diseased, revealing the genes whose expression are most influential for the classification decision. By applying this framework to three separate snRNAseq datasets characterizing the midbrain of individuals with PD and healthy controls, we demonstrate the pan-dataset generalizability of the cell-type specific genes identified by LIME, highlight their relevance to the disease, and reveal a novel genetic association in PD by subjecting a LIME-identified gene — *GPC6* — to targeted genomic analysis.

Our framework for identifying gene expression changes in diseased cells has distinct advantages over traditional DGE methods. First, the ML workflow pinpointed the minimal set of genes that warrant further investigation, simplifying the challenge of navigating numerous statistically significant signals often seen in scnRNAseq due to inflated significance. By using permutation tests with decreasing feature counts as input to the classifiers, we optimized the balance between the number of input features and their ability to distinguish between diseased and healthy cells. Second, we directly evaluated the generalizability of the LIME-identified genes obtained from an exploratory dataset by training a separate model on a validation set and benchmarking its classification accuracy against randomly selected features. This potential for pan-dataset investigations helps mitigate the limitations posed by small sample sizes, facilitating cohort-scale studies necessary to address the considerable heterogeneity of complex diseases, such as PD. Furthermore, this approach is well-suited for the anticipated influx of new datasets as the technology becomes increasingly feasible. The greater the number of datasets that a set of LIME-identified genes can accurately classify, the more confident we will be that they adequately capture the molecular features of diseased cells.

An important finding of this study was the identification of cell-type specific LIME optimal gene sets that accurately identified PD cells across three separate datasets (mean balanced accuracy = 0.917), especially when compared to the accuracy achieved when using an equal number of randomly selected genes (mean balanced accuracy = 0.586). However, we note an important distinction between generalizable genes and generalizable models. A generalizable model would suggest that the same NN could accurately classify cells from all three independent dataset. Although generalizable models are necessary for clinical applications, including, for example, diagnosis or predicting patient response to therapeutics, their application to scnRNAseq is complicated by technical variability across datasets and fell beyond the scope of this study (57). In contrast, we aimed to identify generalizable genes — that is, a set of genes that can be input to dataset-specific classifiers to achieve high classification accuracy across all datasets. Indeed, the generalizability of the LIME-identified genes from this study suggest that they captured replicable readouts across datasets, which is critical for establishing their relevance to PD.

We initially hypothesized that the most influential genes for classifying diseased cells would inform PD biology. Employing a systems-level, overrepresentation analysis to the cell type-specific LIME-identified gene sets revealed an enrichment of biological processes that have been well-established in PD. This confirmed that the identified genes were not only valuable for *in silico* disease status classification but also reflect relevant biological processes. The enrichment of genes related to the heat shock and unfolded protein response was most prominent across cell types. All non-neuronal cell types were enriched for genes involved in proteostasis, largely driven by the HSP90- and HSP70-based chaperone machinery, which play key roles during the aggregation of neurotoxic proteins. Importantly, HSP90 and HSP70 have opposing effects on the stability of client proteins: HSP90 inhibits their degradation while HSP70 promotes it (58). Consequently, it has been suggested that inhibiting HSP90 and enhancing HSP70 could help mitigate α-synuclein aggregation in PD (59). In this study, we observed increased expression of both HSP90 and HSP70 in individuals with PD, reinforcing that targeted molecular inhibition of HSP90 warrants further investigation as a potential strategy to combat the aggregation of neurotoxic proteins in the Parkinsonian brain.

While a systems-level approach is valuable for generating new hypotheses about the pathophysiology of neurodegeneration, a gene-level approach is essential for identifying the genetic drivers of aberrant processes and informing disease etiology. Consequently, we employed a reductionist approach to pinpoint the most biologically informative genes identified by LIME. An intriguing finding from this exercise was the identification of well-established genetic determinants from distinct diseases that influenced the classification of PD cells, reinforcing emerging evidence for shared mechanisms among neurological disorders. (60). Namely, *DAB1* (spinocerebellar ataxia 37) (61), *GRID2* (spinocerebellar ataxia 18) (62), and *FTL* (neuroferritinopathy) (63) have all been identified as the monogenic cause for their respective neurological disorders. LIME also identified genes that have been previously associated with PD risk via GWAS, including *RIMS1*, *TMEM163*, *BAG3*, *SIPA1L2*, and *PAM*, supporting our approach for nominating prospective disease-associated genes. Interestingly, these genes were uniquely identified by LIME in distinct cell types, corroborating the growing evidence of the pathophysiological role of multiple cell types in PD (16) and suggesting cell type-specificity of the PD-associated genes. *LRRK2*, a well-established PD-associated, serves as a key example of how certain genes contribute to the disease through specific cell types, exhibiting minimal expression in neurons and high expression in microglia (64). If future studies can support the involvement of the LIME-identified PD-associated genes in specific cell types, as suggested by this analysis, it would enable us to substantially refine our mechanistic hypotheses regarding the etiology of PD.

Beyond, these previously-described genetic-loci associated with PD risk, LIME nominated genes that showed statistically significant, gene-level associations with PD that, to the best of our knowledge, have not been described in the literature. *ARL17B*, which is predicted to be involved in protein trafficking and vesicle-mediated transport, was identified by LIME in astrocytes, microglia, and oligodendrocytes, and showed statistically significant gene-level common variant associations with both PD and AD from the NDKP cohorts (50). Furthermore, the expression of *ARL17B* co-localized with PD-associated variants in *WNT3* in the aforementioned cell types (49), while a transcriptome-wide association study revealed that decreased expression of this gene is correlated with increased PD risk (65), reinforcing the role of altered *ARL17B* expression in PD pathology. Furthermore, individuals with PD from the NDKP cohorts exhibited an enrichment of rare variants in *CHORDC1* and *GPC6*. Rare variants — those with minor allele frequencies ≤ 1% — likely account for a significant proportion of the missing heritability in PD, as their contributions are not currently captured by GWAS due to limited sample sizes (5). Thus, a robust method that can identify these genes is critical to establish a comprehensive understanding of the genetic architecture of complex, polygenic traits like PD.

Among the most intriguing genes identified by LIME was *GPC6*, a glycosylphosphatidylinositol-anchored heparan sulfate proteoglycan that was uniquely identified in DaNeurons. One major challenge of transcriptomics is deciphering between gene expression changes that stem from genetic factors, those that stem from environmental exposures, and those that occur in response to the disease process. The application of scRNAseq to patient-derived iPSCs can mitigate this limitation since it is unlikely that environmental factors and pathophysiology-induced transcriptional changes persists after iPSC differentiation (66). In this work, we observed that *GPC6* showed increased expression in iPSC-derived DaNeurons from individuals with PD compared to those derived from heathy controls, reflecting the transcriptional patterns observed in post-mortem midbrains. When combined with the enrichment of rare *GPC6* variants observed in individuals with PD from the NDKP, PD Genome, and IPDGC cohorts, the results suggest that *GPC6* may contribute to the complex interplay among variants of moderate effect size in sporadic PD. Recent studies have demonstrated that heparan sulfate proteoglycans play a role in the intracellular accumulation of α-synuclein preformed fibrils, thereby facilitating disease propagation through the cell-to-cell spread of fibrils in synucleinopathies (67). Notably, we observed that, similar to PD, individuals with AD from the NDKP cohorts showed an enrichment of rare variants in *GPC6*. Heparan sulfate proteoglycans have been observed to promote the conversion of non-fibrillar amyloid-β — the main component of amyloid plaques in AD — into neurotoxic fibrillar amyloid-β (68). Although further functional experiments are required, it is tempting to speculate that DaNeurons may compensate for the LoF or aberrant function of *GPC6* by increasing the expression of related heparan sulfate proteoglycans, which could include *GPC6* in the presence of feedback regulation, to promote α-synuclein fibril binding and uptake in this vulnerable cell type. In line with this hypothesis, GPC6 was identified as a cis-eQTL that enhanced expression in excitatory neurons (49); however, it remains to be seen whether select variants in *GPC6* produce similar expression changes in DaNeurons. On the other hand, recent findings from animal models have revealed the importance of glycosylphosphatidylinositol-anchored heparan sulfate proteoglycan in the proper function of synapses (69). Therefore, it is possible that aberrant *GPC6* promotes synaptic dysfunction and triggers DaNeuron degeneration. Of course, both hypotheses are not mutually exclusive as synaptic dysfunction related to α-synuclein aggregates may lead to PD (42).

In addition to *GPC6*, we have outlined 15 LIME-identified genes with the strongest evidence for their implication in Parkinson’s disease in **Table 2**. It is our intention that these genes inspire future investigations in the field to further characterize their implication in PD through multi-omic approaches and functional validation. Beyond these 15 genes, those that were identified by LIME and have been implicated as the monogenic cause of distinct neurological disorders, including *DAB1*, *GRID2*, *FTL*, are of equal merit of further investigation in order to elucidate the prospective intersection of genetic determinants in brain diseases.

We anticipate that this framework for identifying molecular features that distinguish diseased cells from their healthy counterparts will prove opportunistic across diseases in biomedical research, either through a pan-dataset approach or applied to individual datasets. While we applied a reductionist strategy largely tailored to our intentions of pinpointing the most biologically relevant genes for targeted genomic analyses, it is likely that investigators will have varying intentions for the data. Indeed, the LIME outputs derived from the provided pipeline are primed for customization. In this initial, proof-of-concept study we elected to focus primarily on HVGs, but that does not discount the value of the other interpretable feature selection methods, such as NMF, applied in a similar framework, which warrant further investigation. Of course, an inherent limitation of inputting HVGs to the classifiers is limiting discovery to a subset of genes characterized in the dataset. Leveraging interpretable dimensionality reduction techniques can mitigate this limitation, but a careful balance must be struck between maintaining sufficient features for discovery and introducing noise into the classifiers. Overall, we conclude that our interpretable, ML-based approach for identifying characteristic gene expression changes in diseased cells constitutes a notable advancement towards the pressing demand for novel approaches to further resolve the intricate genetic architecture of complex, polygenic diseases such as PD.

## Methods

### Data acquisition

#### Single-nuclei RNA sequencing of post-mortem midbrains

We leveraged three snRNAseq datasets characterizing the midbrain of individuals with PD and healthy controls prepared by Kamath et al. (13), Wang et al. (14), and Smajic et al. (15) as previously described. For Kamath et al., filtered feature-barcode expression matrices and the corresponding metadata were obtained from the Single Cell Portal with accession SCP1768 and filtered to exclude LBD samples. For Wang et al., filtered feature-barcode expression matrices were obtained from the gene expression omnibus (GEO) with accession code GSE184950. Samples were matched to the metadata published in the supplemental material of the corresponding manuscript and filtered to only include individuals with PD and healthy controls, excluding PD samples with frontotemporal dementia. For Smajic et al., filtered feature-barcode expression matrices were obtained from GEO with accession code GSE157783 and matched to the metadata published in this repository.

Pre-processing and quality control of the snRNAseq data was performed independently for each dataset using Seurat (v4.3.0.1) (70) within the scRNAbox analytical pipeline (19). Cells were filtered based on the number of unique features and total counts per cell, adhering to the thresholds utilized in the original manuscripts (13–15). Cells with mitochondrial reads > 10% and ribosomal proteins > 10% were excluded. Putative doublets were detected and removed with DoubletFinder (v2.0.4) (71). Cells were then clustered on the basis of their transcriptional similarity to facilitate cell type annotation using Louvain network detection. Broad cell type annotations were informed by known marker genes and gene enrichment analyses. Marker genes for each cluster were identified with Seurat’s *FindAllMarkers* function using the Wilcoxon rank-sum test. Significant DEGs (Log2FC > 0.25 and p-value < 0.05) were analyzed with EnrichR (72), and cell types were predicted using the *Cell Marker Augmented 2021* (73) and *Azimuth Cell Types 2021* (74) libraries. To assess cell type replicability across the datasets, we utilized MetaNeighbor (v1.18.0) (32), which evaluates the transcriptional similarity of cell types from independent datasets based on their nearest neighbour frequency in a shared gene expression space. Classification models were trained on the reference dataset using the *trainModel* function, and the AUROC values for the target dataset were computed with the *MetaNeighborUS* function. To enable the implementation of the ML models on the snRNAseq data in Python (v3.8.10), the raw count data (RNA assay) from the dataset-specific Seurat objects were converted to AnnData objects for compatibility with Scanpy (v1.9.2) (75) using the SeuratDisk (v0.0.0.9021) package.

#### Single-cell RNA sequencing of induced pluripotent stem cell-derived dopaminergic neurons

To corroborate the gene expression patterns of LIME-identified genes observed in post-mortem DaNeurons, we leveraged a scRNAseq dataset characterizing iPSC-derived DaNeurons from individuals with PD and healthy controls produced as part of the Foundational Data Initiative for Parkinson’s Disease (FOUNDIN-PD) (53). In brief, the iPSC lines were differentiated towards a DaNeuron fate, underwent scRNAseq after day 65 of differentiation, and were annotated to seven distinct, broad cell types, as previously described (53). A controlled access, fully processed Seurat object was obtained from the Parkinson’s Progression Markers Initiative (PPMI) data portal (https://www.ppmi-info.org/; accessed 09-17-2023) and filtered to only include the cells annotated as DaNeurons in the 65 days differentiated iPSC cultures, from individuals with idiopathic PD and healthy controls.

### Feature selection for input into machine learning classifiers

Feature selection was performed to reduce the number of features input into the ML models for disease status classification. Raw snRNAseq counts were first normalized to ensure equal total counts per cell using the *normalize_total* Scanpy function, and then log-transformed with the *log1p* Scanpy function prior to feature selection. Feature selection was performed separately for each cell type and for all cells together within the two exploratory datasets (Kamath et al. and Wang et al.). To maintain the independence of the test sets from the training phase, feature selection was performed on the 80% of the dataset allocated for training the classifiers. To identify the most effective methods for classifying single-nuclei transcriptomes by disease status, we tested four feature selection techniques: i) HVGs, ii) PCA, iii) NMF, and iv) ETM.

#### Highly variable genes

HVGs are defined as genes exhibiting substantial expression variability across all cells, irrespective of disease or treatment status. To identify HVGs, we applied Scanpy’s *highly_variable_genes* function to the test set, which categorizes genes into 20 bins based on their average expression levels across individual cells. The function then z-normalizes the dispersion measures within each bin to highlight genes with significantly variable expression relative to others with similar average expression levels (76). For HVG selection, genes were required to have a minimum average expression level of 0.25, a maximum average expression level of 3, and a minimum dispersion of 0.5. The single-cell expression levels of the cell type- and dataset-specific HVGs were input to the ML models for disease status classification.

#### Principal component analysis

PCA was performed to reduce the dimensionality of single-cell gene expression data by transforming the feature-barcode expression matrices into a set of PCs. PCA was performed on 80% of the dataset designated for training the classifiers using scikit-learn (v1.2.1) (77). This dimensionality reduction model was then applied to project the test set into the same PC space. To determine the optimal number of PCs to retain for each cell type, we applied Seurat’s *JackStraw* function to evaluate their significance and retained all PCs up to the first one that did not meet statistical significance (p-value < 0.05). The result of the PCA dimensionality reduction is a cell-by-PC matrix where each entry represents a cell’s coordinates in PC space. These cell type- and dataset-specific matrices were input into the ML models for disease status classification.

#### Non-negative matrix factorization

NMF was performed to reduce the dimensionality of single-cell gene expression data by decomposing the feature-barcode expression matrices into a set of non-negative components. NMF generates two matrices: the basis matrix *W* (cell-component matrix) and the coefficient matrix *H* (feature-component matrix). The basis matrix *W* represents the components that capture the underlying patterns in the data, while the coefficient matrix *H* provides the feature weights for these components. NMF was performed on 80% of the dataset allocated for training the classifiers using scikit-learn. The decomposition model was then applied to the test set to project it into the same component space. To determine the optimal number of components for each cell type, we first performed NMF on the test set with varying numbers of components, ranging from 1 to 100. For each component count, we reconstructed the original data matrix by multiplying the *W* and *H* matrices and computed the Residual Sum of Squares (RSS) to assess the reconstruction error. We plotted the RSS values against the number of components and used the *uik* function from the inflection (v1.3.6) R package to identify the elbow of the curve using the Unit Invariant Knee (UIK) method (78). The number of components corresponding to this elbow was selected as the optimal number for NMF. The cell type- and dataset-specific *W* matrices were input into the ML models for disease status classification.

#### Embedded topic modelling

We leveraged scETM to perform embedded topic modeling and uncover latent topics within data (31). In brief, scETM factorizes the feature-barcode expression matrices into three components: a topics matrix (θ), which captures the underlying structure of the data; a topics-by-embedding matrix (α), which represents the distribution of cells across latent topics; and an embedding-by-genes matrix (ρ), which captures the contribution of each gene to the topics. The scETM model was initially trained on 80% of the dataset allocated for training the classifiers using default parameters, with the number of epochs set to 100. The trained model was then applied to the test set to generate cell embeddings in the same latent topic space. The resulting cell type- and dataset-specific unnormalized cell topics mixture, which described the cell embeddings across the latent topics, were input into the ML models for disease status classification. To determine the optimal number of topics, we trained and evaluated scETM models with 10, 25, 50, 75, and 100 topics and input the resulting cell embedding into the ML disease status classifiers. The optimal number of topics was selected as that which yielded the highest disease status classification accuracy, on average, across all tested classifiers, including NN, RF, LR, and SVM.

### Machine learning models for disease status classification

We trained ML models to classify single-nuclei transcriptomes according to disease status — PD or healthy control. The ML models were implemented using scikit-learn in Python. The cell type- and dataset-specific AnnData objects were divided into 80% to train the ML models and 20% to evaluate the classification accuracy. As input into the ML classifiers, we used the features obtained from the feature selection methods described above — that is, cell-level expression of the HVGs, cell-level PC embeddings from PCA, cell-level component embeddings from NMF, or cell-level topic embeddings from ETM. During the training phase, we first employed five-fold cross-validation to optimize the model’s hyperparameters and ensure robust performance estimates across different subsets of the dataset. During the cross validation, the training data is further split into 80% for training and 20% for validation, creating five distinct training/validation sets. The five independent models were then applied to the 20% of the unseen test data reserved for model evaluation. To identify the most effective method for classifying single-nuclei transcriptomes according to disease status, we tested four ML classifiers: i) NN, ii) logistic regression, iii) random forest, and iv) support vector machine.

#### Neural network

We implemented NNs for disease classification using the *MLPClassifier* model class from scikit-learn, configured with one hidden layer and 100 nodes, the Rectified Linear Unit (ReLU) activation function for non-linearity, and the Adam optimization algorithm. The training process was limited to a maximum of 500 iterations (77).

#### Logistic regression

We implemented logistic regression for disease classification using the *LogisticRegression* model class from scikit-learn, configured with an L1 regularization penalty, a regularization strength of 2.0, and the liblinear solver. The training process was limited to a maximum of 100 iterations (77).

#### Random forest

We implemented random forest for disease classification using the *RandomForestClassifier* model class from scikit-learn, configured with 100 distinct decision trees and the Gini impurity measure to evaluate the quality of splits at each node in the trees. The minimum number of samples required to split an internal node was set to 2 and the minimum number of samples required to be a leaf node was set to 1. Bootstrapping was enabled to ensure that each tree was trained on a randomly subsampled subset of the data with replacement (77).

#### Support vector machine

We implemented SVM for disease classification using the *SVC* model class from scikit-learn, configured with a linear kernel and a regularization strength of 1. For SVM, we did not set a limit on the number of iterations for the training process (77).

### Machine learning classifier performance evaluation

We evaluated the performance of the ML classifiers using balanced accuracy, which is well-suited for binary classification tasks involving imbalanced datasets, such as when the number of cells from individuals with PD differs considerably from those of healthy controls. Balanced accuracy was calculated as follows: 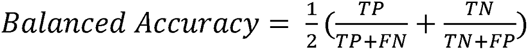, where *TP* denotes the number of cells correctly classified as PD; *TN* denotes the number of cells correctly classified as healthy control; *FP* denotes the number of cells incorrectly classified as PD; and *FN* denotes the number of cells incorrectly classified as healthy control. A cell was deemed correctly classified if the disease status predicted by the ML classifiers matched the actual disease status of the individual from whom the cell was derived. We employed the Wilcoxon rank-sum test for inter-model performance comparisons.

### *In silico* simulations to evaluate machine learning classifier performance

We conducted in silico simulations to evaluate how various dataset metrics, including the number of cells available for model training and sequencing depth, influence the performance of NN classifiers. To investigate the impact of cell numbers on model performance, we trained and evaluated the classifiers using 100%, 75%, 50%, or 25% of the available cells for each cell type. We used the number of unique genes per cell and unique molecular identifiers (UMI) per cell as proxy metrics for sequencing depth to assess their effect on NN performance. For both metrics separately, we divided the cell type-specific cells into quartiles, and then trained and evaluated the classifiers using each quartile independently. In addition, we evaluated the impact of randomized disease status labels — PD or healthy control — on the performance of the NN classifiers. For each cell type independently, we randomized either 0%, 25%, 50%, 75%, or 100% of the disease status labels, and then trained and evaluated the classifiers. Here, cells were deemed correctly classified if the NN’s prediction matched the randomized disease status label.

### Application of LIME to machine learning classifiers

We leveraged LIME to decode the NN classifiers and elucidate the most important genes for classifying a cell as either diseased or healthy control (30). LIME perturbs proximal data points in *d*-dimensional feature space — that is, LIME alters gene expression patterns of neighbouring cells — and observes the deviations in the classification predictions when the perturbed data are input to a simple model intended to mimic the complex model. For each cell, LIME returns a ranked list of all features input to the complex model ordered according to their influence on the classification decision. LIME was implemented using the *LimeTabularExplainer* function from the lime library in Python (30).

To approximate global explanations from LIME’s local explanations and identify the most important features for classifying PD cells, we first calculated the average feature importance of each gene across cells belonging to an individual PD subject using the formula 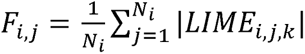, where *F_i,j_* represents the average absolute feature importance for gene *j* in subject *i*, *N_i_* is the number of test cells for that subject, and *LIME_i,j,k_* is the LIME feature importance for gene *j* in the *k*-th cell of subject *i*. This process produced distinct lists of LIME feature importance values for each subject. Next, we averaged these values across all PD subjects using the formula 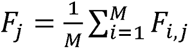, where *M* is the number of PD subjects, resulting in a single ranked list of features. To prioritize genes with consistent expression across cells — those more likely to represent true biological signals — we computed the percent expression of each feature using 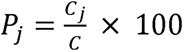, where *P_j_* is the percent expression of gene *j*, *C_j_* is the number of cells expressing gene *j*, and *C* is the total number of cells. We then normalized these percent expression values so that they sumed to 1 with the equation 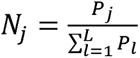, where *N* is the normalized percent expression of gene *j* and *L* is the total number of genes. This normalization allowed us to weight the LIME importance scores as *W_j_ = N_j_ X F_j_*. Finally, we applied a Z-score transformation to standardize the weighted importance score for comparison across datasets, calculated as 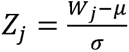, where *µ* is the mean of the weighted scores and *σ* is the standard deviation of those scores.

### Multi-step process to generate the final LIME-identified gene set

We employed a six-step process to refine the list of cell type-specific LIME-identified features and pinpoint the most informative genes for classifying PD cells. In step 1, we applied LIME to decode the cell type- and dataset-specific NN classifiers from both exploratory datasets: Kamath et al. and Wang et al. This process provided distinct ranked lists of HVGs ordered according to their LIME feature importance Z-score value for each cell type from each dataset, as described above.

In step 2, we identified cell type-specific genes that were consistently important for disease status classification across both exploratory datasets. Utilizing the outputs from step 1, we computed the cell type-specific, average LIME feature importance Z-scores across both exploratory datasets for all unique HVGs initially input into the NN classifiers. This process generated unified, cell type-specific lists describing the average LIME feature importance across both datasets, from which we only retained genes with an average Z-score > 1. Notably, the Wang et al. dataset characterized insufficient DaNeurons for the ML workflow; thus, for this cell type we retained genes with a Z-score > 1 in Kamath et al. only.

In step 3, we performed permutation tests using progressively fewer genes as input to the NN models to determine the optimal number of cell type-specific genes for disease status classification. We assigned percentile ranks to all HVGs based on their mean LIME feature importance Z-score across both datasets, with a rank of 1.00 indicating the highest importance. Starting from the percentile rank corresponding to a mean Z-score = 1, we trained a NN disease status classifier using all genes meeting this threshold and evaluated its performance. We then incrementally increased the percentile threshold by 0.001, thereby decreasing the number of genes input to the NN model, and re-trained and re-evaluated the classifier. We repeated this process until we reached a percentile rank of 1.00. We performed the same permutation tests with both randomly selected HVGs and randomly selected genes as a benchmark to compare the NN classifiers performance when using LIME-identified genes. We defined the optimal number of cell type-specific genes as that which maximizes the sum of the discrepancies in NN balanced accuracy when using LIME-identified genes versus random genes, across both exploratory datasets. Let *NN_LIME_* and *NN_gene_* be the balanced accuracy of the NN classifier using LIME-identified genes and randomly selected genes, respectively, and let *ΔACC_dataset_* be the discrepancy in the balanced accuracy of the NN classifiers when using LIME-identified genes or random genes for a give exploratory dataset, then our method is represented mathematically as *Optimal gene set = arg max_G_ (ΔACC_Kamath et al._ + ΔACC_Wang et al._)*, where *ACC_dataset_ = NN_LIME_ - NN_gene_*. This process resulted in LIME optimal gene sets for each cell type, informed by the LIME feature importance values across both exploratory datasets.

In step 4, we used the cell type-specific LIME optimal gene sets to classify cells in the Smajic et al. validation dataset and assess the generalizability of the gene signatures. Specifically, we trained cell type-specific NN classifiers on this dataset, using the LIME optimal gene sets identified in step 3 as input. For comparison, we trained additional NN classifiers using an equal number of randomly selected HVGs and total genes, matched to the respective cell type-specific optimal gene set.

In step 5, we filtered the cell type-specific LIME optimal gene sets to only include genes that met the designated threshold for change in expression between PD and healthy cells. We first integrated the Kamath et al., Wang et al., and Smajic et al. datasets to create unified, cell type-specific datasets characterizing all of the subjects in the analysis. We then computed the Log2FC for all genes in the corresponding cell-type specific LIME optimal gene set and only retained those genes with an absolute value Log2FC > 0.25 between PD and healthy control cells. Additionally, we retained genes previously associated with PD through GWAS and excluded mitochondrially encoded genes, ribosomal proteins, sex-linked genes, and non-coding RNAs.

In step 6, we retained the genes if they met at least one of two criteria: i) mean LIME feature importance Z-score across all datasets > 5 in a particular cell type or ii) mean LIME feature importance Z-score > 2 across all datasets in a particular cell type but identified by LIME in ≥ 3 cell types. Importantly, for DaNeurons, which displayed a LIME feature importance Z-score distribution suggestive of a greater number of genes with a moderate contribution to the NN classification decision, we employed a Z-score threshold > 3.

### Genetic analysis of LIME-identified genes

For the characterization of the LIME-identified genes, we used publicly available databases with genetic and expression data on PD and other neurodegenerative diseases, including cell type-specific cis-eQTLs, PD GWAS, and gene-level associations from the NDKP.

#### Cell type cis-expression quantitative trait loci

To determine whether the LIME-identified genes were previously reported as eQTLs, we used cell type-specific cis-eQTL summary statistics from Bryois et al. (49). In brief, cell type cis-eQTLs were identified through snRNAseq of eight brain cell types sourced from the post-mortem prefrontal cortex, temporal cortex, and white matter of 192 individuals, as previously described (49). In our analysis, we reported cell type cis-eQTLs with an FDR-adjusted p-value < 0.05 that were identified in the same cell type as the corresponding LIME-identified gene. For LIME-identified genes in neurons and DaNeurons, we specifically reported cis-eQTLs from both inhibitory and excitatory neurons as documented by Bryois et al. Similarly, for cell type cis-eQTL co-localization with PD-associated genes, we reported associations with a PP > 0.70 that were identified in the same cell type as the corresponding LIME-identified gene.

#### Parkinson’s disease genome wide association study

To determine whether the LIME-identified genes had been previously associated with PD, we utilized PD GWAS summary statistics from Nalls et al. (8) and Kim et al. (9), available from the GWAS catalog (https://www.ebi.ac.uk/gwas/; accessed 08-01-2024). Both studies employed a meta-analysis approach; Nalls et al. analyzed 37,688 individuals with PD, 18,618 UK Biobank proxy-cases, and 1,394,791 healthy controls of European ancestry (8), while Kim et al. analyzed 49,049 individuals with PD, 18,618 UK Biobank proxy-cases, and 2,458,063 healthy controls from diverse ancestries, including European, East Asian, Latin American, and African (9). In our analysis, we reported the mapped gene for associations that achieved genome-wide significance (p-value < 5.00e-8) and were identified by LIME.

#### Neurodegenerative Disease Knowledge Portal gene-level associations

Gene-level association statistics were obtained from the NDKP (https://ndkp.hugeamp.org/; accessed 08-05-2024). We investigated common variant associations identified through MAGMA (51), rare variant gene burden analyses, and HuGE Scores (52) for PD, AD, ALS, LBD, and MS. The methodologies and datasets used in the NDKP have been previously described (50). For common and rare variant associations, which are pre-computed in the NDKP database, we reported all LIME-identified genes with nominally significant associations (p-value < 0.05) and highlighted those with a MAGMA p-value < 2.5e-6 and gene burden analyses achieving exome-wide significance (p-value < 6.57e-7). HuGE Scores, which provide insights into the genetic support for a gene’s involvement in disease based on rare and common variation were computed using the HuGE Calculator (https://ndkp.hugeamp.org/hugecalculator.html; 08-05-2024). We reported LIME-identified genes with a HuGE score of ≥ 3, indicating *Moderate* evidence for their involvement in disease.

### *GPC6* rare variant gene burden analyses

To corroborate the enrichment of rare variants in *GPC6* in individuals with PD compared to healthy controls, which we initially observed in the NDKP, we performed rare variant gene burden analyses using two independent case-control cohorts.

#### Participants and genetic sequencing

We obtained genomics data from the PD Genome Project and the IPDGC Exome Sequencing Project, downloaded from the Parkinson’s Disease Variant Browser (https://pdgenetics.shinyapps.io/VariantBrowser/; 08-25-2024) (55). The PD Genome Project includes whole-genome sequences from 2,745 individuals with PD and 4,071 healthy controls. The IPDGC Exome Sequencing project includes whole-exome sequences from 2,110 individuals with PD and 2,978 healthy controls. As part of their integration into the Parkinson’s Disease Variant Browser, both datasets were subjected to rigorous quality control measures, including disease status and sequencing depth assessment at a sample level, data trimming to exome calling regions, and relatedness assessment, as previously described (55).

#### Variant annotations and filtering

Variants in *GPC6* were annotated with CADD scores (v1.6) (79), AlphaMissense (80), variant consequences, Genome Aggregation Database (gnomAD; v2.1.1) allele frequency and allele counts (81), and the Human Genome Variant Society (HGVS) coding sequence and protein nomenclature using the Variant Effect Predictor (VEP; v.112.0) (82). Coding variant consequences were reclassified into three categories: (1) protein-truncating variant (PTV), (2) missense, and (3) synonymous. PTVs included those classified as “splice_donor_variant,” “splice_acceptor_variant,” “frameshift_variant,” “stop_gained,” “start_loss,” and “stop_loss”; however, only one PTV variant was observed across both dataset and were therefore excluded from the analyses. Missense variants included those classified as “missense_variant”, “missense_variant,splice_region_variant”, “inframe_insertion”, and “inframe_deletion”. We further classified missense variants into missense damaging and missense benign. Missense damaging variants included those with an AlphaMissense “pathogenic” annotation or an AlphaMissense “ambiguous” annotation and a CADD score > 20.000, which denotes the 1% most deleterious variants (79). We filtered the datasets to only include those that were rare in the general population (allele frequency < 0.01 in gnomAD).

#### Quantification of the enrichment of rare variants in individuals with PD

To assess the enrichment of rare variants in *GPC6* in individuals with PD, we conducted a two-sided Fisher’s exact test. Rare variants were categorized into bins as described, and we compared the proportions of observed variants in individuals with PD to those in healthy controls within each bin, analyzing the data separately for the PD Genome Project and the IPDGC Exome Sequencing Project. We then performed a combined analysis across both cohorts using the CMH test, which, in our case, assesses the association between rare variants in *GPC6* and disease status while controlling for cohort as the stratifying variable (83).

### Differential gene expression analysis

Cell type-specific DGE was calculated using MAST (36) and DESeq2 (37), implemented in the scRNAbox analytical pipeline as previously described (19). MAST, which is tailored to single-cell gene expression data, treats each cell as an independent replicate and employs a two-part generalized linear model to account for both the presence and the continuous level of gene expression. Conversely, DESeq2 utilizes a pseudo-bulk approach, aggregating counts from all cells of a given subject and treating each subject as a independent replicate. It models the count data with a negative binomial distribution and estimates the size factors for normalization. Genes with a Bonferroni-adjusted p-value < 0.05 and an absolute value Log2FC > 1.00 were deemed differentially expressed.

### Gene set overrepresentation analysis

Gene set overrepresentation analysis was performed using the g:Profiler web interface (https://biit.cs.ut.ee/gprofiler/gost; 07-22-2024) (84), configured to use all genes identified in the snRNAseq midbrain datasets as the statistical domain scope. Terms with a g:SCS adjusted p-value < 0.05 were considered significantly overrepresented. The g:SCS method for multiple hypothesis testing correction was employed because it is more appropriate for overrepresentation analyses, as it accounts for the hierarchical relationships among predefined gene sets, which consist of related general and specific terms, unlike Bonferroni and Benjamin-Hochberg methods which assume independent tests (85).

### Statistical analyses

We performed all statistical analyses using the R statistical software (v4.2.2) (86). We used the ggplot2 R package (v3.4.2) for data visualization (87).

## Declarations

### Ethics approval and consent to participate

Not applicable.

### Consent for publication

Not applicable.

### Availability of data and analysis code

SnRNAseq data prepared by Kamath et al. is available from the Single Cell Portal (https://singlecell.broadinstitute.org/single_cell/study/SCP1768). SnRNAseq data prepared by Wang et al. is available from the GEO with accession code GSE184950. SnRNAseq data prepared by Smajic et al. is available from the GEO with accession code GSE157783. ScRNAseq data chracterizing the iPSC-derrived DaNeurons prepared by Bressan et al. are available from the PPMI database (www.ppmi-info.org/access-dataspecimens/download-data), RRID:SCR 006431. For up-to-date information on the study, visit www.ppmi-info.org. The genomics data from the PD Genome Project and IPDGC Exome Sequencing Project are available from the Parkinson’s Disease Variant Browser (https://pdgenetics.shinyapps.io/VariantBrowser/). The code used for the analyses presented in the work, as well as the NN-LIME pipeline are available on GitHub (https://github.com/mfiorini9/scRNAseq-NN-LIME).

### Authors’ contributions

All authors conceived the study. MRF and JL wrote the machine learning classifier Python code. MRF developed and implemented the LIME-based framework. MRF developed the NN-LIME pipeline and created the GitHub site. MRF performed the formal analyses. MRF produced the figures and tables. MRF, EAF, SMKF, and RAT interpreted the data. MRF wrote the manuscript with input from all authors. SMKF and RAT supervised the project.

## Acknowledgments

The authors acknowledge Dr. Allison Dilliott for her helpful insight throughout the project.

## Authors’ information

MRF is supported by a CIHR Canada Graduate Scholarships-Master’s Award and a Fonds de Recherche Santé Québec Master’s Award. EAF holds a Canada Research Chair (Tier 1) in Parkinson’s disease. SMKF received funding from Brain Canada and the Montreal Neurological Institute-Hospital. RAT received funding through the McGill Healthy Brains for Healthy Lives (HBHL) Postdoctoral Fellowship and Molson NeuroEngineering Fellowship.

