## Additional File 1 for "Interpretable machine learning classifiers implicate *GPC6* in Parkinson’s disease from single-nuclei midbrain transcriptomes"

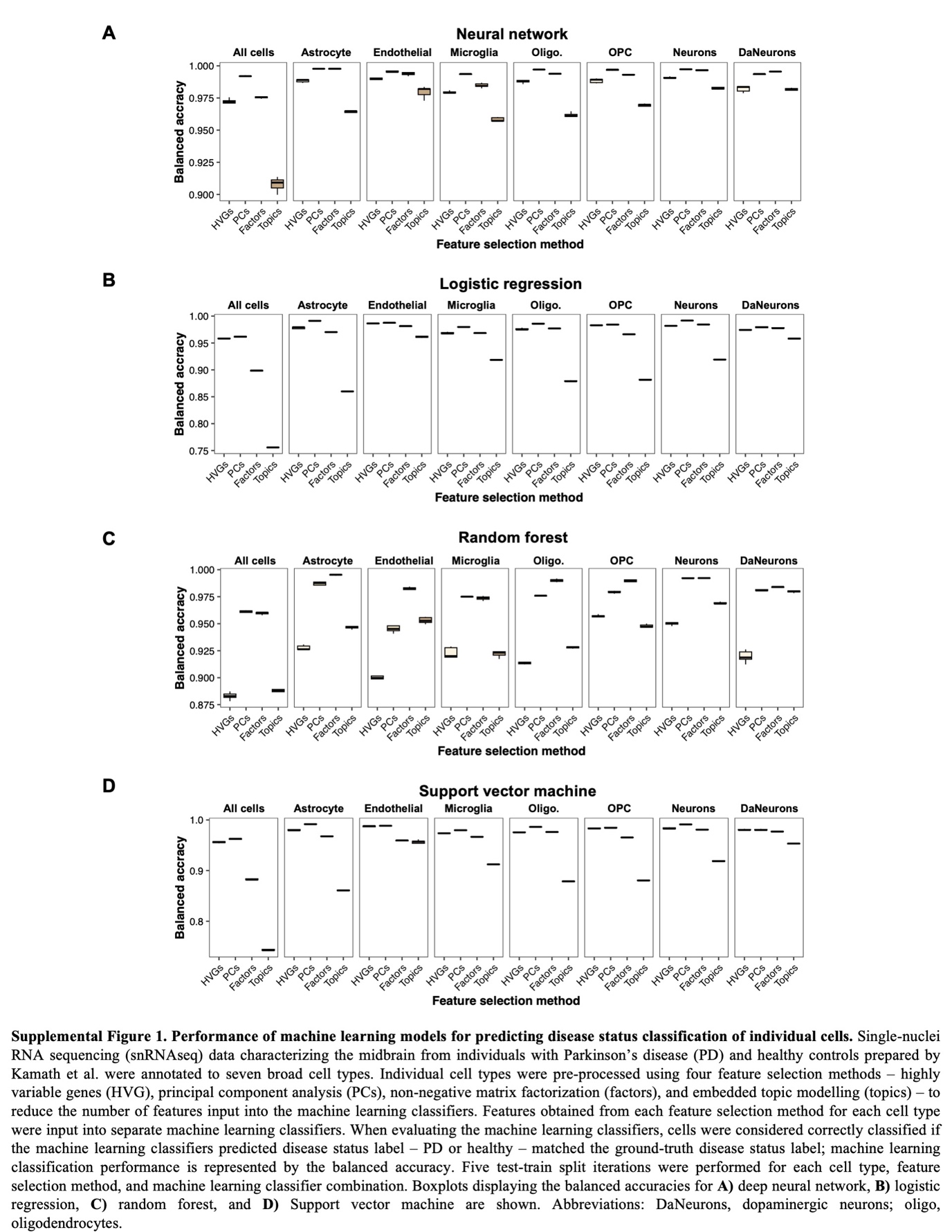

**Figure S1. Performance of machine learning methods for classifying single-nuclei transcriptomes by disease status.** Single-nuclei RNA sequencing (snRNAseq) data characterizing the midbrain from individuals with Parkinson’s disease (PD) and healthy controls prepared by Kamath et al. were annotated to seven broad cell types. Individual cell types were pre-processed using four feature selection methods —highly variable genes (HVG), principal component analysis (PCs), non-negative matrix factorization (factors), and embedded topic modelling (topics) — to reduce the number of features input into the machine learning classifiers. Features obtained from each feature selection method for each cell type were input into separate machine learning classifiers. When evaluating the machine learning classifiers, cells were considered correctly classified if the machine learning classifiers predicted disease status label — PD or healthy — matched the ground-truth disease status label. Machine learning classification performance is represented by the balanced accuracy. Boxplots displaying the balanced accuracies following five-fold cross validation are shown for **A)** neural network, **B)** logistic regression, **C)** random forest, and **D)** Support vector machine. Abbreviations: DaNeurons, dopaminergic neurons; oligo, oligodendrocytes; OPC, oligodendrocyte precursor cells.

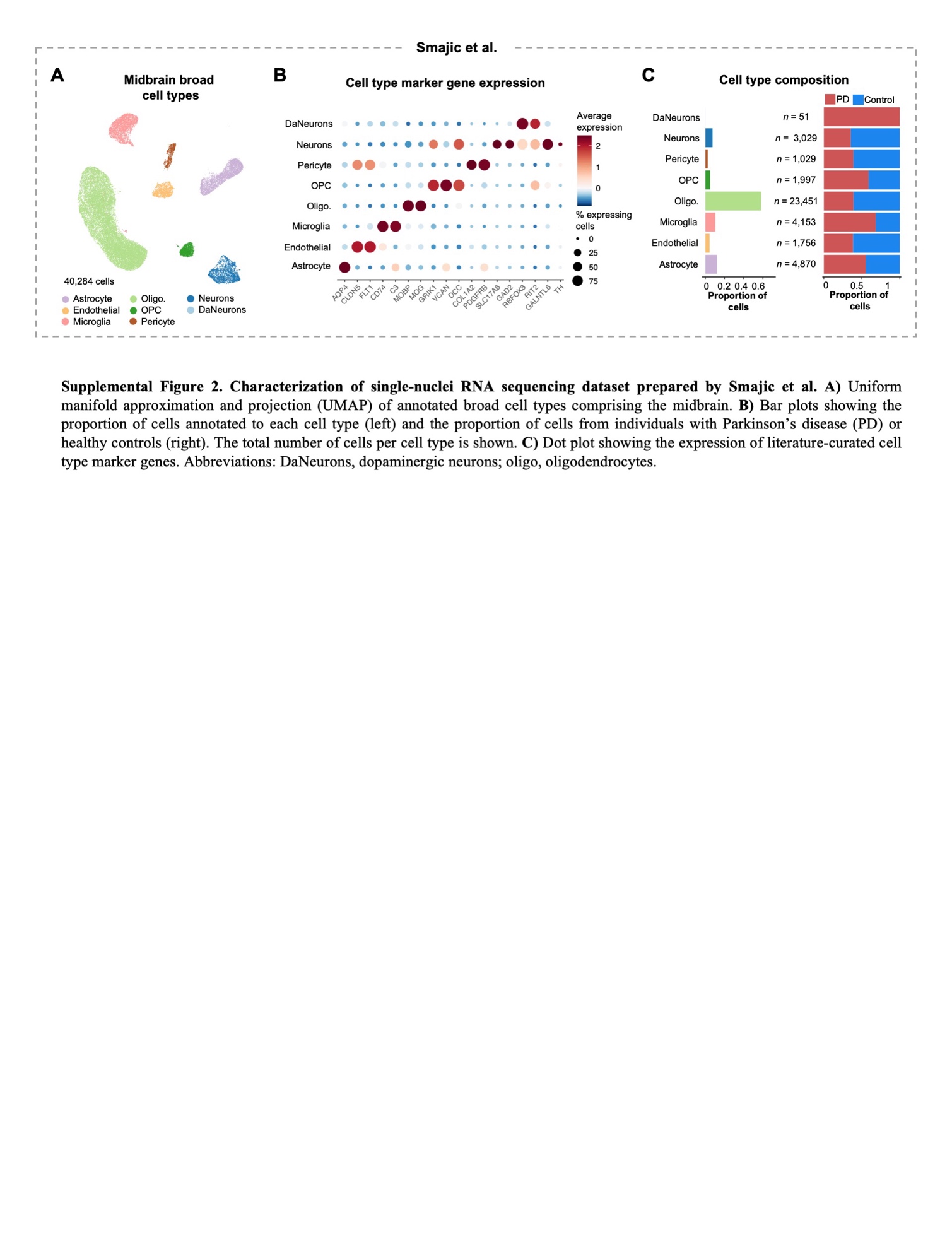

**Figure S2. Characterization of single-nuclei RNA sequencing dataset prepared by Smajic et al. A)** Uniform manifold approximation and projection (UMAP) of annotated broad cell types comprising the midbrain. **B)** Dot plot showing the expression of literature-curated cell type marker genes. **C)** Bar plots showing the proportion of cells annotated to each cell type (left) and the proportion of cells from individuals with Parkinson’s disease (PD) or healthy controls (right). The total number of cells per cell type is shown. Abbreviations: DaNeurons, dopaminergic neurons; oligo, oligodendrocytes; OPC, oligodendrocyte precursor cells.

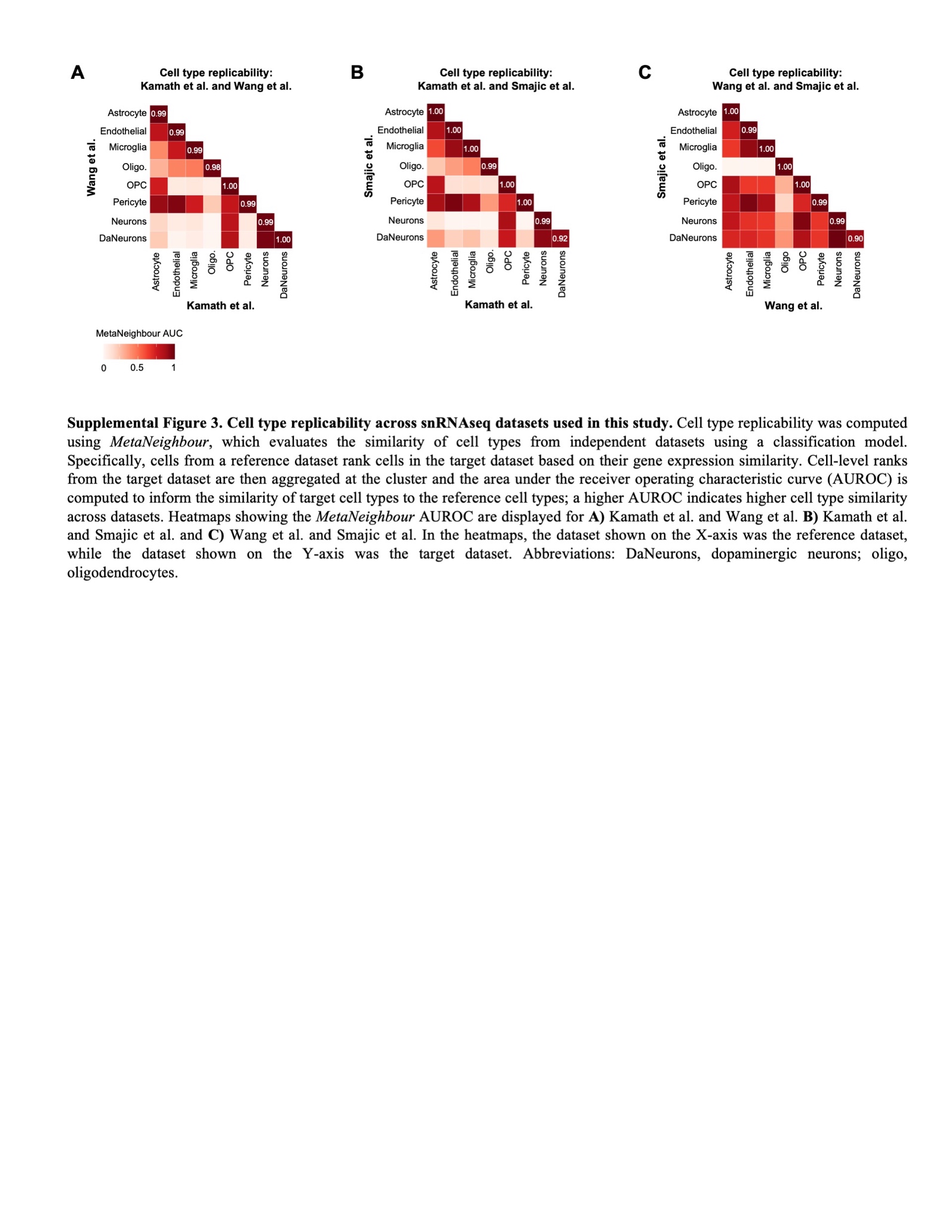

**Figure S3. Cell type replicability across snRNAseq datasets used in this study.** Cell type replicability was computed using *MetaNeighbour*, which evaluates the similarity of cell types from independent datasets using a classification model. Specifically, cells from a reference dataset rank cells in the target dataset based on their proximity in a shared gene expression space. Cell-level ranks from the target dataset are then aggregated at the cluster level and the area under the receiver operating characteristic curve (AUROC) is computed to inform the similarity of target cell types to the reference cell types. A higher AUROC indicates higher cell type similarity across datasets. Heatmaps showing the *MetaNeighbour* AUROC are displayed for **A)** Kamath et al. and Wang et al. **B)** Kamath et al. and Smajic et al. and **C)** Wang et al. and Smajic et al. In the heatmaps, the dataset shown on the X-axis was the reference dataset, while the dataset shown on the Y-axis was the target dataset. Abbreviations: DaNeurons, dopaminergic neurons; oligo, oligodendrocytes; OPC, oligodendrocyte precursor cells.

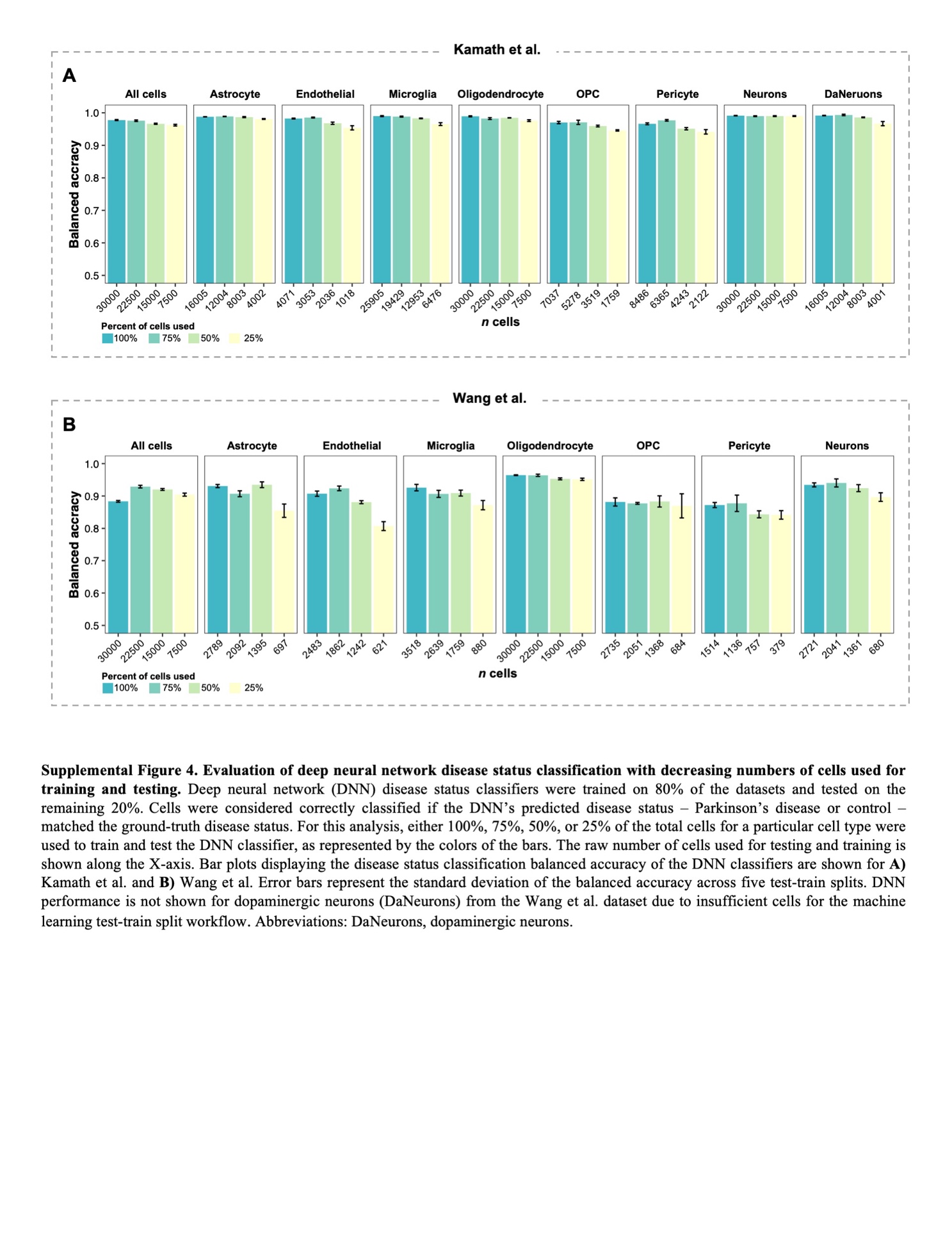

**Figure S4. Evaluation of neural network disease status classification with decreasing numbers of cells used for training and testing.** Neural network (NN) disease status classifiers were trained on 80% of the datasets and tested on the remaining 20%. Cells were considered correctly classified if the NN’s predicted disease status — Parkinson’s disease or control — matched the ground-truth disease status. For this analysis, either 100%, 75%, 50%, or 25% of the total cells for a particular cell type were used to train and test the NN classifier, as represented by the colors of the bars. The raw number of cells used for testing and training is shown along the X-axis. Bar plots displaying the disease status classification balanced accuracy of the NN classifiers following five-fold cross validation are shown for **A)** Kamath et al. and **B)** Wang et al. Error bars represent the standard deviation of the balanced accuracy across all models from five-fold cross validation. NN performance is not shown for dopaminergic neurons (DaNeurons) from the Wang et al. dataset due to insufficient cells for the machine learning test-train split workflow. Abbreviations: OPC, oligodendrocyte precursor cells.

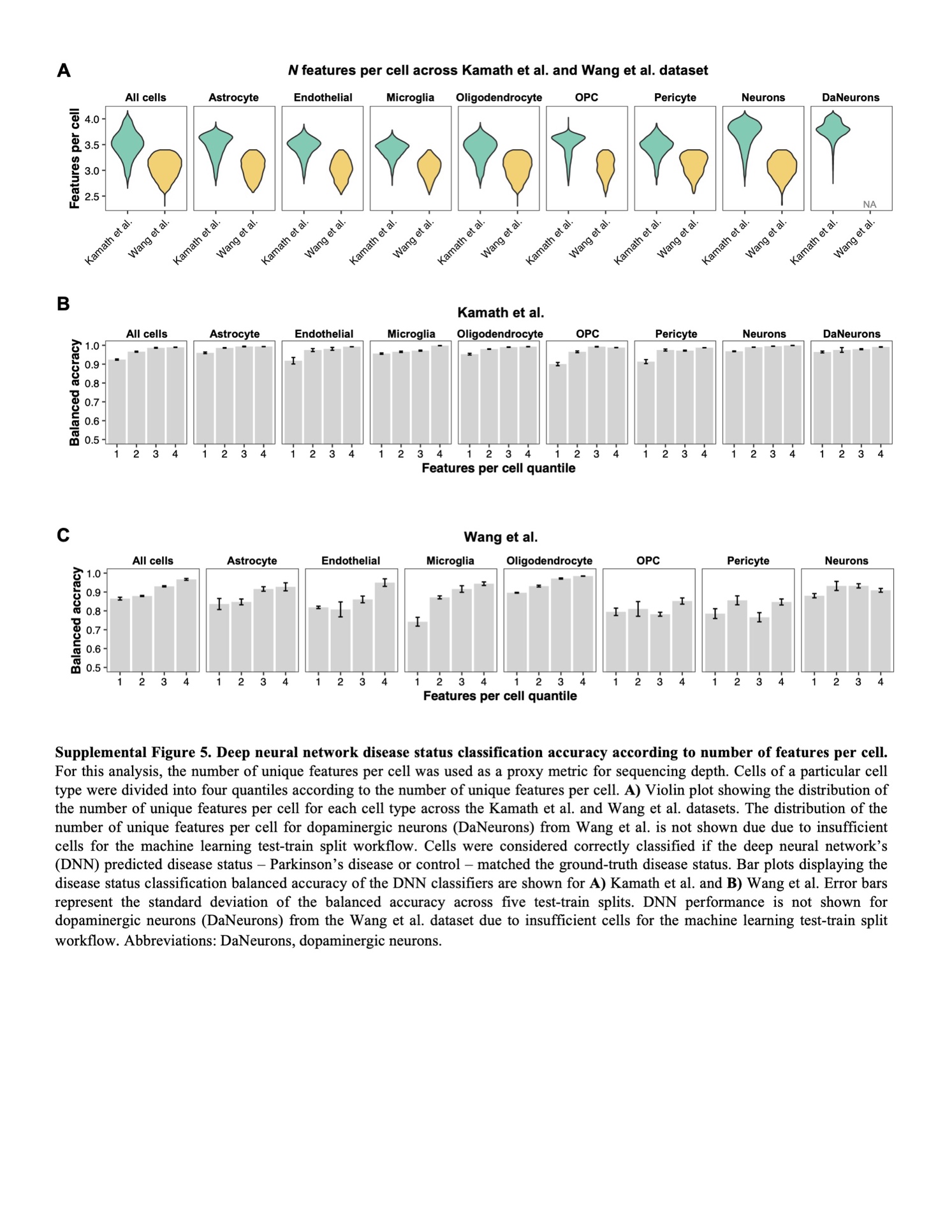

**Figure S5. Neural network disease status classification accuracy according to number of features per cell.** For this analysis, the number of unique features per cell was used as a proxy metric for sequencing depth. Cells of a particular cell type were divided into four quantiles according to the number of unique features per cell. **A)** Violin plot showing the distribution of the number of unique features per cell for each cell type across the Kamath et al. and Wang et al. datasets. The distribution of the number of unique features per cell for dopaminergic neurons (DaNeurons) from Wang et al. is not shown due to insufficient cells for the machine learning test-train split workflow. Cells were considered correctly classified if the neural network’s (NN) predicted disease status — Parkinson’s disease or control — matched the ground-truth disease status. Bar plots displaying the disease status classification balanced accuracy of the NN classifiers are shown for **A)** Kamath et al. and **B)** Wang et al. Error bars represent the standard deviation of the balanced accuracy across all models from five-fold cross validation. NN performance is not shown for DaNeurons from the Wang et al. dataset due to insufficient cells for the machine learning test-train split workflow. Abbreviations: OPC, oligodendrocyte precursor cells.

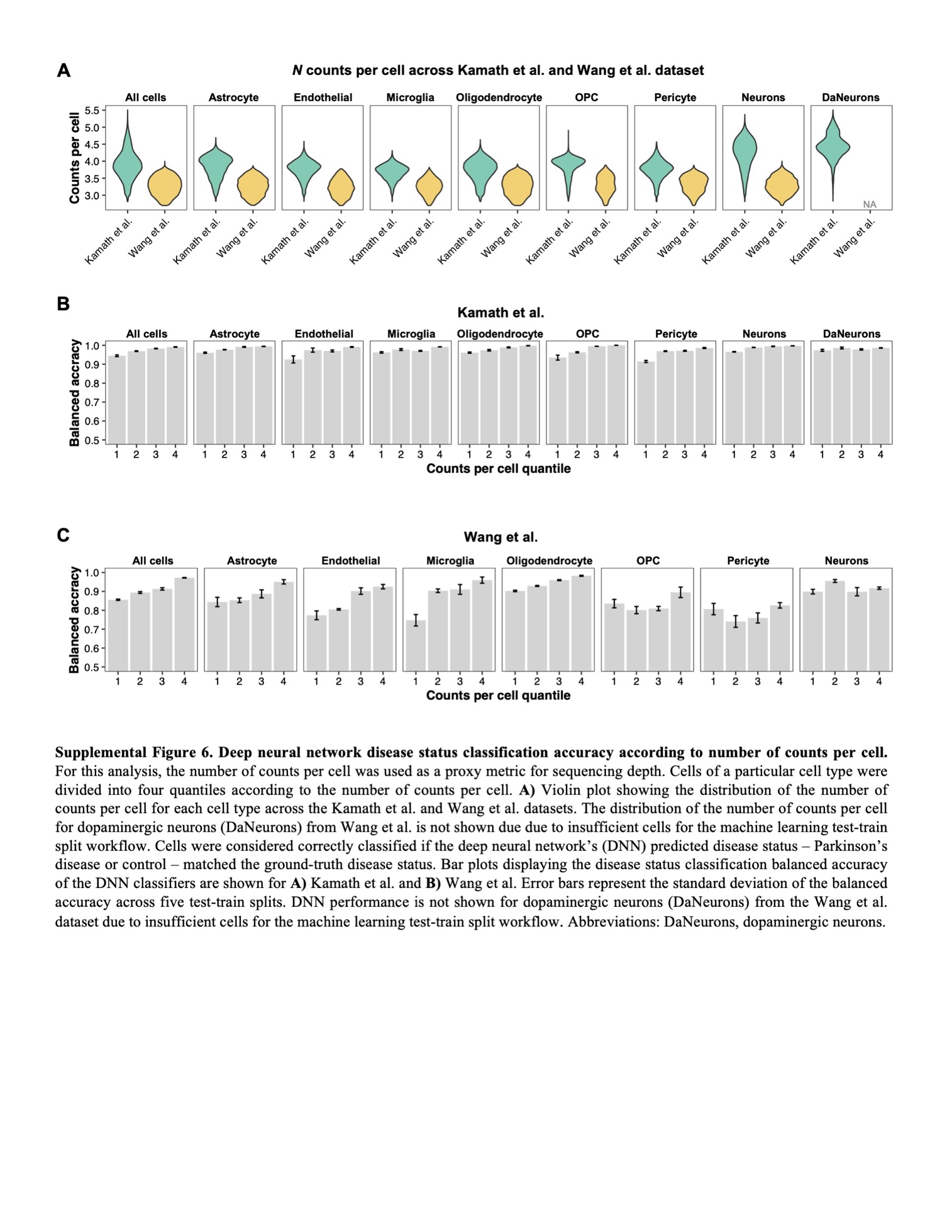

**Figure 6. Neural network disease status classification accuracy according to number of counts per cell.** For this analysis, the number of counts per cell was used as a proxy metric for sequencing depth. Cells of a particular cell type were divided into four quantiles according to the number of counts per cell. **A)** Violin plot showing the distribution of the number of counts per cell for each cell type across the Kamath et al. and Wang et al. datasets. The distribution of the number of counts per cell for dopaminergic neurons (DaNeurons) from Wang et al. is not shown due to insufficient cells for the machine learning test-train split workflow. Cells were considered correctly classified if the neural network’s (NN) predicted disease status — Parkinson’s disease or control — matched the ground-truth disease status. Bar plots displaying the disease status classification balanced accuracy of the NN classifiers are shown for **A)** Kamath et al. and **B)** Wang et al. Error bars represent the standard deviation of the balanced accuracy across all models from five-fold cross validation. NN performance is not shown for DaNeurons from the Wang et al. dataset due to insufficient cells for the machine learning test-train split workflow. Abbreviations: OPC, oligodendrocyte precursor cells.

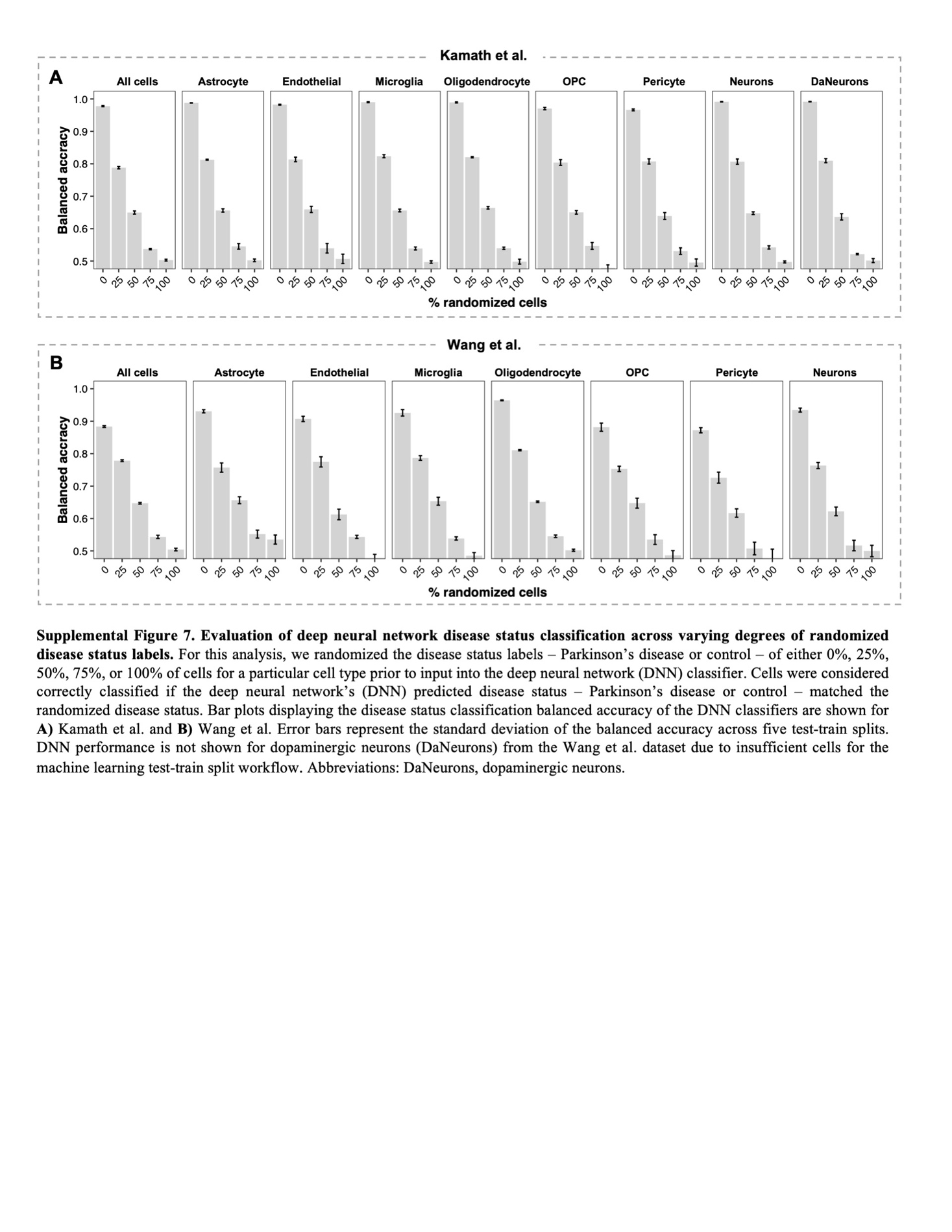

**Figure S7. Evaluation of neural network disease status classification across varying degrees of randomized disease status labels.** For this analysis, we randomized the disease status labels — Parkinson’s disease (PD) or control — of either 0%, 25%, 50%, 75%, or 100% of cells for a particular cell type prior to input into the neural network (NN) classifier. Cells were considered correctly classified if the NN’s predicted disease status — PD or control — matched the randomized disease status. Bar plots displaying the disease status classification balanced accuracy of the NN classifiers are shown for **A)** Kamath et al. and **B)** Wang et al. Error bars represent the standard deviation of the balanced accuracy across all models from five-fold cross validation. NN performance is not shown for dopaminergic neurons (DaNeurons) from the Wang et al. dataset due to insufficient cells for the machine learning test-train split workflow. Abbreviations: OPC, oligodendrocyte precursor cells.

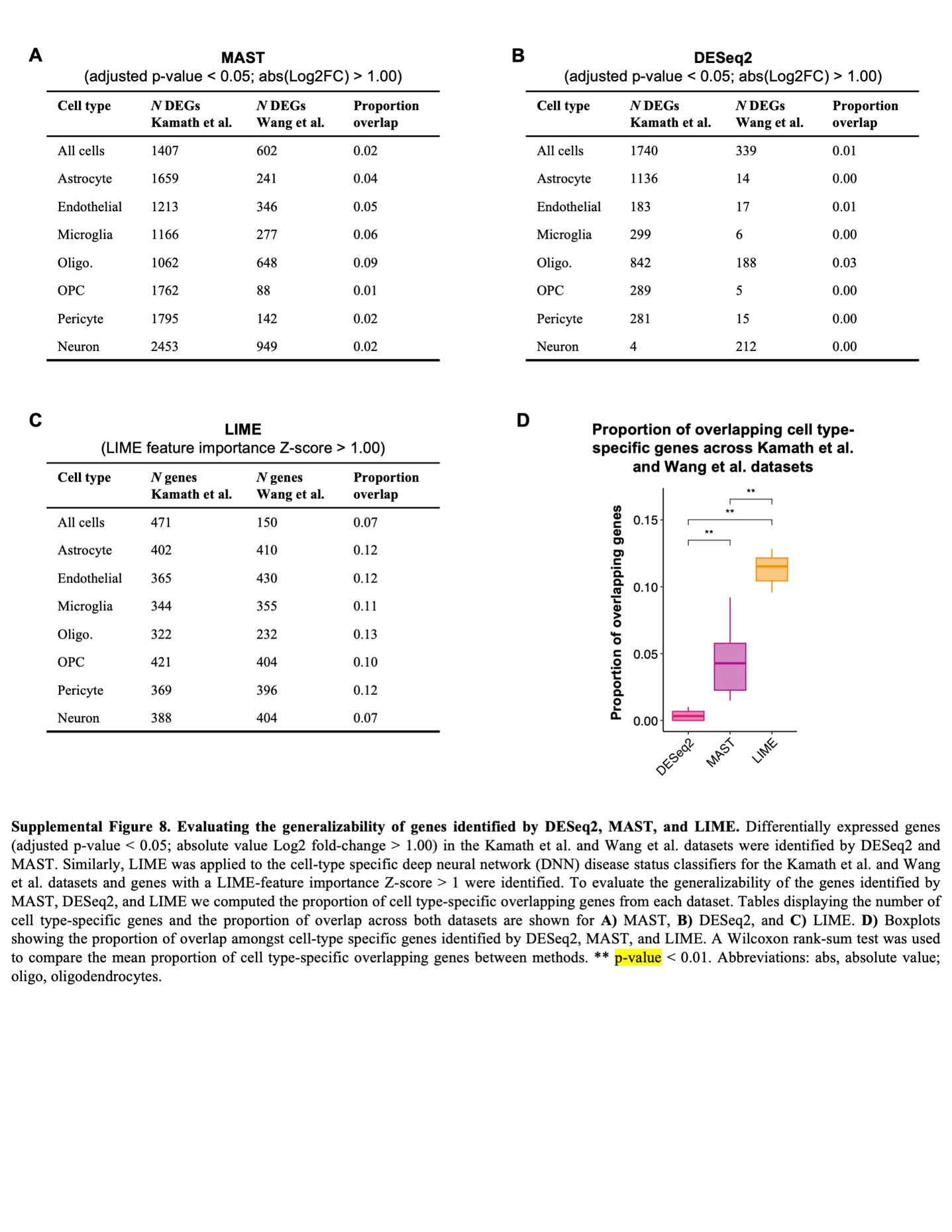

**Figure S8. Evaluating the generalizability of genes identified by DESeq2, MAST, and LIME.** Differentially expressed genes (adjusted p-value < 0.05; absolute value Log2 fold-change > 1.00) in the Kamath et al. and Wang et al. datasets were identified by DESeq2 and MAST. Similarly, LIME was applied to the cell-type specific neural network (NN) disease status classifiers for the Kamath et al. and Wang et al. datasets and genes with a LIME-feature importance Z-score > 1 were identified. To evaluate the generalizability of the genes identified by MAST, DESeq2, and LIME, we computed the proportion of cell type-specific overlapping genes from each dataset. Tables displaying the number of cell type-specific genes and the proportion of overlap across both datasets are shown for **A)** MAST, **B)** DESeq2, and **C)** LIME. **D)** Boxplots showing the proportion of overlap amongst cell-type specific genes identified by DESeq2, MAST, and LIME. A Wilcoxon rank-sum test was used to compare the mean proportion of cell type-specific overlapping genes between methods. ** p-value < 0.01. Abbreviations: abs, absolute value; oligo, oligodendrocytes; OPC, oligodendrocyte precursor cells.

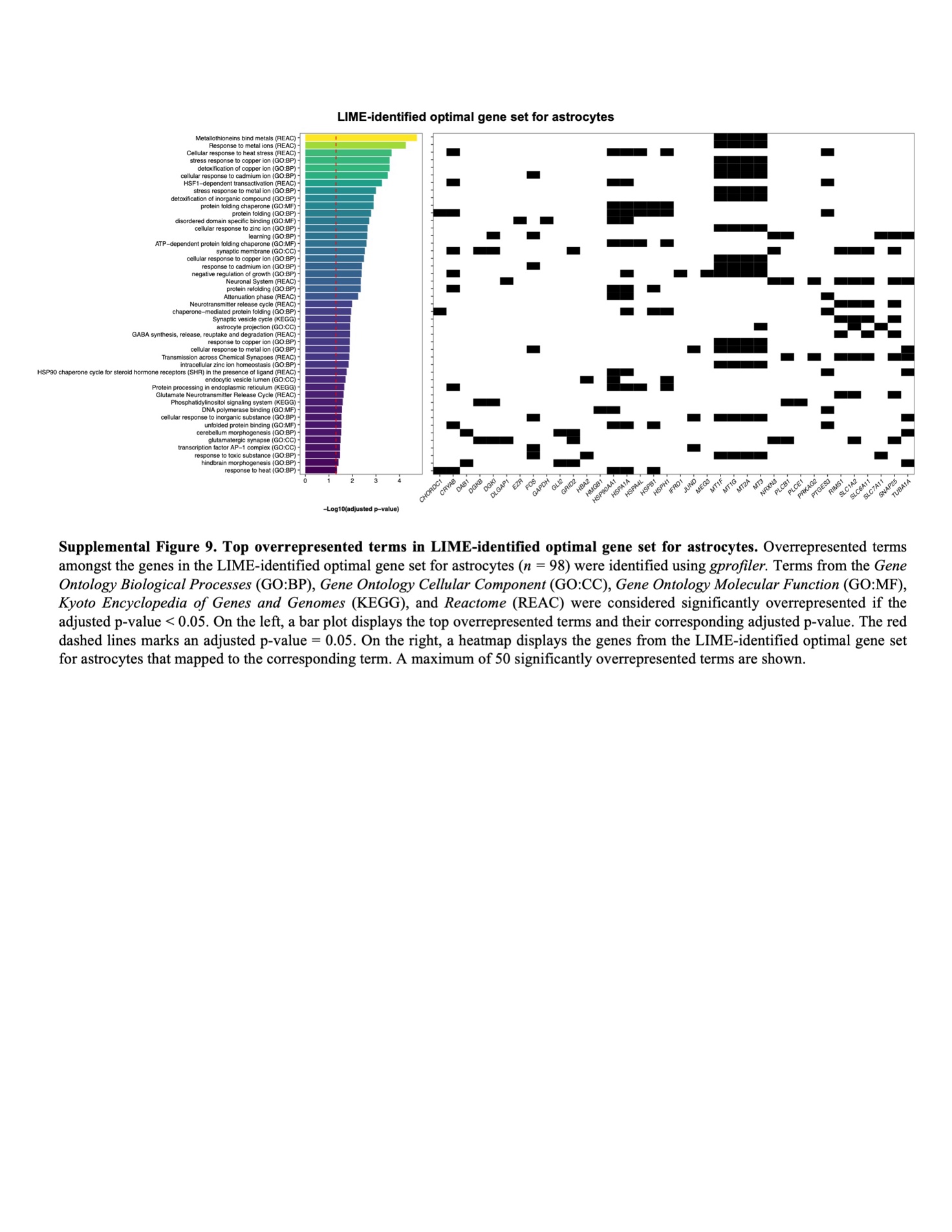

**Figure S9. Top overrepresented terms in LIME-identified optimal gene set for astrocytes.** Overrepresented terms amongst the genes in the LIME-identified optimal gene set for astrocytes (*n =* 98) were identified using *gprofiler.* Terms from the *Gene Ontology Biological Processes* (GO:BP), *Gene Ontology Cellular Component* (GO:CC), *Gene Ontology Molecular Function* (GO:MF), *Kyoto Encyclopedia of Genes and Genomes* (KEGG), and *Reactome* (REAC) were considered significantly overrepresented if the adjusted p-value was < 0.05. On the left, a bar plot displays the top overrepresented terms and their corresponding adjusted p-value. The red dashed lines marks an adjusted p-value = 0.05. On the right, a heatmap displays the genes from the LIME-identified optimal gene set for astrocytes that mapped to the corresponding term. A maximum of 50 significantly overrepresented terms are shown.

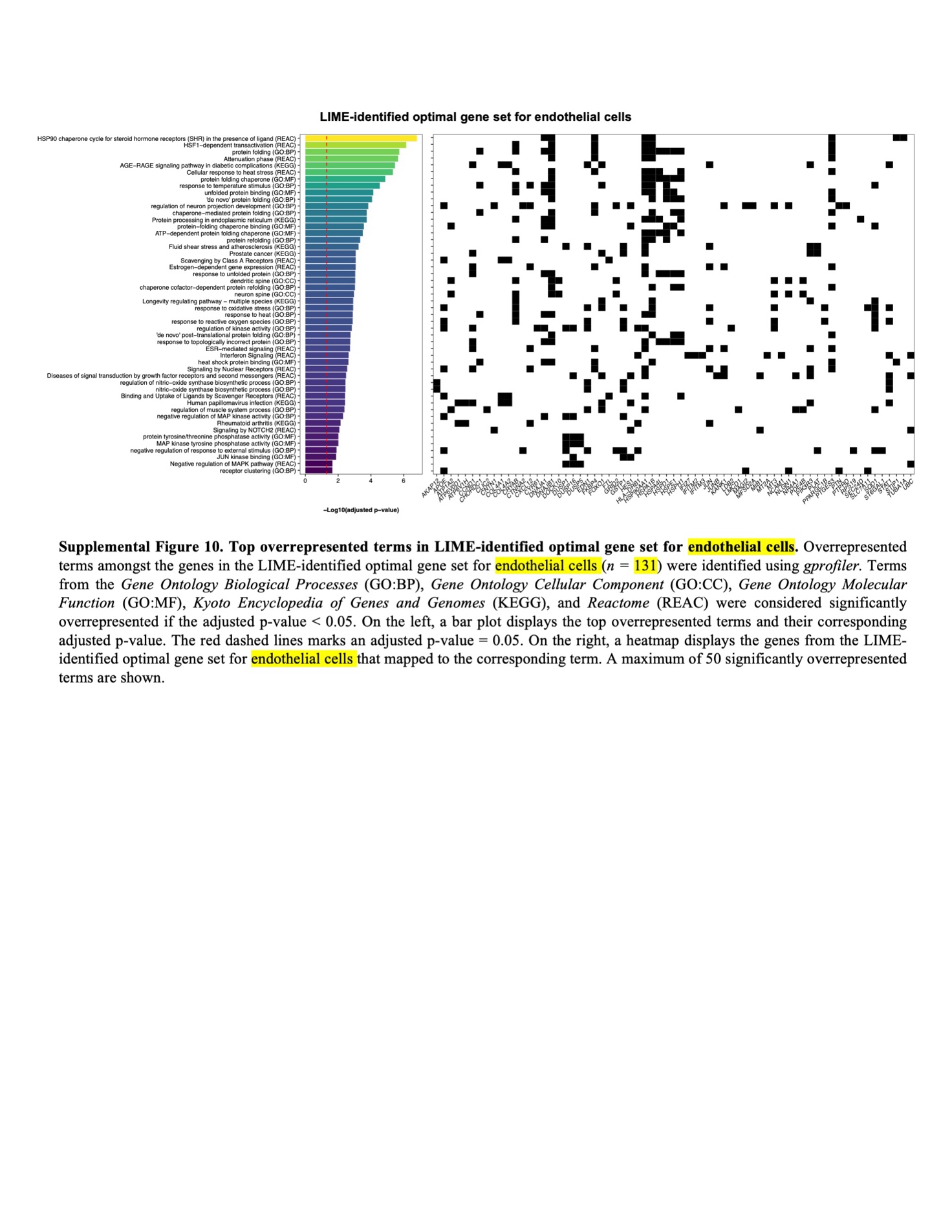

**Figure S10. Top overrepresented terms in LIME-identified optimal gene set for endothelial cells.** Overrepresented terms amongst the genes in the LIME-identified optimal gene set for endothelial cells (*n =* 131) were identified using *gprofiler.* Terms from the *Gene Ontology Biological Processes* (GO:BP), *Gene Ontology Cellular Component* (GO:CC), *Gene Ontology Molecular Function* (GO:MF), *Kyoto Encyclopedia of Genes and Genomes* (KEGG), and *Reactome* (REAC) were considered significantly overrepresented if the adjusted p-value was < 0.05. On the left, a bar plot displays the top overrepresented terms and their corresponding adjusted p-value. The red dashed lines marks an adjusted p-value = 0.05. On the right, a heatmap displays the genes from the LIME-identified optimal gene set for endothelial cells that mapped to the corresponding term. A maximum of 50 significantly overrepresented terms are shown.

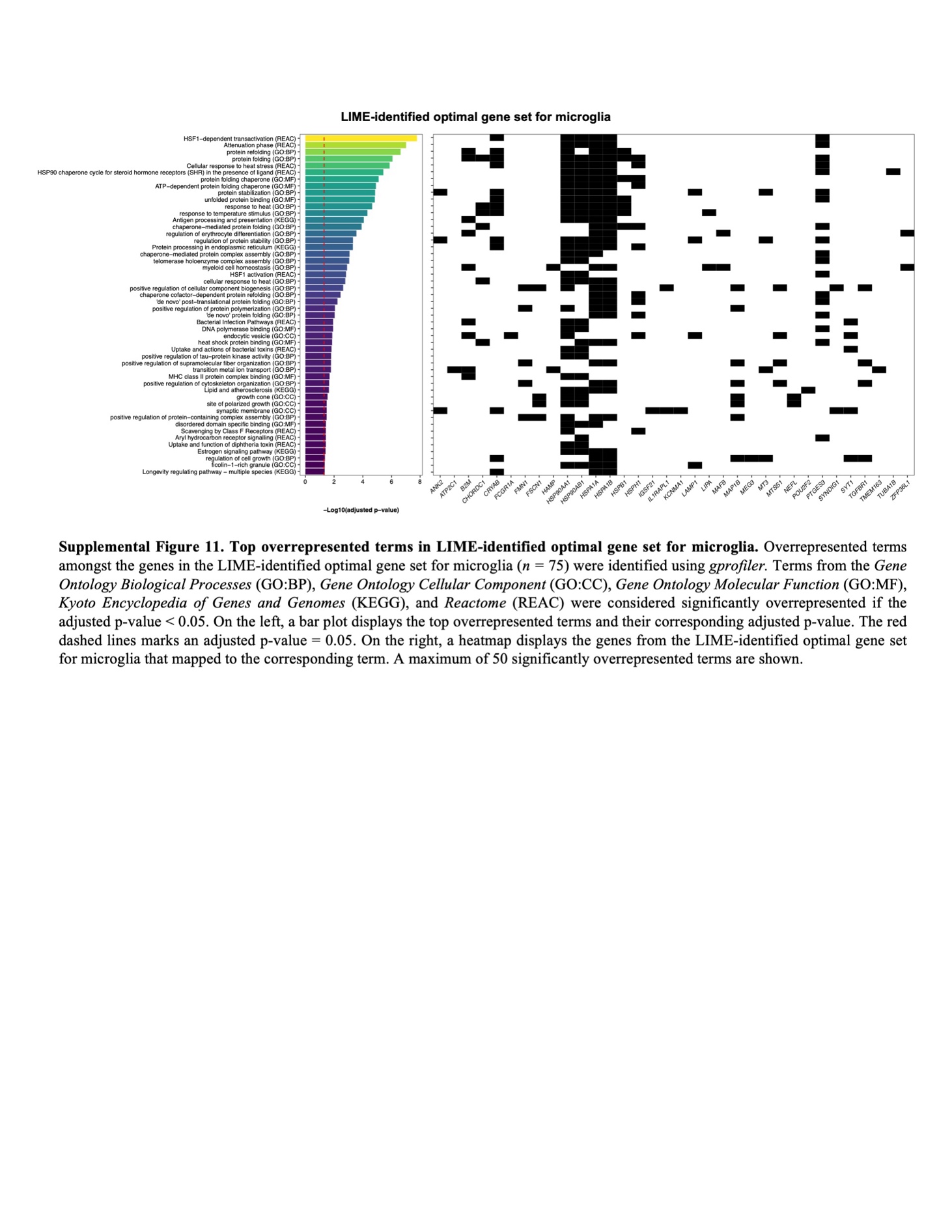

**Figure S11. Top overrepresented terms in LIME-identified optimal gene set for microglia.** Overrepresented terms amongst the genes in the LIME-identified optimal gene set for microglia (*n =* 75) were identified using *gprofiler.* Terms from the *Gene Ontology Biological Processes* (GO:BP), *Gene Ontology Cellular Component* (GO:CC), *Gene Ontology Molecular Function* (GO:MF), *Kyoto Encyclopedia of Genes and Genomes* (KEGG), and *Reactome* (REAC) were considered significantly overrepresented if the adjusted p-value was < 0.05. On the left, a bar plot displays the top overrepresented terms and their corresponding adjusted p-value. The red dashed lines marks an adjusted p-value = 0.05. On the right, a heatmap displays the genes from the LIME-identified optimal gene set for microglia that mapped to the corresponding term. A maximum of 50 significantly overrepresented terms are shown.

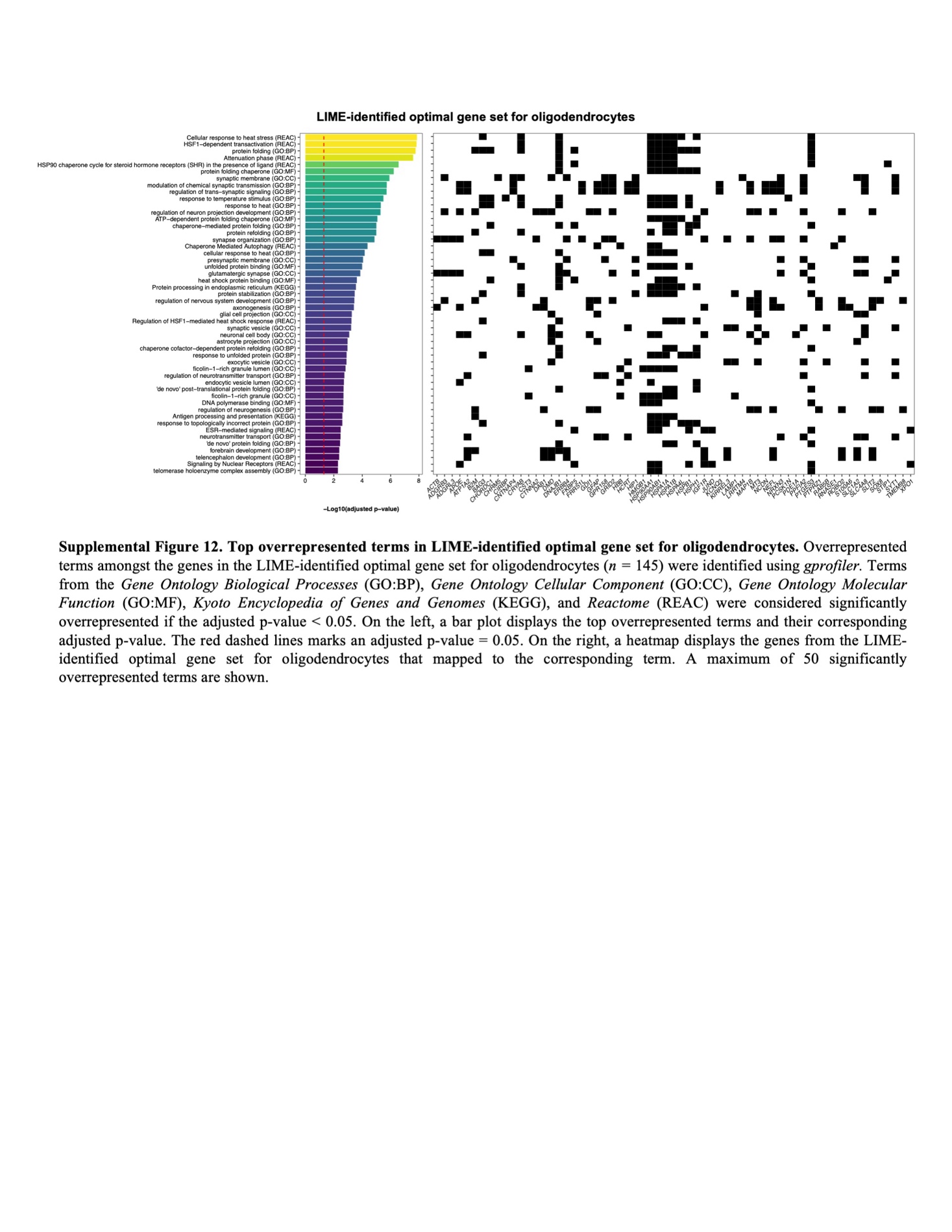

**Figure S12. Top overrepresented terms in LIME-identified optimal gene set for oligodendrocytes.** Overrepresented terms amongst the genes in the LIME-identified optimal gene set for oligodendrocytes (*n =* 145) were identified using *gprofiler.* Terms from the *Gene Ontology Biological Processes* (GO:BP), *Gene Ontology Cellular Component* (GO:CC), *Gene Ontology Molecular Function* (GO:MF), *Kyoto Encyclopedia of Genes and Genomes* (KEGG), and *Reactome* (REAC) were considered significantly overrepresented if the adjusted p-value was < 0.05. On the left, a bar plot displays the top overrepresented terms and their corresponding adjusted p-value. The red dashed lines marks an adjusted p-value = 0.05. On the right, a heatmap displays the genes from the LIME-identified optimal gene set for oligodendrocytes that mapped to the corresponding term. A maximum of 50 significantly overrepresented terms are shown.

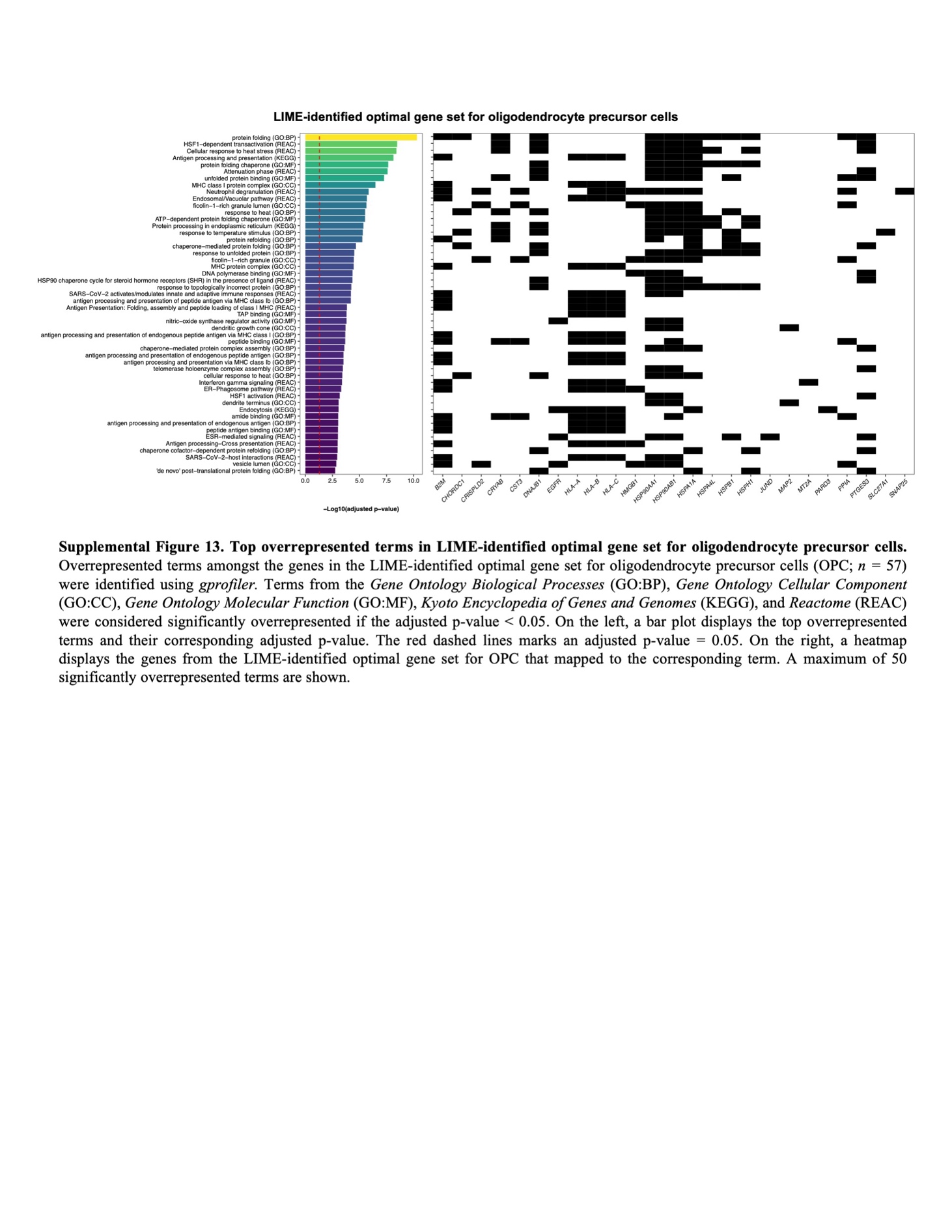

**Figure S13. Top overrepresented terms in LIME-identified optimal gene set for oligodendrocyte precursor cells.** Overrepresented terms amongst the genes in the LIME-identified optimal gene set for oligodendrocyte precursor cells (OPC; *n =* 57) were identified using *gprofiler.* Terms from the *Gene Ontology Biological Processes* (GO:BP), *Gene Ontology Cellular Component* (GO:CC), *Gene Ontology Molecular Function* (GO:MF), *Kyoto Encyclopedia of Genes and Genomes* (KEGG), and *Reactome* (REAC) were considered significantly overrepresented if the adjusted p-value was < 0.05. On the left, a bar plot displays the top overrepresented terms and their corresponding adjusted p-value. The red dashed lines marks an adjusted p-value = 0.05. On the right, a heatmap displays the genes from the LIME-identified optimal gene set for OPC that mapped to the corresponding term. A maximum of 50 significantly overrepresented terms are shown.

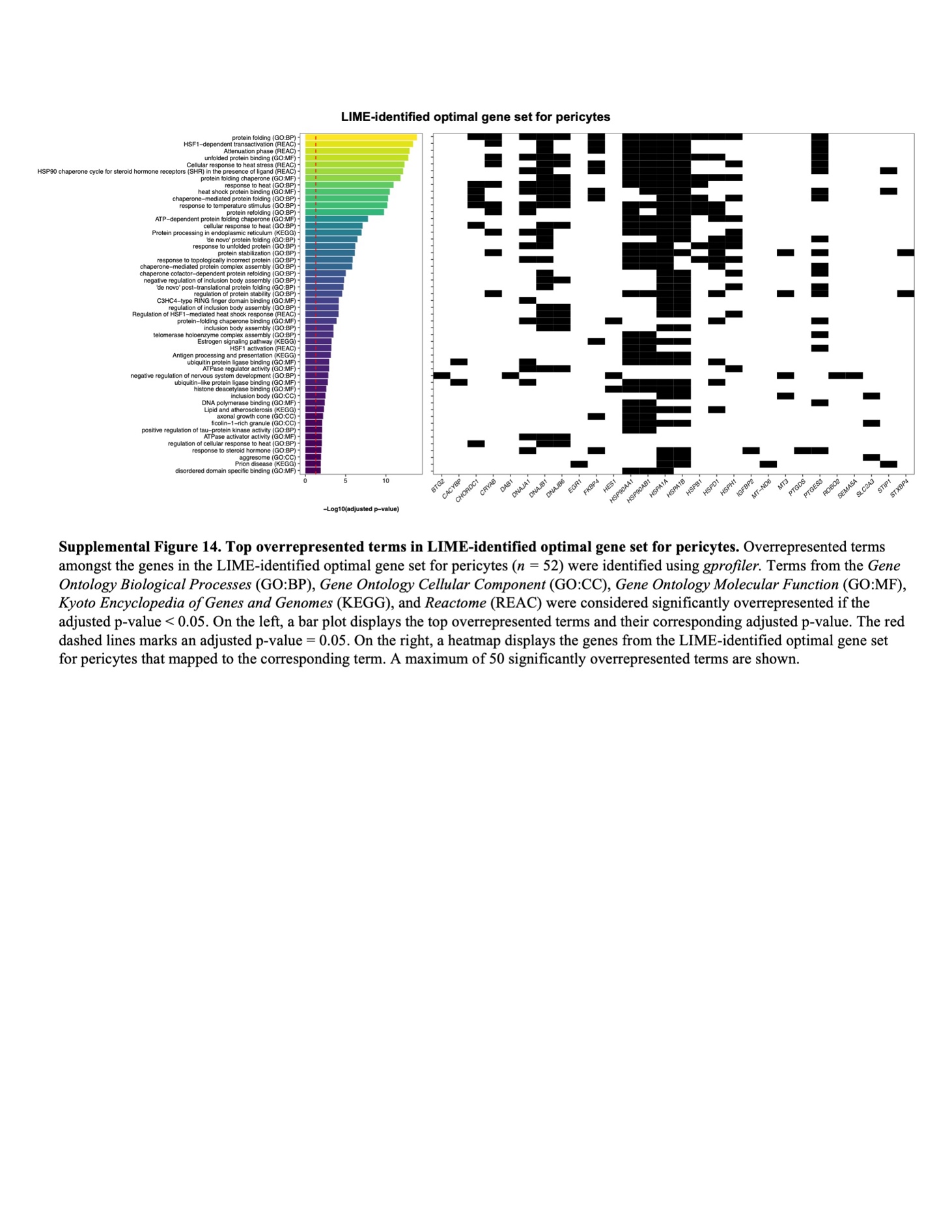

**Figure S14. Top overrepresented terms in LIME-identified optimal gene set for pericytes.** Overrepresented terms amongst the genes in the LIME-identified optimal gene set for pericytes (*n =* 52) were identified using *gprofiler.* Terms from the *Gene Ontology Biological Processes* (GO:BP), *Gene Ontology Cellular Component* (GO:CC), *Gene Ontology Molecular Function* (GO:MF), *Kyoto Encyclopedia of Genes and Genomes* (KEGG), and *Reactome* (REAC) were considered significantly overrepresented if the adjusted p-value was < 0.05. On the left, a bar plot displays the top overrepresented terms and their corresponding adjusted p-value. The red dashed lines marks an adjusted p-value = 0.05. On the right, a heatmap displays the genes from the LIME-identified optimal gene set for pericytes that mapped to the corresponding term. A maximum of 50 significantly overrepresented terms are shown.

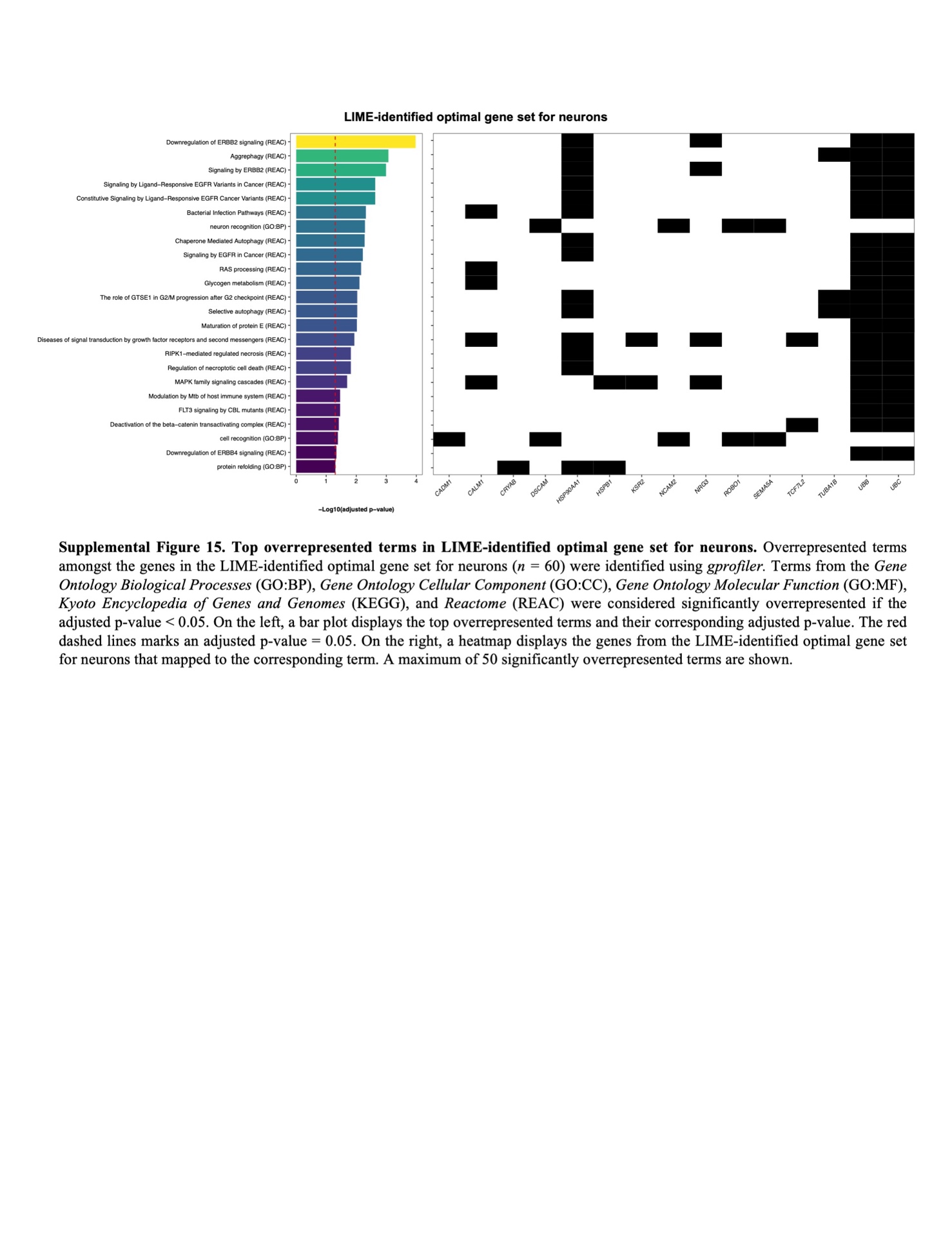

**Figure S15. Top overrepresented terms in LIME-identified optimal gene set for neurons.** Overrepresented terms amongst the genes in the LIME-identified optimal gene set for neurons (*n =* 60) were identified using *gprofiler.* Terms from the *Gene Ontology Biological Processes* (GO:BP), *Gene Ontology Cellular Component* (GO:CC), *Gene Ontology Molecular Function* (GO:MF), *Kyoto Encyclopedia of Genes and Genomes* (KEGG), and *Reactome* (REAC) were considered significantly overrepresented if the adjusted p-value was < 0.05. On the left, a bar plot displays the top overrepresented terms and their corresponding adjusted p-value. The red dashed lines marks an adjusted p-value = 0.05. On the right, a heatmap displays the genes from the LIME-identified optimal gene set for neurons that mapped to the corresponding term. A maximum of 50 significantly overrepresented terms are shown.

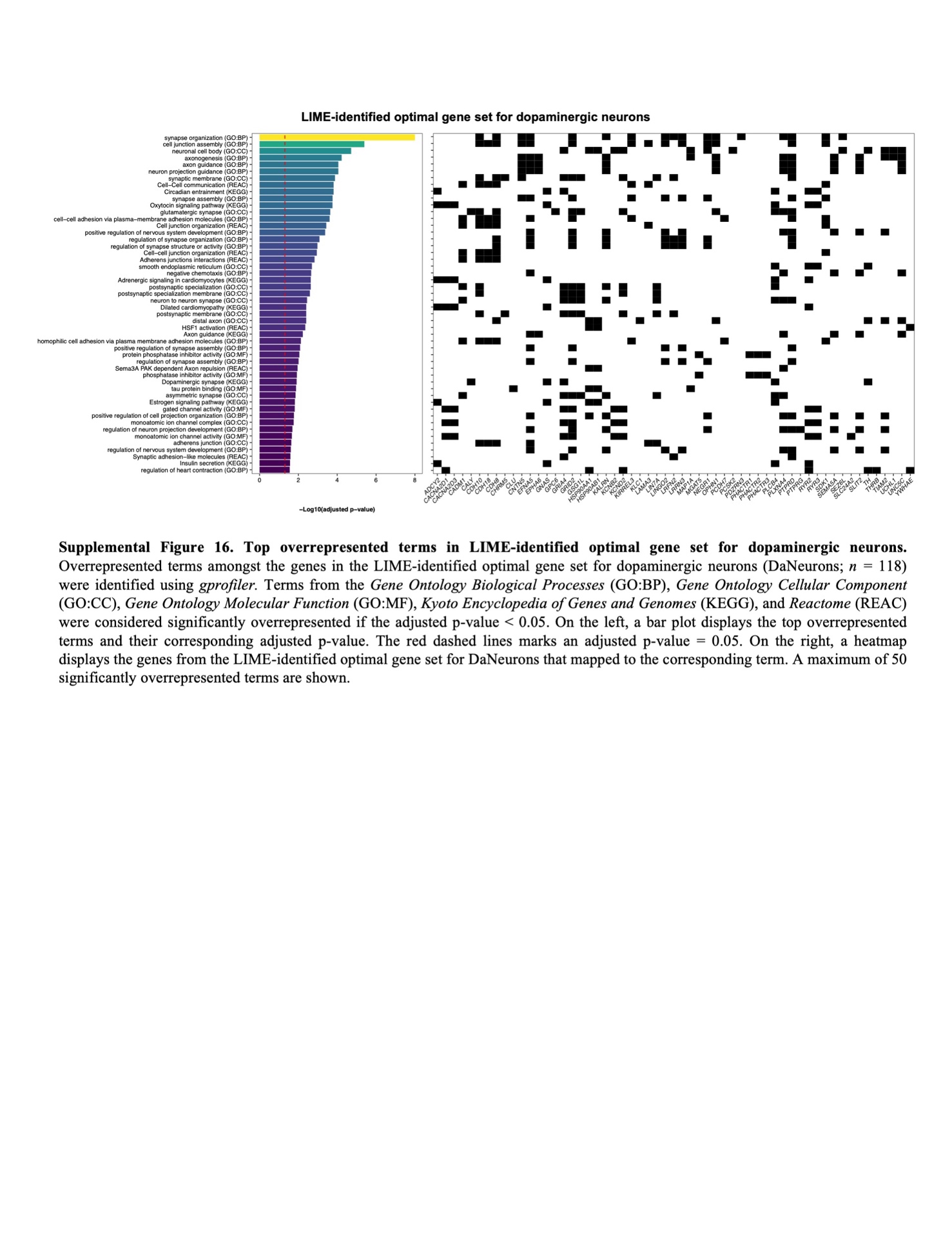

**Figure S16. Top overrepresented terms in LIME-identified optimal gene set for dopaminergic neurons.** Overrepresented terms amongst the genes in the LIME-identified optimal gene set for dopaminergic neurons (DaNeurons; *n =* 118) were identified using *gprofiler.* Terms from the *Gene Ontology Biological Processes* (GO:BP), *Gene Ontology Cellular Component* (GO:CC), *Gene Ontology Molecular Function* (GO:MF), *Kyoto Encyclopedia of Genes and Genomes* (KEGG), and *Reactome* (REAC) were considered significantly overrepresented if the adjusted p-value was < 0.05. On the left, a bar plot displays the top overrepresented terms and their corresponding adjusted p-value. The red dashed lines marks an adjusted p-value = 0.05. On the right, a heatmap displays the genes from the LIME-identified optimal gene set for DaNeurons that mapped to the corresponding term. A maximum of 50 significantly overrepresented terms are shown.

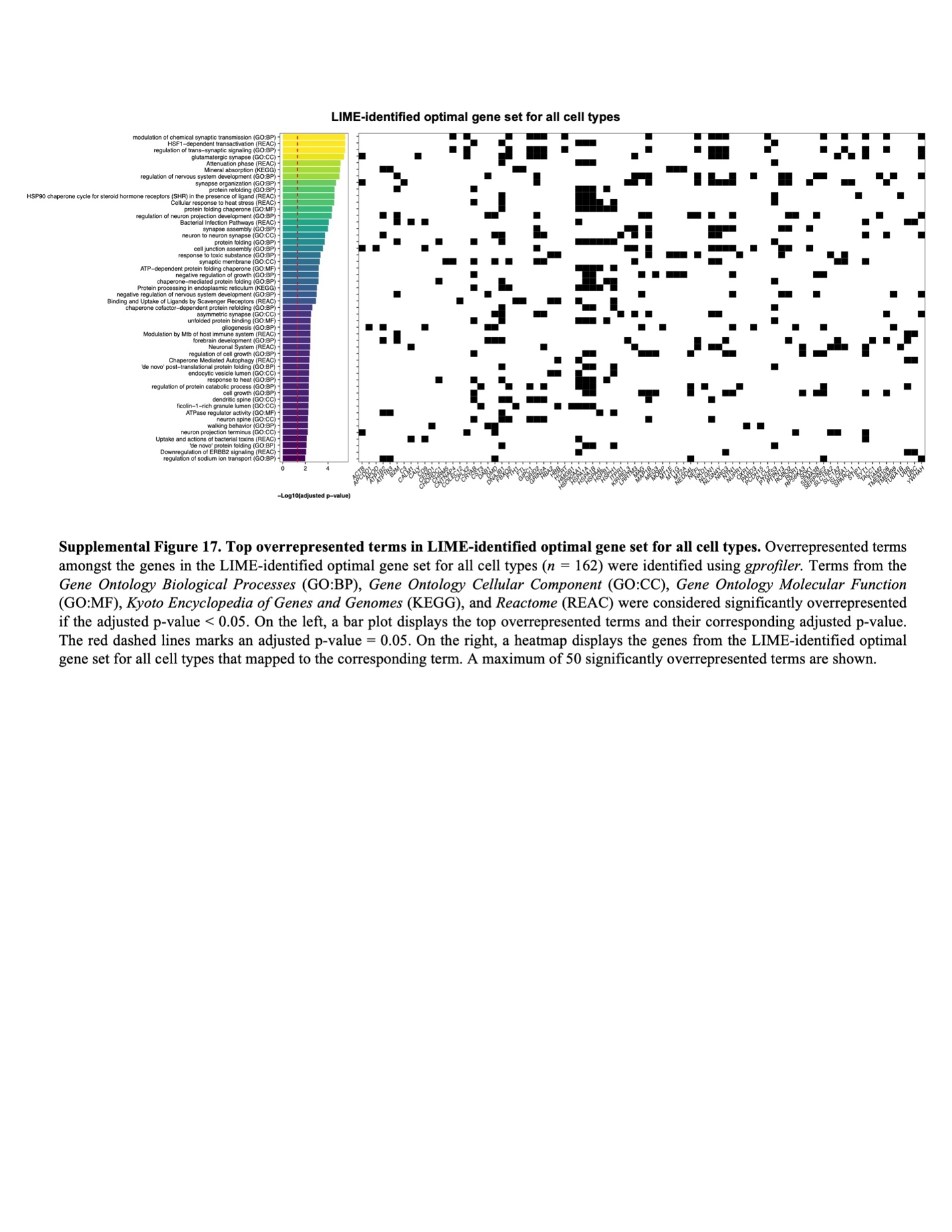

**Figure S17. Top overrepresented terms in LIME-identified optimal gene set for all cell types.** Overrepresented terms amongst the genes in the LIME-identified optimal gene set for all cell types (*n =* 162) were identified using *gprofiler.* Terms from the *Gene Ontology Biological Processes* (GO:BP), *Gene Ontology Cellular Component* (GO:CC), *Gene Ontology Molecular Function* (GO:MF), *Kyoto Encyclopedia of Genes and Genomes* (KEGG), and *Reactome* (REAC) were considered significantly overrepresented if the adjusted p-value was < 0.05. On the left, a bar plot displays the top overrepresented terms and their corresponding adjusted p-value. The red dashed lines marks an adjusted p-value = 0.05. On the right, a heatmap displays the genes from the LIME-identified optimal gene set for all cell types that mapped to the corresponding term. A maximum of 50 significantly overrepresented terms are shown.

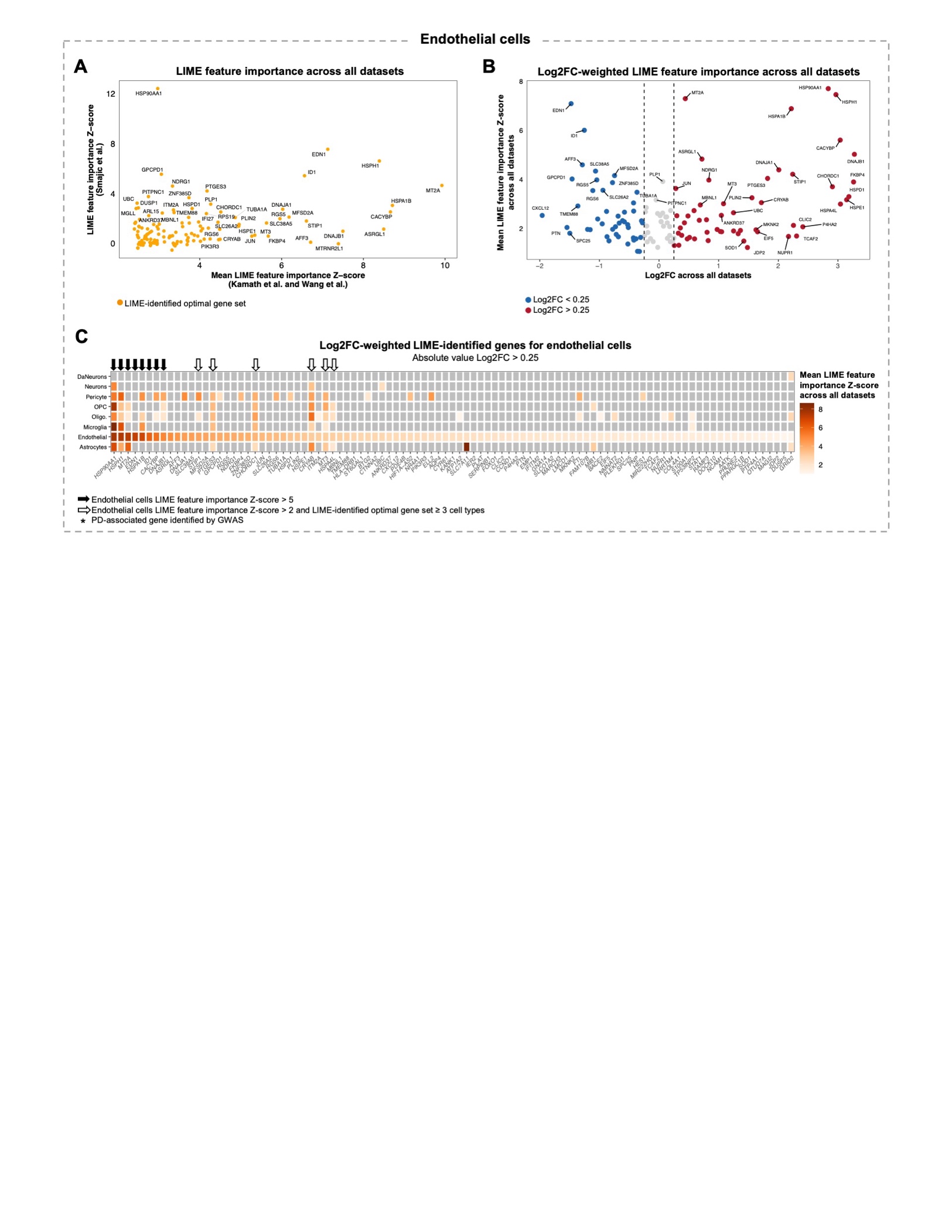

**Figure S18. Prioritization of LIME-identified genes for characterizing Parkinson’s disease endothelial cells. A)** Scatter plot showing the LIME feature importance Z-score for genes in the LIME-identified optimal gene set for endothelial cells. **B)** Volcano plot showing the mean LIME feature importance Z-score across the Kamath et al., Wang et al., and Smajic et al. datasets (Y-axis) and the gene expression log2 fold-change (Log2FC) between PD and control endothelial cells across all datasets (X-axis). Only genes comprising the LIME-identified optimal gene set for endothelial cells are shown. **C)** Heatmap showing the mean LIME feature importance Z-score for genes comprising the endothelial cells LIME-identified optimal gene set with an absolute value Log2FC > 0.25. In the heatmap, grey tiles indicate that the gene was not included in the cell type-specific optimal gene set. Abbreviations: DaNeurons, dopaminergic neurons; oligo, oligodendrocytes; OPC, oligodendrocyte precursor cells.

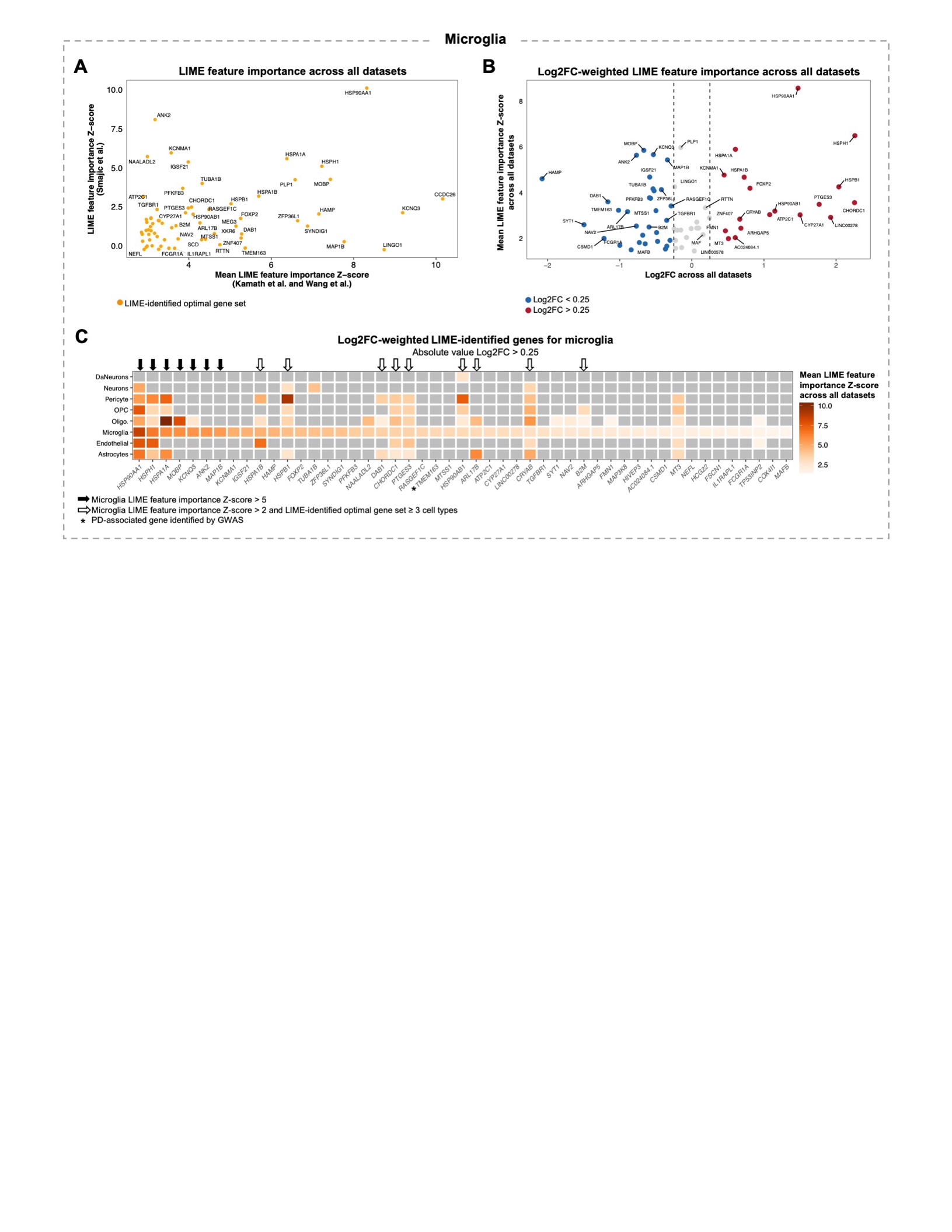

**Figure S19. Prioritization of LIME-identified genes for characterizing Parkinson’s disease microglia. A)** Scatter plot showing the LIME feature importance Z-score for genes in the LIME-identified optimal gene set for microglia. **B)** Volcano plot showing the mean LIME feature importance Z-score across the Kamath et al., Wang et al., and Smajic et al. datasets (Y-axis) and the gene expression log2 fold-change (Log2FC) between PD and control microglia across all datasets (X-axis). Only genes comprising the LIME-identified optimal gene set for microglia are shown. **C)** Heatmap showing the mean LIME feature importance Z-score for genes comprising the microglia LIME-identified optimal gene set with an absolute value Log2FC > 0.25. In the heatmap, grey tiles indicate that the gene was not included in the cell type-specific optimal gene set. Abbreviations: DaNeurons, dopaminergic neurons; oligo, oligodendrocytes; OPC, oligodendrocyte precursor cells.

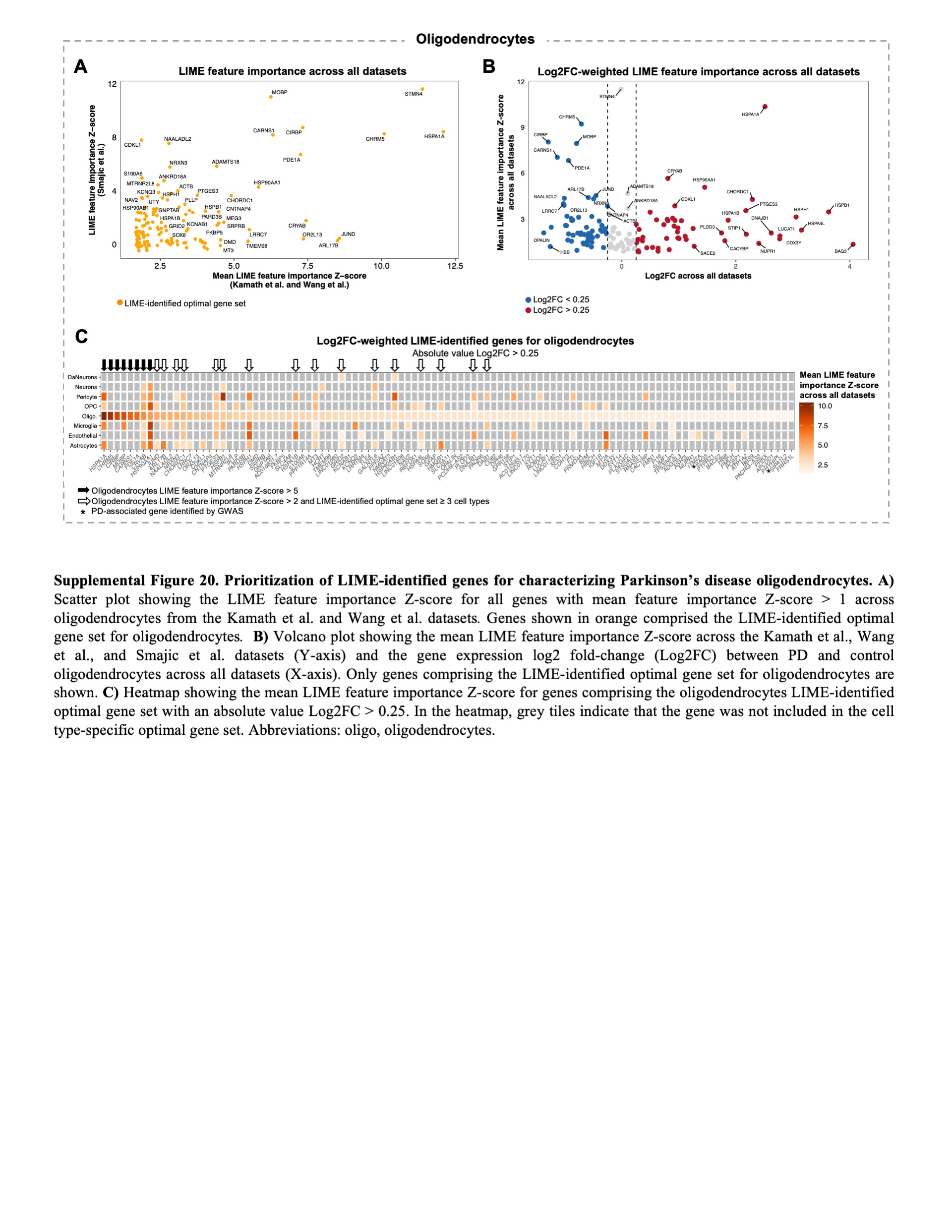

**Figure S20. Prioritization of LIME-identified genes for characterizing Parkinson’s disease oligodendrocytes. A)** Scatter plot showing the LIME feature importance Z-score for genes in the LIME-identified optimal gene set for oligodendrocytes. **B)** Volcano plot showing the mean LIME feature importance Z-score across the Kamath et al., Wang et al., and Smajic et al. datasets (Y-axis) and the gene expression log2 fold-change (Log2FC) between PD and control oligodendrocytes across all datasets (X-axis). Only genes comprising the LIME-identified optimal gene set for oligodendrocytes are shown. **C)** Heatmap showing the mean LIME feature importance Z-score for genes comprising the oligodendrocytes LIME-identified optimal gene set with an absolute value Log2FC > 0.25. In the heatmap, grey tiles indicate that the gene was not included in the cell type-specific optimal gene set. Abbreviations: DaNeurons, dopaminergic neurons; oligo, oligodendrocytes; OPC, oligodendrocyte precursor cells.

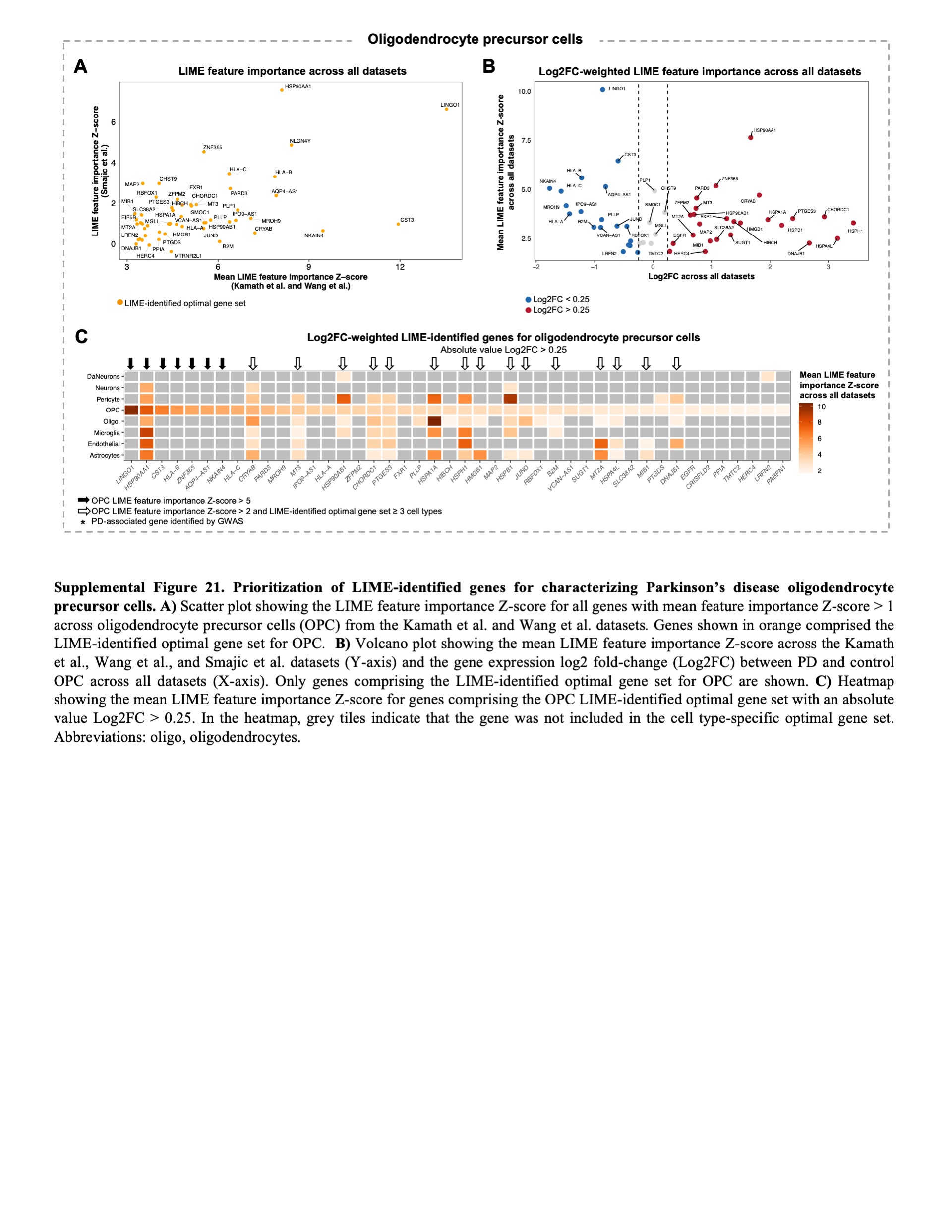

**Figure S21. Prioritization of LIME-identified genes for characterizing Parkinson’s disease oligodendrocyte precursor cells. A)** Scatter plot showing the LIME feature importance Z-score for genes in the LIME-identified optimal gene set for oligodendrocyte precursor cells (OPC). **B)** Volcano plot showing the mean LIME feature importance Z-score across the Kamath et al., Wang et al., and Smajic et al. datasets (Y-axis) and the gene expression log2 fold-change (Log2FC) between PD and control OPCs across all datasets (X-axis). Only genes comprising the LIME-identified optimal gene set for OPCs are shown. **C)** Heatmap showing the mean LIME feature importance Z-score for genes comprising the OPCs LIME-identified optimal gene set with an absolute value Log2FC > 0.25. In the heatmap, grey tiles indicate that the gene was not included in the cell type-specific optimal gene set. Abbreviations: DaNeurons, dopaminergic neurons; oligo, oligodendrocytes.

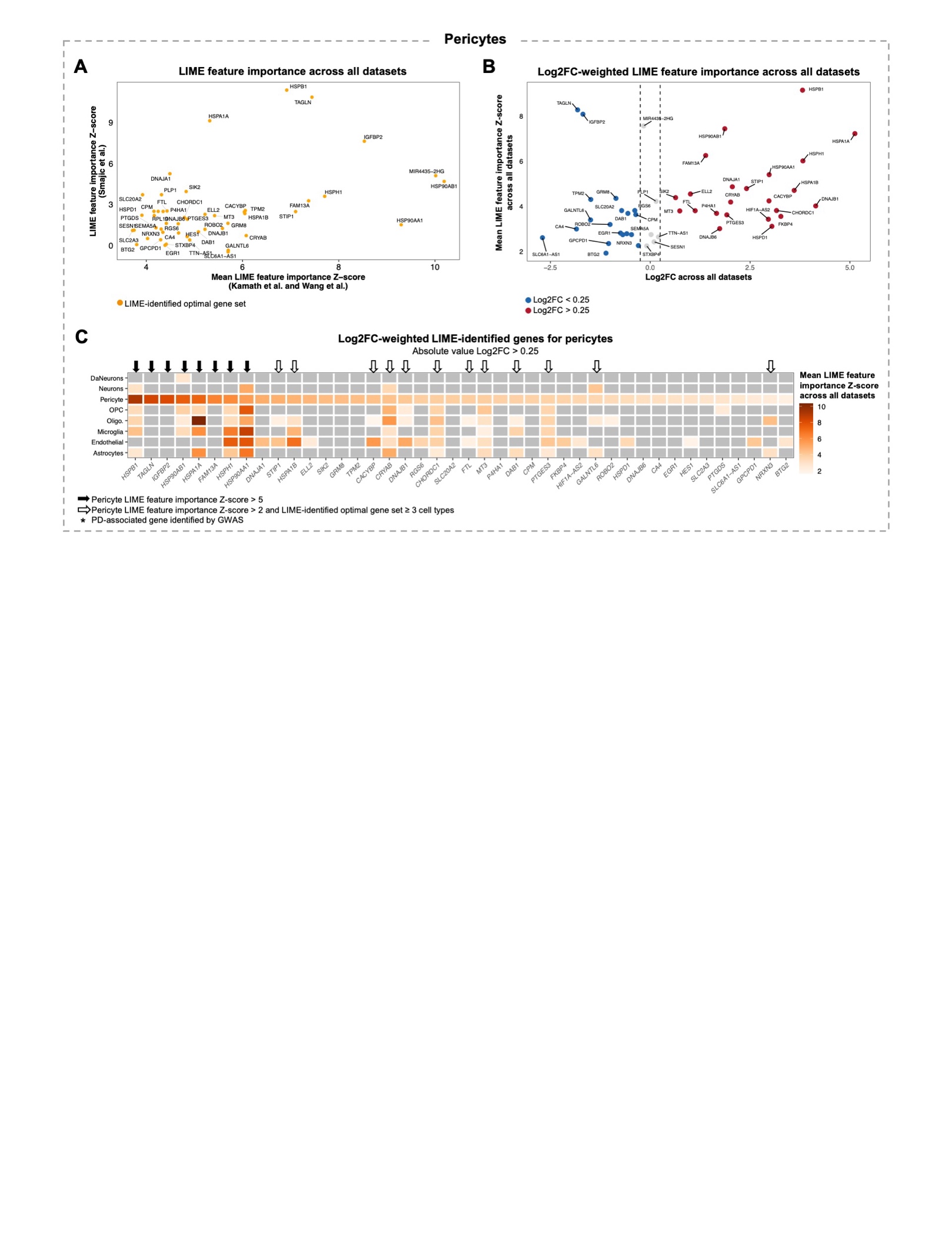

**Figure S22. Prioritization of LIME-identified genes for characterizing Parkinson’s disease pericytes. A)** Scatter plot showing the LIME feature importance Z-score for genes in the LIME-identified optimal gene set for pericytes. **B)** Volcano plot showing the mean LIME feature importance Z-score across the Kamath et al., Wang et al., and Smajic et al. datasets (Y-axis) and the gene expression log2 fold-change (Log2FC) between PD and control pericytes across all datasets (X-axis). Only genes comprising the LIME-identified optimal gene set for pericytes are shown. **C)** Heatmap showing the mean LIME feature importance Z-score for genes comprising the pericytes LIME-identified optimal gene set with an absolute value Log2FC > 0.25. In the heatmap, grey tiles indicate that the gene was not included in the cell type-specific optimal gene set. Abbreviations: DaNeurons, dopaminergic neurons; oligo, oligodendrocytes; OPC, oligodendrocyte precursor cells.

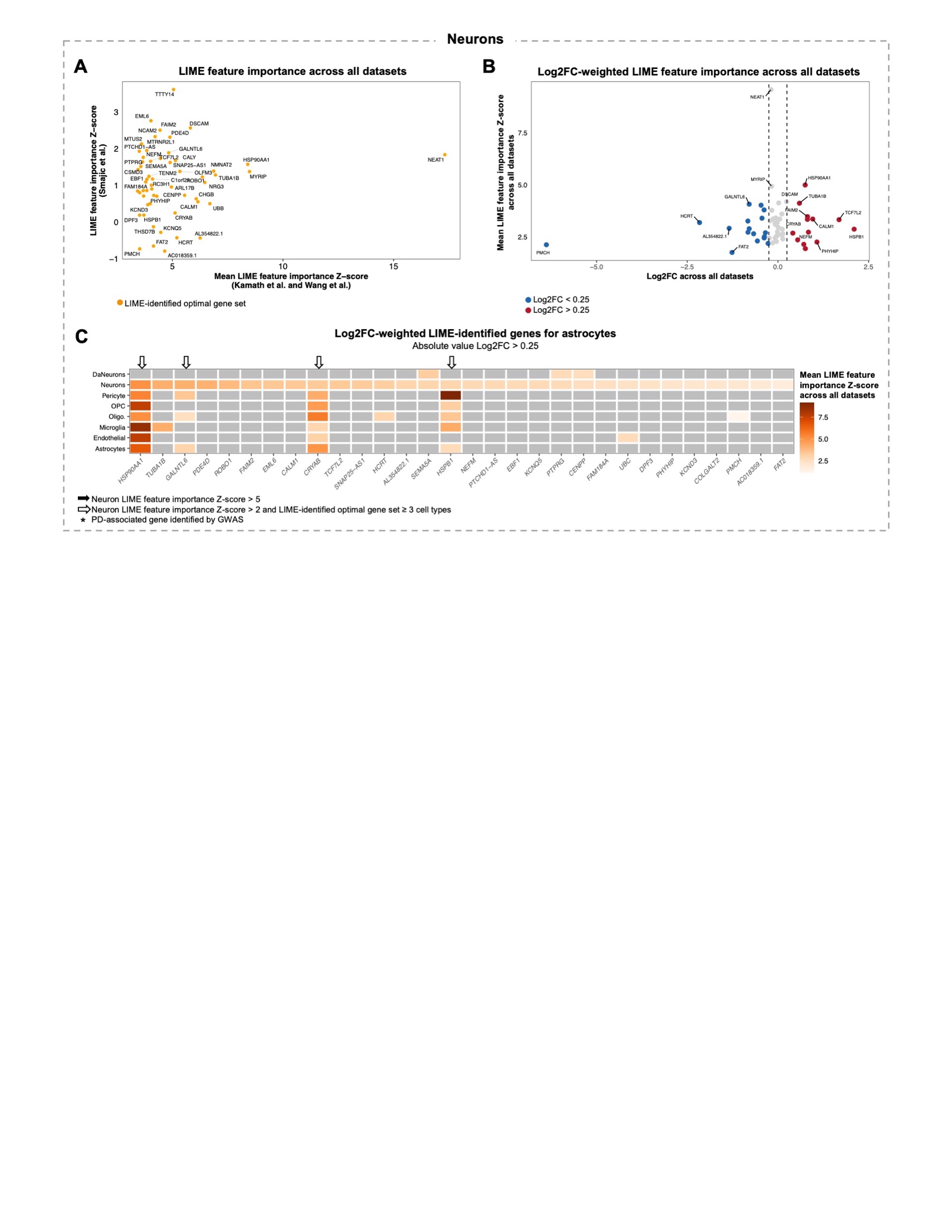

**Figure S23. Prioritization of LIME-identified genes for characterizing Parkinson’s disease neurons. A)** Scatter plot showing the LIME feature importance Z-score for genes in the LIME-identified optimal gene set for neurons. **B)** Volcano plot showing the mean LIME feature importance Z-score across the Kamath et al., Wang et al., and Smajic et al. datasets (Y-axis) and the gene expression log2 fold-change (Log2FC) between PD and control neurons across all datasets (X-axis). Only genes comprising the LIME-identified optimal gene set for neurons are shown. **C)** Heatmap showing the mean LIME feature importance Z-score for genes comprising the neurons LIME-identified optimal gene set with an absolute value Log2FC > 0.25. In the heatmap, grey tiles indicate that the gene was not included in the cell type-specific optimal gene set. Abbreviations: DaNeurons, dopaminergic neurons; oligo, oligodendrocytes; OPC, oligodendrocyte precursor cells.

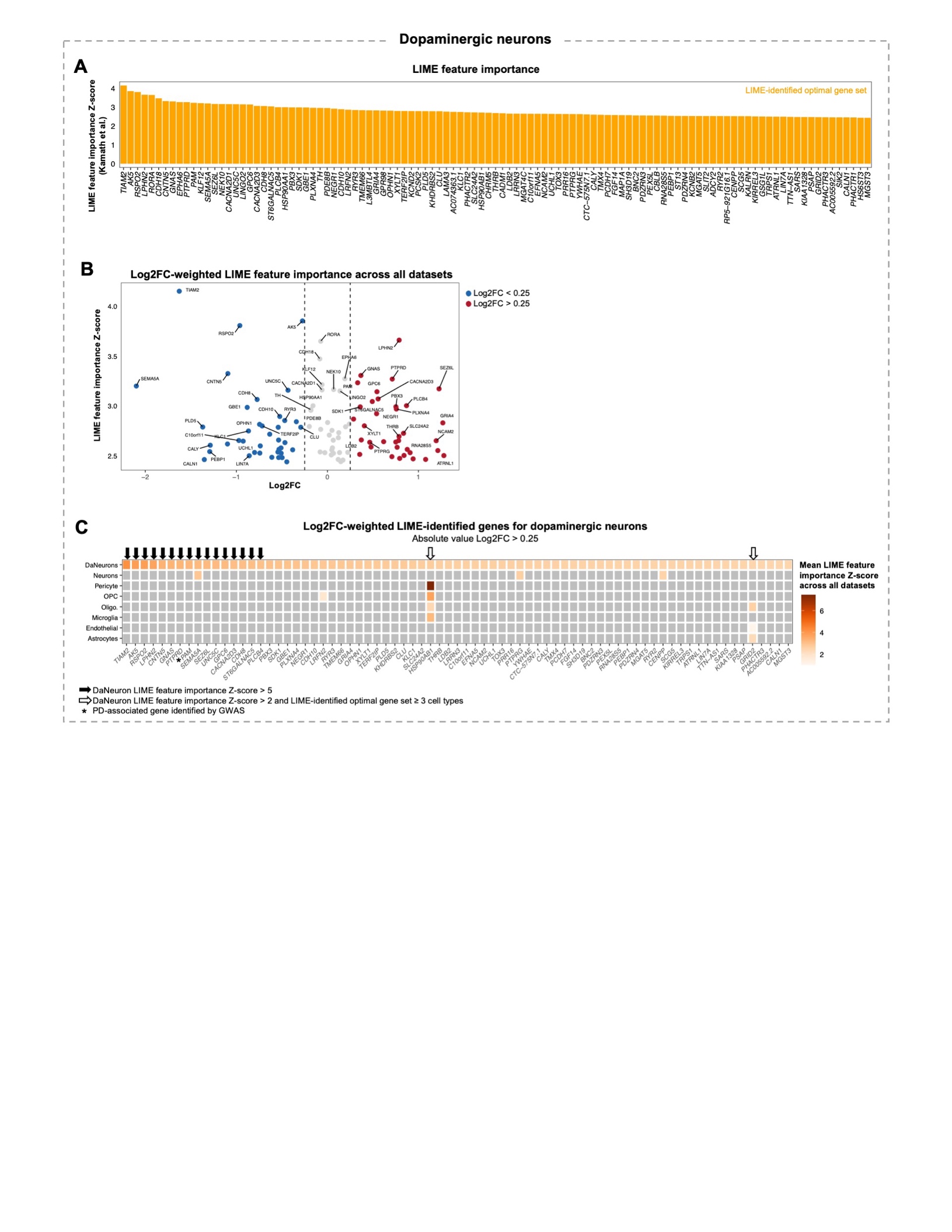

**Figure S24. Prioritization of LIME-identified genes for characterizing Parkinson’s disease dopaminergic neurons. A)** Bar plot showing the LIME feature importance Z-score in Kamath et al. genes comprising the LIME-identified optimal gene set for dopaminergic neurons (DaNeurons). **B)** Volcano plot showing the LIME feature importance Z-score in the Kamath et al. dataset (Y-axis) and the gene expression log2 fold-change (Log2FC) between PD and control DaNeurons (X-axis). Only genes comprising the LIME-identified optimal gene set for DaNeurons are shown. **C)** Heatmap showing the mean LIME feature importance Z-score for genes comprising the DaNeurons LIME-identified optimal gene set with an absolute value Log2FC > 0.25. In the heatmap, grey tiles indicate that the gene was not included in the cell type-specific optimal gene set. Abbreviations: oligo, oligodendrocytes; OPC, oligodendrocyte precursor cells.

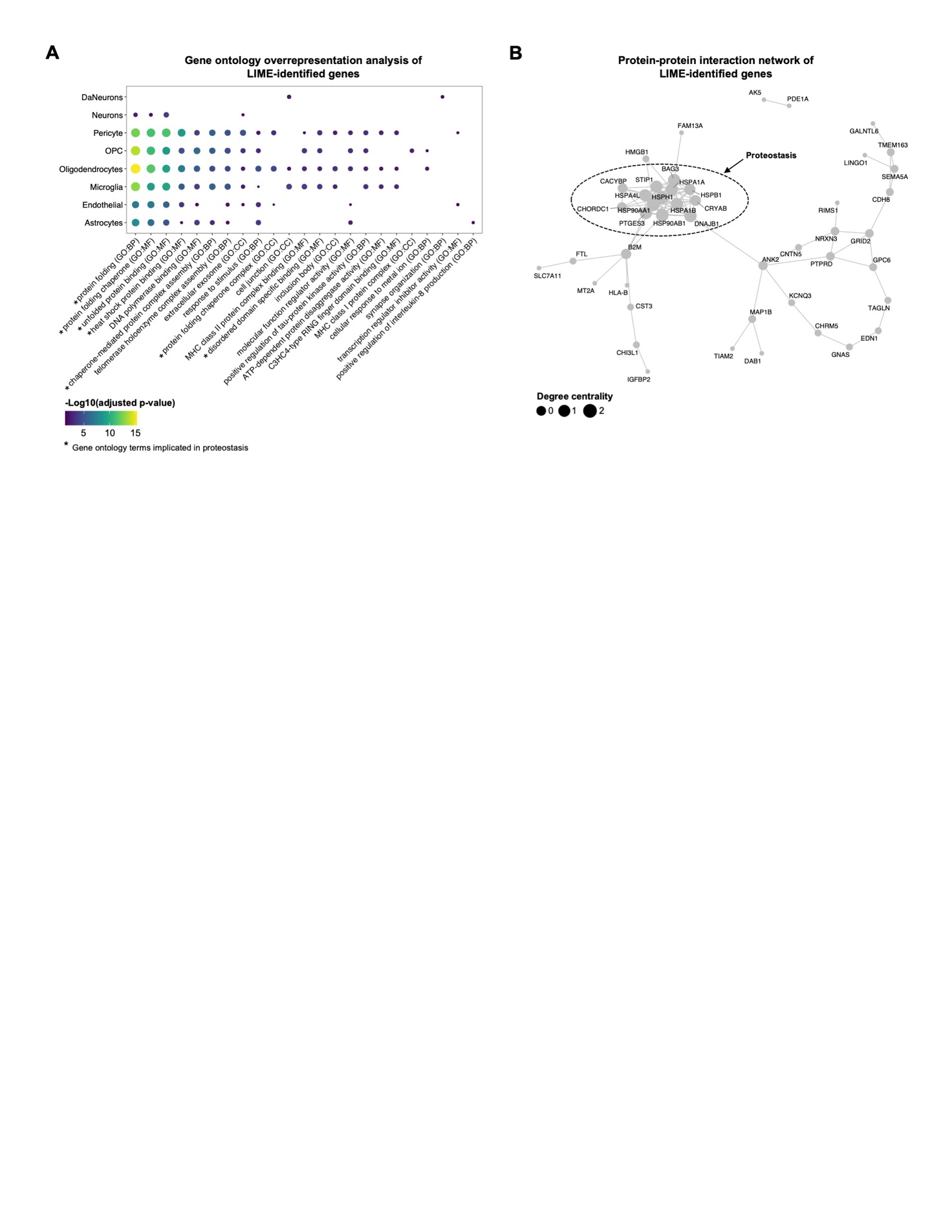

**Figure S25. Functional characterization of LIME-identified genes.** The final LIME-identified gene set comprised 66 unique genes across all cell types. **A)** Significantly overrepresented terms across all LIME-identified genes (*n =* 66) were identified using *gprofiler.* Terms from the *Gene Ontology Biological Processes* (GO:BP), and *Gene Ontology Cellular Component* (GO:CC) were considered significantly overrepresented if the adjusted p-value was < 0.05. The dot plot shows the enrichment of terms across the final LIME-identified genes for each specific cell type. Missing dots indicate that the specific term was not significantly enriched in the LIME-identified genes for corresponding cell type. **B)** Protein-protein interaction network informed by the *STRING* database. The size of the points represent the degree centrality, which is a measure of the connectedness of a node based on the number of direct connections it has to other nodes. Genes implicated in terms related to proteostasis are highlighted.

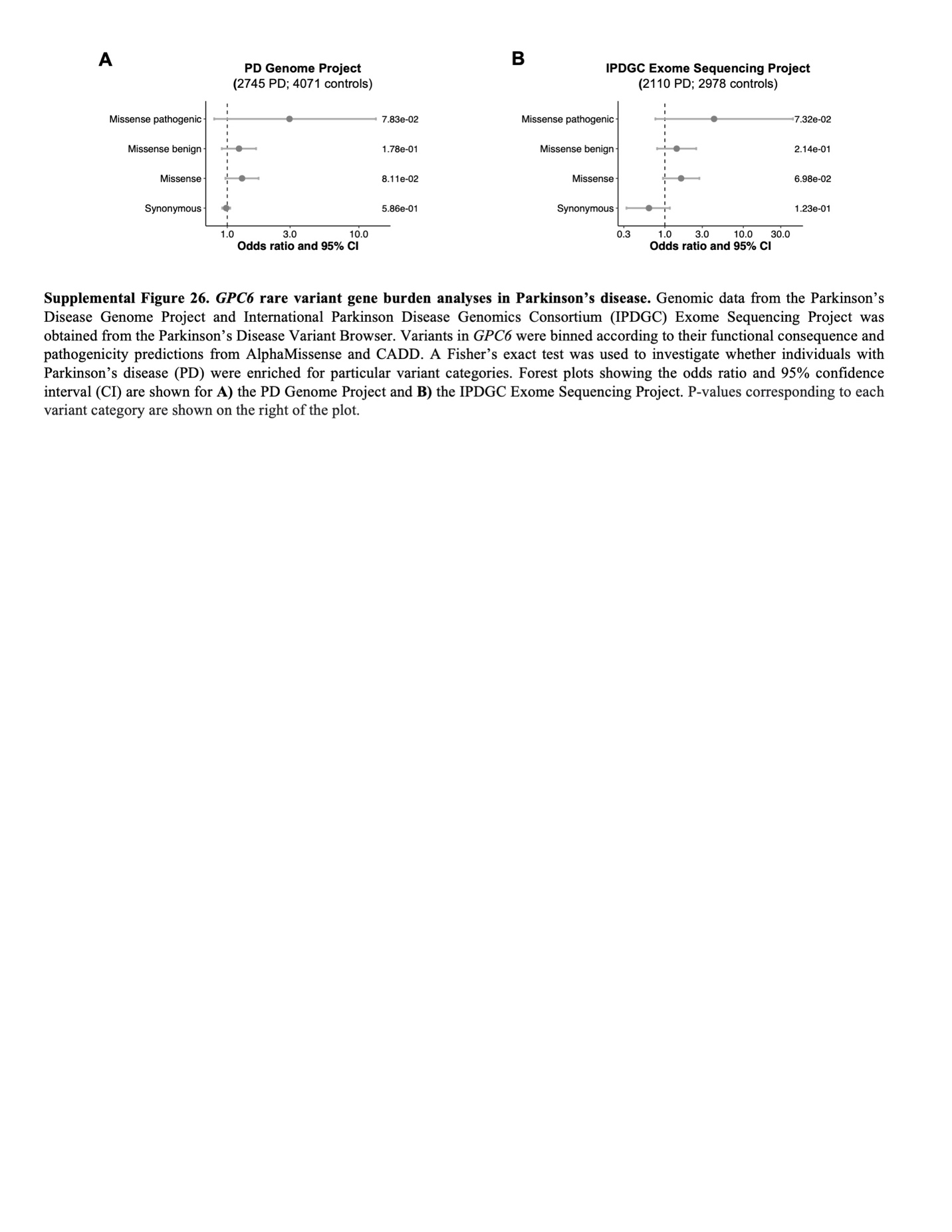

**Figure S26. *GPC6* rare variant gene burden analysis in Parkinson’s disease.** Fisher’s exact tests were performed to determine if individuals with Parkinson’s disease from two separate cohorts were enriched for rare variants in *GPC6* compared to healthy controls*.* P-values corresponding to each variant category are shown on the right of the plot. Forest plots showing the results of the Fisher’s exact test are shown for **A)** the Parkinson’s Disease Genome Project cohort and B**)** the International Parkinson’s Disease Genomics Consortium (IPDGC) Exome Sequencing Project cohort. Abbreviations: CI, confidence interval.
